# Cost-Effectiveness Analysis of Nirsevimab and RSVpreF Vaccine Prevention Strategies for Respiratory Syncytial Virus Disease in infants: A Canadian Immunisation Research Network (CIRN) Study

**DOI:** 10.1101/2023.07.14.23292675

**Authors:** Affan Shoukat, Elaheh Abdollahi, Alison P. Galvani, Scott A. Halperin, Joanne M. Langley, Seyed M. Moghadas

**Affiliations:** Agent-Based Modelling Laboratory, York University, Toronto, Ontario, Canada; Center for Infectious Disease Modeling and Analysis (CIDMA), Yale School of Public Health, New Haven, CT, USA; Canadian Center for Vaccinology, Dalhousie University, IWK Health and Nova Scotia Health, Halifax, Nova Scotia, Canada

**Keywords:** RSV, nirsevimab, RSVpreF vaccine, simulation, cost-effectiveness

## Abstract

**Background:** The cost-effectiveness of immunisation strategies with a long-acting monoclonal antibody (nirsevimab) and/or a protein-based maternal vaccine (RSVpreF) for protecting infants from Respiratory Syncytial Virus (RSV)-associated illness has not been previously determined for Canada. We estimated the health benefits and cost-effectiveness of nirsevimab for immunising the entire birth cohort regardless of gestational age or other risk factors. Additionally, we evaluated a combined strategy of year-round vaccination of pregnant women with RSVpreF and immunisation of high-risk infants with nirsevimab during RSV season.

**Methods:** We developed a discrete-event simulation model, parameterized with the data on RSV incidence, outpatient care, hospitalisations, and deaths. Intervention scenarios targeting twelve monthly birth cohorts and pregnant women were evaluated over a time horizon of one year. Taking into account the costs associated with RSV-related outcomes, we calculated the net monetary benefit using the quality-adjusted life-year (QALY) gained. Further, we determined the range of price-per-dose (PPD) for nirsevimab and RSVpreF within which the program was cost-effective. Cost-effectiveness analyses were conducted from both healthcare and societal perspectives.

**Findings:** Using a willingness-to-pay of CAD$50,000 per QALY gained, we found that immunising the entire birth cohort with nirsevimab would be cost-effective from a societal perspective for a PPD of up to $290, with an annual budget impact of $83,978 for 1,113 infants per 100,000 population. An alternative, combined strategy of vaccinating pregnant women and immunising only high-risk infants would lead to a lower budget impact of $49,473 per 100,000 population with a PPD of $290 and $195 for nirsevimab and RSVpreF, respectively. This combined strategy would reduce infant mortality by 76% to 85%, comparable to 78% reduction achieved through a nirsevimab-only program for immunising the entire birth cohort. PPD for cost-effective programs with nirsevimab was sensitive to the target population among infants.

**Interpretation:** Passive immunisation of infants under 6 months of age with nirsevimab and vaccination of pregnant women with RSVpreF could be a cost-effective strategy for protecting infants during their first RSV season.

**Funding:** This study was supported by the Canadian Immunisation Research Network (CIRN) and the Canadian Institutes of Health Research (CIHR). Seyed M. Moghadas acknowledges support from the Natural Sciences and Engineering Research Council of Canada (MfPH and Discovery grants). Alison P. Galvani acknowledges support from the The Notsew Orm Sands Foundation.

**Research in context:** *Evidence before this study:* Prevention of RSV disease in infants under 1 year of age has relied on palivizumab, a short-acting monoclonal antibody, administered monthly to high-risk infants during the period in which RSV is circulating in annual epidemics. New preventive measures including nirsevimab (a long-acting monoclonal antibody for immunising infants) and RSVpreF (a protein-based vaccine for immunising pregnant women) have been developed to reduce the risk of severe RSV illness in the first six months of life. However, no prior study has evaluated cost-effectiveness of these interventions in Canada with recently available efficacy estimates from randomised controlled clinical trials.

*Added value of this study:* Using a discrete-event simulation model, we found that immunising the entire birth cohort with nirsevimab would be cost-effective from a societal perspective for a price per dose of up to $290. Year-round vaccination of pregnant women with RSVpreF, followed by immunising infants at high-risk of severe RSV disease with nirsevimab as a combined strategy required a lower budget impact compared to the nirsevimab-only program for the entire birth cohort during the RSV season, while averting similar RSV-related infant mortality.

*Implications of all the available evidence:* Prevention strategies against RSV disease in infants using nirsevimab and RSVpreF vaccine could be cost-effective. A combined strategy of these interventions could reduce the budget impact to the healthcare system.

## Introduction

Respiratory Syncytial Virus (RSV) is the most common cause of lower respiratory tract illness (LRTI) in children under five years old worldwide,^1–3^ with the highest burden in the first six months of life. In high income countries, 1 to 2 % of the birth cohort is hospitalised for care of RSV-associated illness. The case fatality rate of hospitalised children can reach up to 2.8%.^1^ The direct (e.g., outpatient and inpatient care) and indirect (e.g., loss of productivity, parental costs, and psychological health) costs of RSV disease among infants are substantial.^4–7^

In the absence of a preventive vaccine, efforts to curb the burden of RSV among infants in the last two decades have relied on passive immunisation with the anti-RSV monoclonal antibody palivizumab. Palivizumab is currently administered in five monthly doses to infants at high-risk of severe RSV disease during the local RSV epidemic season.^8^ With the advent of structure-based vaccinology,^9^ vaccine candidates are being developed across active- and passive-immunising platforms with the aim of protecting infants during the highest risk period directly or through maternal immunisation. For instance, nirsevimab is a long-acting monoclonal antibody to the RSV fusion protein in its pre-fusion conformation (preF)^10,11^ that has been recently authorised for single dose administration to infants in Europe and Canada. Another strategy to prevent RSV-associated illness in the first six months of life is immunisation of pregnant women with a preF RSV protein-based vaccine (RSVpreF), providing passive immunisation to newborns through transplacental antibody transfer.^12^ With the availability of these products, the landscape of RSV prevention and disease burden is likely to change. However, feasibility and cost-effectiveness of infant and maternal immunisation programs will play an important role in recommendations for use, such as providing long-acting monoclonal antibodies to the entire birth cohort during the RSV season, targeting high-risk infants only, vaccinating pregnant women, or a combination of these strategies.

In this study, we aimed to conduct a comprehensive cost-effectiveness analysis of RSV infant and maternal immunisation strategies based on population demographics in the Canadian south (i.e., southern provinces of Canada excluding the three northern territories and Nunavik in Quebec). We developed a discrete-event simulation model of RSV outcomes and calculated the net-monetary benefit (NMB), incremental cost-effectiveness ratio (ICER), and the budget impact associated with immunisation programs. Accounting for the efficacy of nirsevimab and RSVpreF against RSV-related outcomes in infants, as well as direct and indirect costs of health outcomes and program implementation, we performed cost-effectiveness analyses from both the publicly funded health system (referred to as healthcare) and societal perspectives.

## Methods

### Model structure and study population

We developed a discrete-event simulation model (**Figure 1**) with 1,113 infants per 100,000 population as the birth cohort, reflecting the 2021 census data for Ontario, Canada.^13^ Twelve monthly birth cohorts were followed through the first year of their life, categorised as preterm with <29 weeks of gestational age (wGA), 29-32 wGA, 33-36 wGA,^14^ and term infants with 37+ wGA.^15,16^ Preterm infants comprised ∼9% of the cohort, distributed as 7%, 17%, and 76% in the corresponding wGA.^14^ We also considered chronic lung disease (CLD) and congenital heart disease (CHD) as two major risk factors associated with RSV disease outcomes. The rate of CLD was set to 28.1%, 4%, and 2.4% for wGA <29, 29–32, and 33-36, respectively, among preterm infants.^17^ For CHD, we used an overall prevalence rate of 12.3 per 1000 live births in Canada.^18^

**Figure 1.**
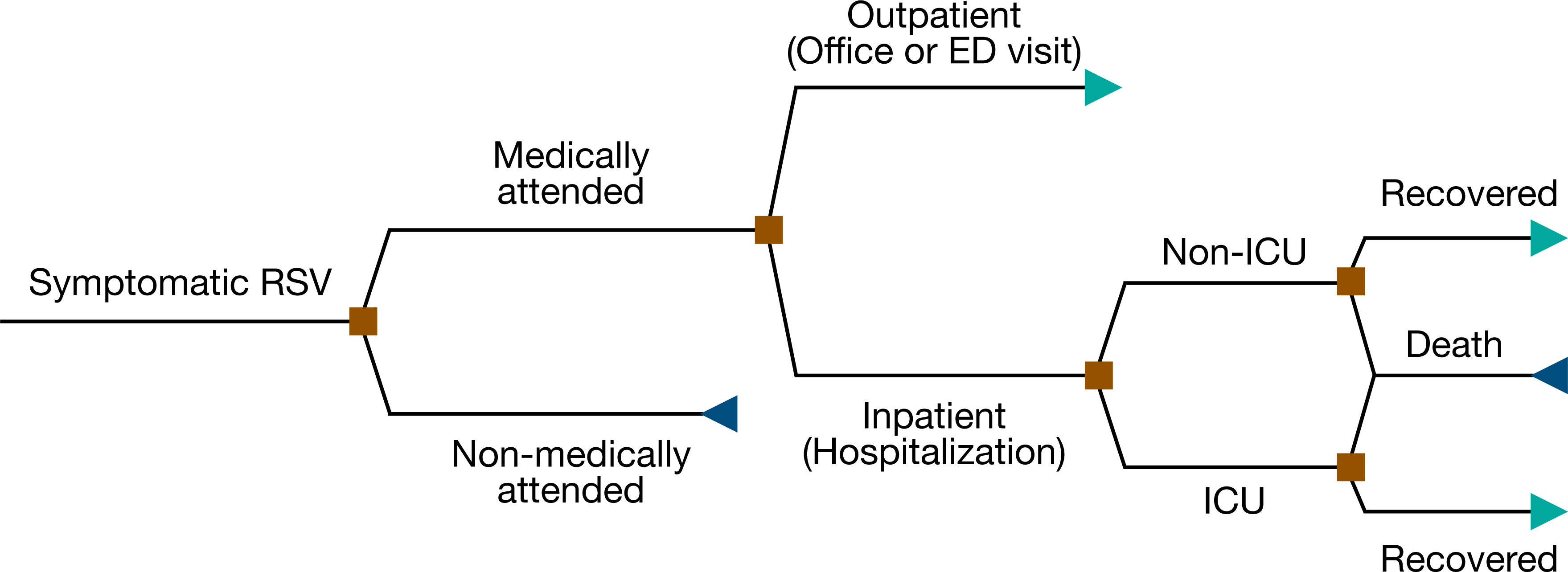
Structure of discrete-event simulation model applied to scenarios in the presence and absence of interventions with different outcomes.

### RSV-related outcomes

The model was parameterized with estimates of the burden of RSV disease in different chronologic and gestational age groups. The annual incidence of medically-attended (MA) RSV cases per 100,000 population was sampled from the range 1001 – 2439, and distributed among infants under one year of age according to estimated rates and seasonality distribution (**Supplementary Material, Figure A1, Table A2**).^7^ In our study, MA RSV refers to outpatient care (i.e., office visit or emergency department (ED) visit without hospital admission) or inpatient care (i.e., hospital admission in paediatric ward or intensive care unit, ICU). We considered the beginning of October as the start of RSV season recognizing that the RSV season varies geographically and temporally (**Supplementary Material, Figure A3**).^7,19^

We allowed for a maximum of two MA RSV events during the first year of life,^20^ with a minimum time-interval of three months between the two events if the second episode occurred. The duration of symptomatic RSV disease for those receiving outpatient care was sampled between 5 to 8 days.^21^ Hospitalisation rates for infants with MA RSV LRTI were based on their age at incidence as well as their wGA (**Supplementary Material, Figure A2, Table A3**). The likelihood of hospitalisation increased by 1.9 and 2.2 times for infants with CLD and CHD, respectively, compared to infants without these conditions (**Supplementary Material**).^22,23^

Among hospitalised cases, ICU admission varied in the range 41.3% – 62.1%, 13.1% – 53.6%, and 5.4% – 30.0% among infants of ≤32, 33–35, and ≥36 wGA, respectively.^21^ For infants of ≤32 wGA, the duration of hospitalisation was sampled from Gamma distributions, with mean values of 6.1 and 9.5 days in paediatric ward and ICU (**Table 1**), respectively.^21,24^ For infants born at 33 or higher wGA, we sampled the duration of stay in paediatric ward and ICU from Gamma distributions with mean values of 3.9 and 5.2 days, respectively (**Table 1**).^21,24,25^ The probability of experiencing a wheezing episode post hospitalisation was 0.31 during the first year of life.^26,27^ The duration of a wheezing episode ranged from 5.2 to 9.8 days.^26,28^ RSV-related mortality for hospitalised infants without CLD or CHD varied in the ranges 0.36% – 3.3%, 0.02% – 1.82%, and 0.02% – 1% for infants of ≤32, 33-35, and 36 or higher wGA, respectively.^29–35^ For hospitalised infants with CLD and CHD, mortality rates were 3.5% – 5.1% and 3.4% – 5.3%, respectively.^29^

**Table 1.**
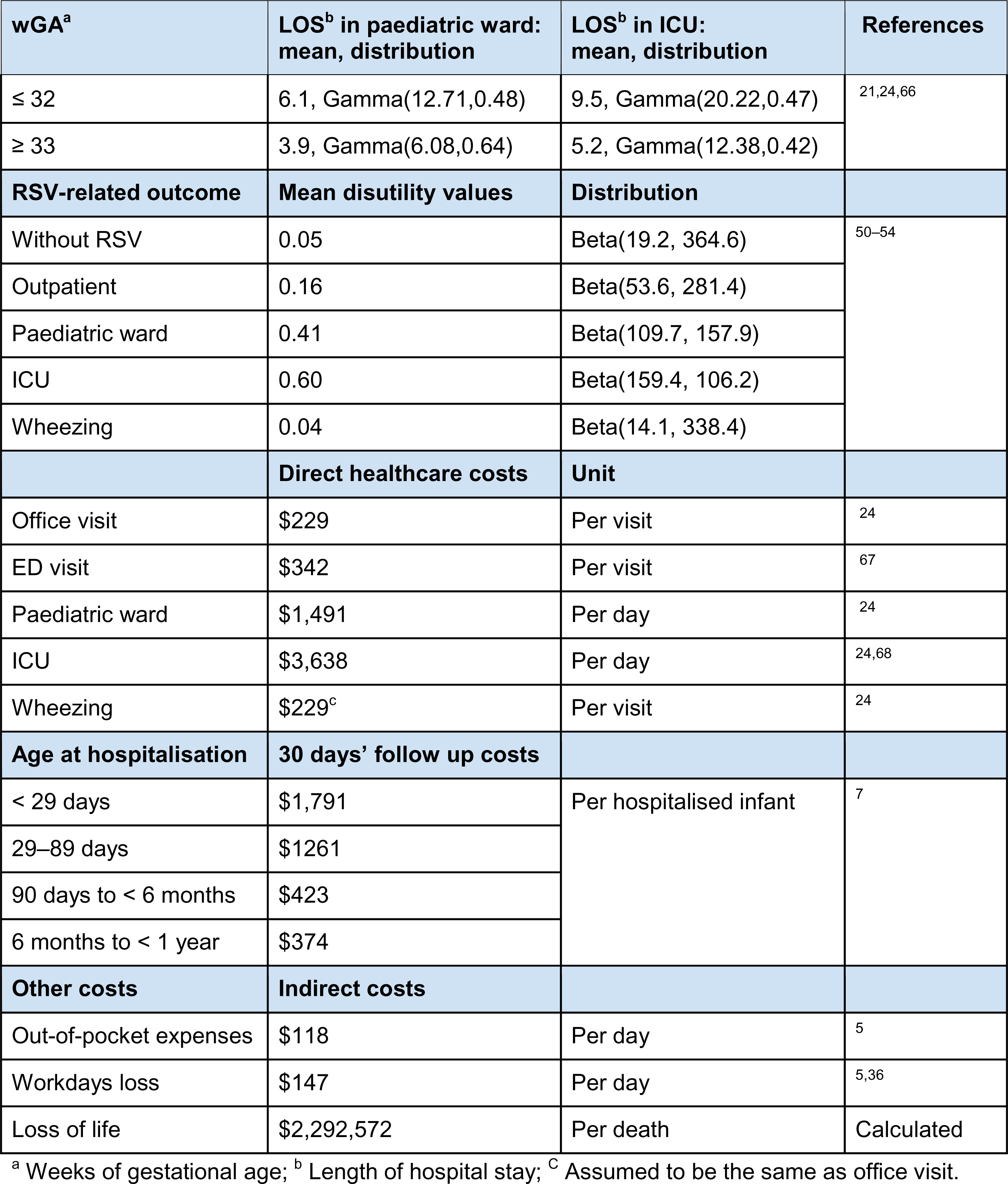
Model parameters used for cost-effectiveness analysis. All costs are inflated to 2023 Canadian dollars.

| **wGA<sup>a</sup>** | **LOS<sup>b</sup> in paediatric ward:**<br>**mean, distribution** | **LOS<sup>b</sup> in ICU:**<br>**mean, distribution** | **References** |
| --- | --- | --- | --- |
| ≤ 32 | 6.1, Gamma(12.71,0.48) | 9.5, Gamma(20.22,0.47) | 21,24,66 |
| ≥ 33 | 3.9, Gamma(6.08,0.64) | 5.2, Gamma(12.38,0.42) |  |
| **RSV-related outcome** | **Mean disutility values** | **Distribution** |  |
| Without RSV | 0.05 | Beta(19.2, 364.6) | 50–54 |
| Outpatient | 0.16 | Beta(53.6, 281.4) |  |
| Paediatric ward | 0.41 | Beta(109.7, 157.9) |  |
| ICU | 0.60 | Beta(159.4, 106.2) |  |
| Wheezing | 0.04 | Beta(14.1, 338.4) |  |
|  | **Direct healthcare costs** | **Unit** |  |
| Office visit | \$229 | Per visit | 24 |
| ED visit | \$342 | Per visit | 67 |
| Paediatric ward | \$1,491 | Per day | 24 |
| ICU | \$3,638 | Per day | 24,68 |
| Wheezing | \$229<sup>c</sup> | Per visit | 24 |
| **Age at hospitalisation** | **30 days' follow up costs** |  |  |
| < 29 days | \$1,791 | Per hospitalised infant | 7 |
| 29–89 days | \$1261 | | |
| 90 days to < 6 months | \$423 | | |
| 6 months to < 1 year | \$374 | | |
| **Other costs** | **Indirect costs** |  |  |
| Out-of-pocket expenses | \$118 | Per day | 5 |
| Workdays loss | \$147 | Per day | 5,36 |
| Loss of life | \$2,292,572 | Per death | Calculated |

<sup>a</sup> Weeks of gestational age; <sup>b</sup> Length of hospital stay; <sup>c</sup> Assumed to be the same as office visit.

### Costs of RSV-related outcomes

Direct costs borne by the healthcare system included office visit, ED visit, hospitalisation, as well as 30 days’ follow up for hospitalised infants (**Table 1**). Indirect costs included out-of-pocket expenses, loss of productivity by parents, and monetary loss of life due to RSV-related infant mortality. Out-of-pocket expenses for families with hospitalised infants were estimated at $118 per day for the duration of hospital stay to account for transportation, over-the-counter medications, meals, child care and other costs.^5^ Indirect costs related to workdays lost of working parents (with an average absenteeism of 49%)^5^ were calculated using the per capita personal income of CAD$53,675 per year (i.e., ∼$147 per day) in Ontario.^36^ We assumed total workdays lost were equal to the length of stay for hospitalised infants and one day for infants who required outpatient care.^5^ We considered the recommended 1.5% discounting rate by the Canadian Agency for Drugs and Technologies in Health,^37^ with an average lifespan of 82 years. Each RSV-related death was estimated to have a total discounted monetary loss of $2,292,572, calculated using the annual personal income, and discounted quality-adjusted life-year (QALY) loss of 45.3. All costs were converted and inflated to 2023 Canadian dollars.

### Infant and maternal RSV prevention strategies

Although year-round RSV activity was implemented in the model according to reported incidence and outcomes (**Supplementary Material, Figure A1**),^7,19^ we considered infant immunisation with nirsevimab to start in October, corresponding to the start of RSV season (**Supplementary Material, Figure A3**). Infants born off-season were immunised at the start of the RSV season following their birth. Based on the current recommendation for use of palivizumab, which is directed at preterm and selected high-risk infants,^21^ we evaluated the following program options (**Table 2**) for passive immunisation with nirsevimab: (i) preterm infants ≤32 wGA and infants with CLD or CHD condition (L1); (ii) preterm infants ≤36 wGA and infants with CLD or CHD condition (L2); (iii) all preterm infants (<37 wGA), infants with CLD or CHD, and term infants born during RSV season (L3); and (iv) the entire birth cohort (L3). The coverage for these immunisation programs was set to 100% for the base-case analysis, but reduced to 80% for the secondary analysis (**Supplementary Material**).

**Table 2.**
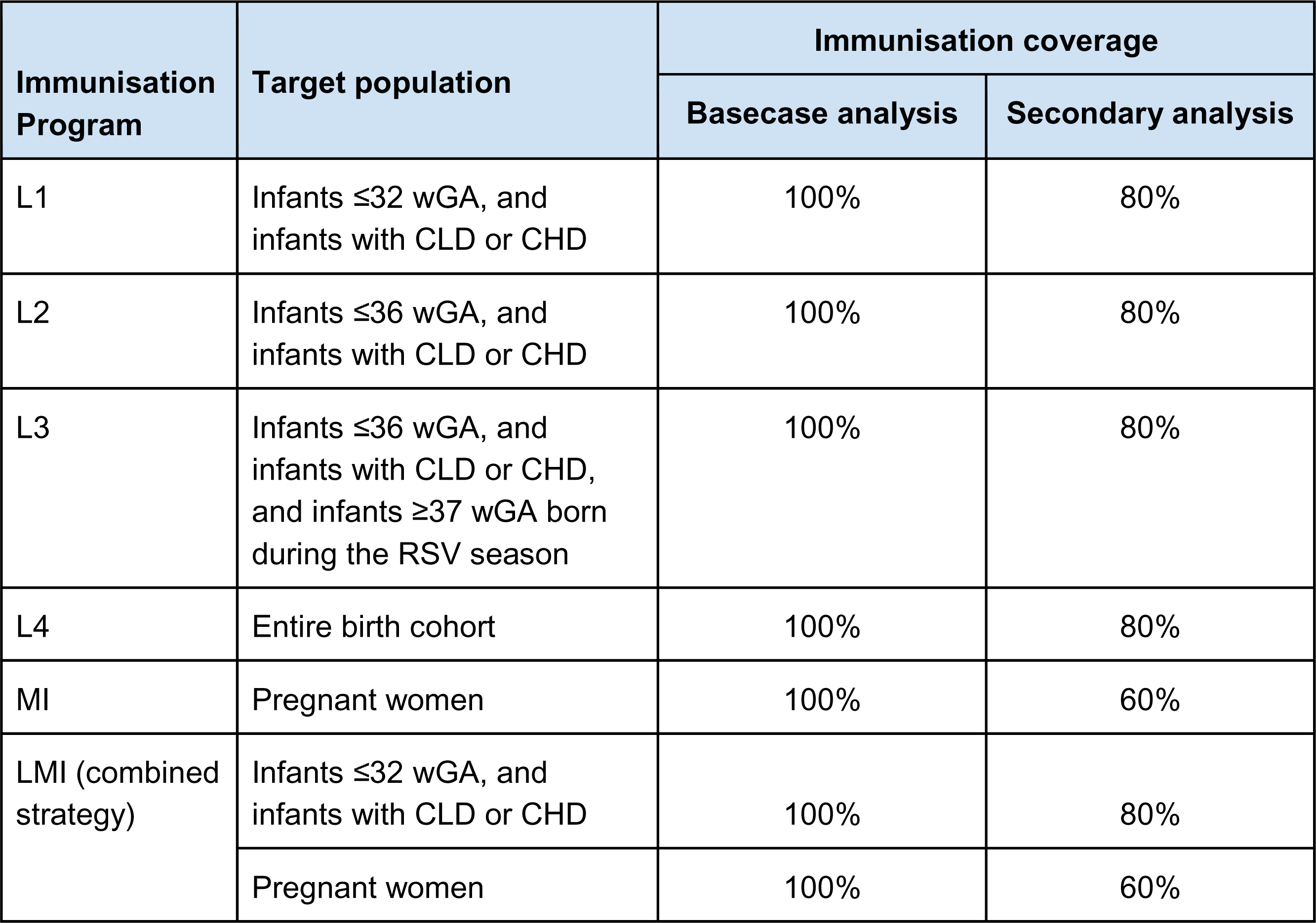
Summary of immunisation programs, target populations, and immunisation coverages.

Maternal immunisation (MI) was implemented as a year-round program, with vaccination of pregnant women who are in their last trimester before gestation week 33 (**Supplementary Material, Figure A3**). In the base-case analysis, vaccination coverage was set to 100%. For the secondary analysis, we assumed a 60% coverage based on estimates of 2021 vaccination coverage against influenza and pertussis in pregnant women in Canada (**Supplementary Material**).^38^

To evaluate the combination of nirsevimab and RSVpreF, we implemented a program (LMI) that includes year-round vaccination of pregnant women followed by administration of nirsevimab to infants at high-risk of severe RSV disease (i.e., preterm infants ≤32 wGA and infants with CLD or CHD condition) during RSV season. Table 2 summarises all the immunisation programs, target populations, and coverages for base-case and secondary analyses.

### Efficacy of nirsevimab and RSVpreF vaccine

We considered the efficacy of nirsevimab and RSVpreF against MA RSV LRTI and severe RSV LRTI. Conservatively, no efficacy against RSV infection or symptomatic RSV disease was assumed. The efficacy of a single dose of nirsevimab against MA RSV-LRTI is estimated at 79.5% (95% CI: 65.9% to 87.7%) through 150 days post-dose.^39^ Mean efficacies against hospitalisation and very severe RSV LRTI (used against ICU admission in our model) are estimated at 77.3% (95% CI: 50.3% to 89.7%) and 86% (95% CI: 62.5% to 94.8%), respectively.^39^

We employed a sigmoidal decay to temporally disaggregate the constant efficacy values for up to 10 months,^40^ while maintaining the same mean efficacy for the first 5 months as estimated in clinical trials (**Supplementary Material, Figure A4**). As sensitivity analysis, we used constant vaccine efficacy profiles with mean estimates as reported in clinical trials, and a linear decline beginning at 5 months post immunisation (**Supplementary Material, Figure A4**).

The efficacy of RSVpreF is estimated at 57.1% (95% CI: 14.7% to 79.8%) against MA RSV LRTI, 67.9% (95% CI: 34.6% to 84.2%) against hospitalisation, and 81.8% (95% CI: 40.6% to 96.3%) against severe MA RSV LRTI (used against ICU admission in our model) for the first 90 days of life.^12,41–43^ Similar to nirsevimab, we used a sigmoidal decay to determine temporal vaccine efficacy over 10 months, with the same mean efficacy as estimated in clinical trials for the first 3 months after birth (**Supplementary Material, Figure A4**). We also performed a sensitivity analysis using constant vaccine efficacy profiles with mean estimates from clinical trials, and a linear decline starting 3 months after birth (**Supplementary Material, Figure A4**).

### Costs of RSV prevention strategies

We varied the single-dose cost of both nirsevimab and RSVpreF between $50 and $1000 to determine the price range within which an immunisation program would be cost-effective. Costs associated with dose administration was set to $15 for both infant and maternal immunisation.^44,45^

### Cost-effectiveness analysis

To determine whether a program was cost-effective for a given willingness-to-pay (WTP) threshold, we calculated the net monetary benefit (NMB) by **NMB = Δ*E* × WTP − Δ*C***, where **Δ*E*** represents QALYs gained using intervention compared to no intervention, and **Δ*C*** is the incremental costs.^46^ A program was considered cost-effective if it resulted in a positive NMB. In the primary analysis, we calculated the monetary value of health using a WTP threshold of $50,000 per QALY gain.^47^ In secondary analyses (**Supplementary Material**), we considered a lower threshold of $30,000^48^ and a higher threshold of $70,000^49^ corresponding to the per capita gross domestic product in Canada. We also estimated the ICER for each intervention as **Δ*C*/Δ*E***, which provides a metric to measure the additional costs required to gain one QALY. Disutility values of RSV-related outcomes were sampled individually for each RSV case from their respective distributions (**Table 1**), and adjusted for the duration of illness and outcomes.^50–54^ Utility values were calculated as (1 – sampled disutility) and used to derive total QALYs in each scenario by adding utility values during the illness and outside the illness duration in one year of life. We sampled a baseline disutility, and calculated the utility without RSV (**Table 1**), accounting for non-RSV health related illnesses.^55^ When immunisation was effective against MA RSV LRTI, preventing outpatient, we considered adjusted utility values for the duration of symptomatic RSV disease in non-MA infants. The distribution of QALY loss calculated using sampled disutility values were consistent with recent estimates.^56^

We considered both healthcare and societal perspectives for cost-effectiveness analysis. The healthcare perspective included all direct medical costs of RSV-related disease and the immunisation program during the first year of life. In the base-case analysis, the societal perspective incorporated direct and indirect costs in the calculation of NMB and ICER, including productivity loss of parents, without considering the monetary loss of life due to RSV-related infant mortality. In the secondary analysis, we also included the monetary loss of life due infant mortality in the societal perspective (**Supplementary Material**). Based on the results of cost-effectiveness analyses, we determined the budget impact of each immunisation program as the difference between immunisation costs and the total direct healthcare savings achieved in the program.

### Model implementation

For each scenario, the model was simulated stochastically using Monte-Carlo sampling for a total of 1000 realisations. All parameters were sampled from their respective distributions and individually for each infant, thus probabilistically accounting for the sensitivity of the model outcomes with respect to input parameters. For parameters for which a statistical distribution was unknown, we sampled uniformly from the estimated ranges. Point estimates of the model outcomes reflected the mean value of the 1000 Monte Carlo simulations. The uncertainty around the point estimates were derived using a nonparametric, bias-corrected and accelerated bootstrap technique with 1000 replicates, and 95% confidence intervals for the mean of estimates were constructed in scenarios evaluated. The computational model is available at https://github.com/affans/rsv_costeffectiveness

### Ethics and guidelines

This study used publicly available estimates and data sources and thus no ethics approval was required. We followed guidelines set forth by the Consolidated Health Economic Evaluation Reporting Standards (CHEERS).^57^

## Results

We estimated the reduction of health outcomes and performed cost-effectiveness analyses of interventions for twelve monthly birth cohorts, followed through the first year of their life.

### Health outcomes with sigmoidal vaccine efficacy profiles

For immunisation strategies in Table 1, we estimated that L1 would reduce RSV-related outpatient care by 2.0% (95% CI: 2.0% to 2.0%) and inpatient care by 6.2% (95% CI: 5.7% to 6.6%) in the base-case analysis (**Figure 2A**). Program extension to all preterm infants in L2 provided a marginal improvement in the reduction of outpatient care at 5.9% (95% CI: 5.8% to 5.9%) and inpatient care at 11.1% (95% CI: 10.6% to 11.6%). L3 was associated with a reduction of 38.9% (95% CI: 38.8% to 39.0%) outpatient care and 61.2% (95% CI: 60.4% to 62.1%) inpatient care. Administration of nirsevimab to the entire birth cohort in L4 reduced outpatient care by 63.4% (95% CI: 63.2% to 63.5%), and inpatient care by 79.3% (95% CI: 78.7% to 80.1%). The reduction in RSV-related infant mortality was 24.3% (95% CI: 16.9% to 33.2%) in L1, 36.3% (95% CI: 26.8% to 46.5%) in L2, 67.9% (95% CI: 58.8% to 77.3%) in L3, and 77.8% (95% CI: 69.6% to 85.3%) in L4 (**Supplementary Material, Table A6**).

**Figure 2.**
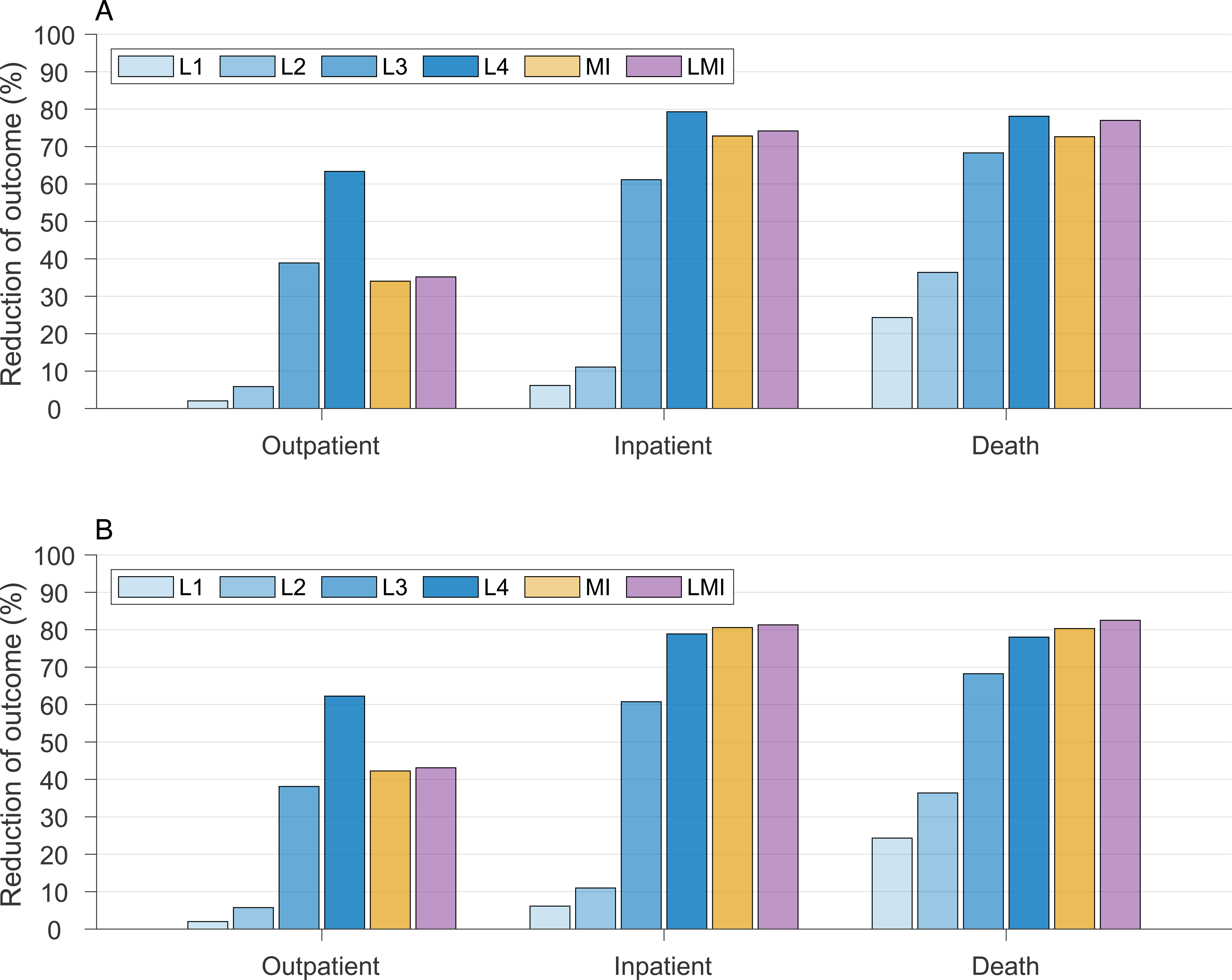
Overall reduction of RSV-related outpatient care (office and ED visits), inpatient care (paediatric ward and ICU admissions), and death among infants under one year of age for standalone immunisation programs with nirsevimab (L1, L2, L3, L4) and RSVpreF (MI), and combined nirsevimab and RSV-preF immunisation program (LMI), compared to the scenario without any prevention strategy. Panel (A) and (B) correspond to the sigmoidal and constant vaccine efficacy profiles, respectively.

MI was estimated to reduce RSV-related outpatient care by 34.0% (95% CI: 33.9% to 34.2%), inpatient care by 72.8% (95% CI: 72.1% to 73.5%), and death by 72.4% (95% CI: 62.5% to 81.9%) (**Figure 2A**). For the immunisation program combining administration of nirsevimab and RSVpreF (LMI), we estimated a reduction of 35.2% (95% CI: 35.0% to 35.3%) for outpatient care, 74.1% (95% CI: 73.5% to 74.9%) for inpatient care, and 76.8% (95% CI: 67.1% to 85.8%) for death, compared with no intervention (**Supplementary Material, Table A6**).

### Health outcomes with constant vaccine efficacy profiles

In the base-case analysis, we estimated that L1 would reduce RSV-related outpatient and inpatient care by 2.0% (95% CI: 2.0% to 2.0%) and 6.1% (95% CI: 5.7% to 6.6%), respectively (**Figure 2B**). Program extension to all preterm infants in L2 provided a reduction of 5.8% (95% CI: 5.7% to 5.8%) in outpatient care and 11.0% (95% CI: 10.5% to 11.6%) for inpatient care. L3 was associated with a reduction of 38.1% (95% CI: 38.0% to 38.2%) in outpatient care and 60.8% (95% CI: 60.0% to 61.7%) for inpatient care. Immunising the entire birth cohort with nirsevimab in L4 reduced outpatient care by 62.3% (95% CI: 62.1% to 62.4%) and inpatient care by 78.9% (95% CI: 78.2% to 79.6%). The reductions in RSV-related infant mortality were estimated to be the same as the corresponding nirsevimab immunisation programs using sigmoidal vaccine efficacy profiles.

MI was estimated to reduce RSV-related outpatient care by 42.3% (95% CI: 42.1% to 42.4%), inpatient care by 80.6% (95% CI: 80.0% to 81.2%), and death by 82.1% (95% CI: 71.4% to 88.9%) (**Figure 2B**). For the combined immunisation program, we estimated that LMI would reduce outpatient care by 43.1% (95% CI: 43.0% to 43.3%), inpatient care by 81.3% (95% CI: 80.6% to 81.9%), and death by 82.3% (95% CI: 74.0% to 90.2%), compared with no intervention (**Supplementary Material, Table A6**).

### Cost-effectiveness of standalone nirsevimab and RSVpreF prevention programs

We determined the price per dose (PPD) of nirsevimab below which the standalone infant immunisation programs were cost-effective at the WTP of $50,000 per QALY gained. From a healthcare perspective (**Table 3**), the maximum PPD for a positive NMB was $615 in L1, and reduced to $375 in L2, $300 in L3, and $215 in L4 using sigmoidal vaccine efficacy profiles (**Figure 3C**). Corresponding to these PPDs, the probabilities of L1, L2, L3, and L4 being cost-effective were 50%, 56%, 79%, and 99%, respectively. For MI, the maximum PPD was $160, at which the program was cost-effective with the probability of 68%. From a societal perspective (**Table 3**) with sigmoidal vaccine efficacy profiles, the maximum PPD for a positive NMB was estimated to be $705 in L1, $455 in L2, $385 in L3, and $290 in L4 (**Figure 3D**). The probabilities of these programs being cost-effective at their maximum PPD were 52%, 51%, 83%, and 55% in L1, L2, L3, and L4, respectively. MI was cost-effective for a PPD up to $200, with the probability of 87%.

**Table 3.**
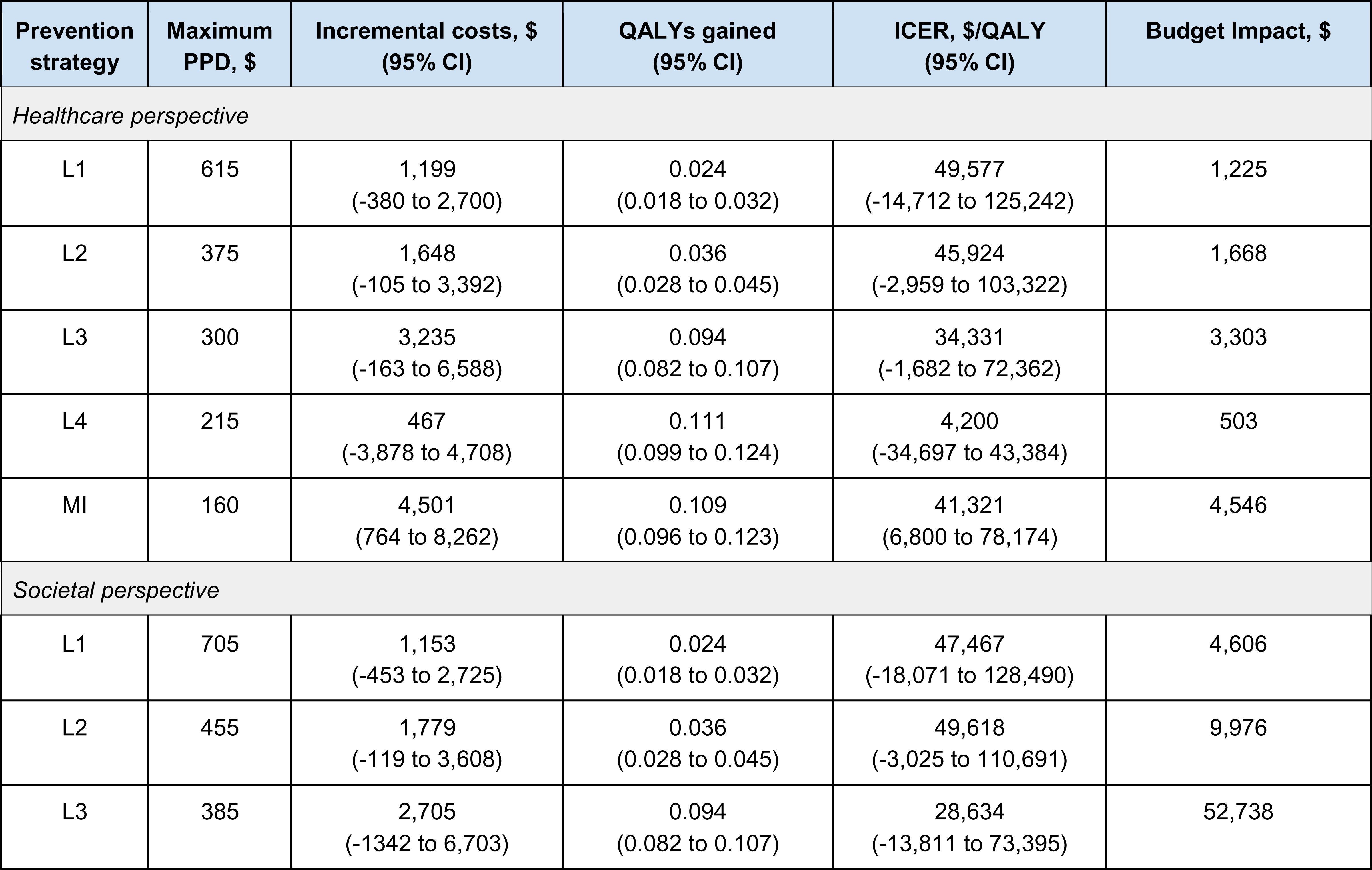
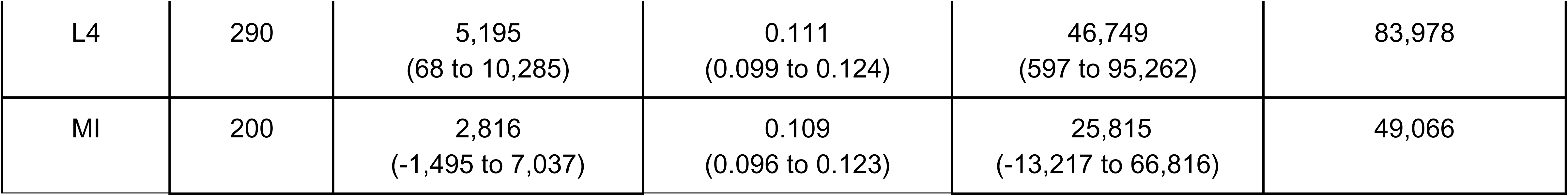
Model estimates of cost-effectiveness analyses associated with infant and maternal immunisation programs as standalone prevention strategies from healthcare and societal perspectives at the WTP of $50,000 using sigmoidal vaccine efficacy profiles. All strategies were compared to the baseline with no intervention.

| Prevention strategy | Maximum PPD, \$ | Incremental costs, \$ (95% CI) | QALYs gained (95% CI) | ICER, \$/QALY (95% CI) | Budget Impact, \$ |
| --- | --- | --- | --- | --- | --- |
| *Healthcare perspective* |  |  |  |  |  |
| L1 | 615 | 1,199<br>(-380 to 2,700) | 0.024<br>(0.018 to 0.032) | 49,577<br>(-14,712 to 125,242) | 1,225 |
| L2 | 375 | 1,648<br>(-105 to 3,392) | 0.036<br>(0.028 to 0.045) | 45,924<br>(-2,959 to 103,322) | 1,668 |
| L3 | 300 | 3,235<br>(-163 to 6,588) | 0.094<br>(0.082 to 0.107) | 34,331<br>(-1,682 to 72,362) | 3,303 |
| L4 | 215 | 467<br>(-3,878 to 4,708) | 0.111<br>(0.099 to 0.124) | 4,200<br>(-34,697 to 43,384) | 503 |
| MI | 160 | 4,501<br>(764 to 8,262) | 0.109<br>(0.096 to 0.123) | 41,321<br>(6,800 to 78,174) | 4,546 |
| *Societal perspective* |  |  |  |  |  |
| L1 | 705 | 1,153<br>(-453 to 2,725) | 0.024<br>(0.018 to 0.032) | 47,467<br>(-18,071 to 128,490) | 4,606 |
| L2 | 455 | 1,779<br>(-119 to 3,608) | 0.036<br>(0.028 to 0.045) | 49,618<br>(-3,025 to 110,691) | 9,976 |
| L3 | 385 | 2,705<br>(-1342 to 6,703) | 0.094<br>(0.082 to 0.107) | 28,634<br>(-13,811 to 73,395) | 52,738 |
| L4 | 290 | 5,195<br>(68 to 10,285) | 0.111<br>(0.099 to 0.124) | 46,749<br>(597 to 95,262) | 83,978 |
| MI | 200 | 2,816<br>(-1,495 to 7,037) | 0.109<br>(0.096 to 0.123) | 25,815<br>(-13,217 to 66,816) | 49,066 |

**Figure 3.**
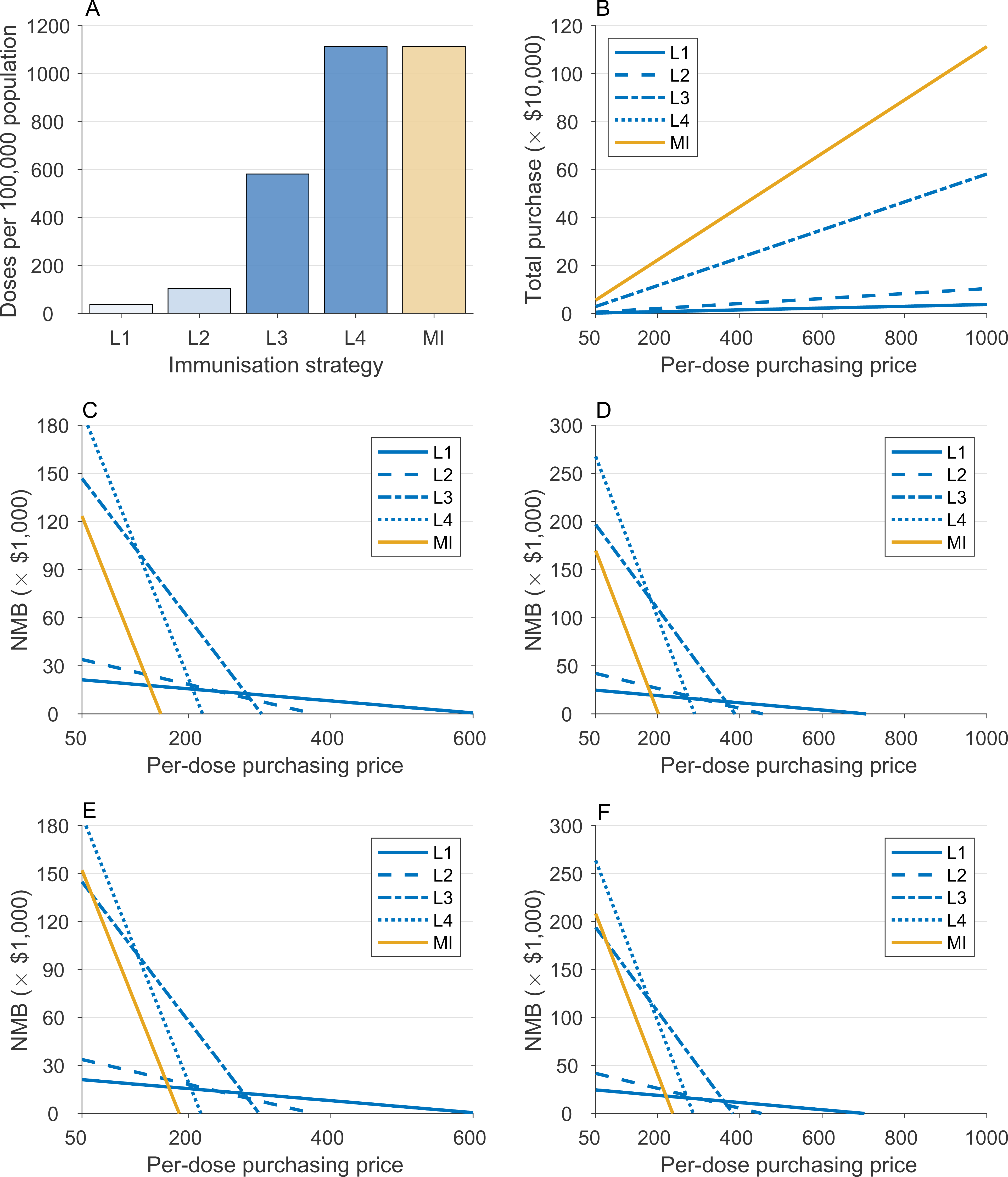
Required doses of nirsevimab and RSVpreF per 100,000 population for immunisation strategies (A), with total purchasing costs (B), and the estimated net monetary benefit (NMB) as a function of price per dose at the WTP threshold of $50,000 per QALY gained. Panels (C) and (D) correspond to the analysis from healthcare and societal perspectives, respectively, with sigmoidal vaccine efficacy profiles. Panels (E) and (F) correspond to the analysis from the healthcare and societal perspectives, respectively, with constant vaccine efficacy profiles. Note: in panel B, curves for MI and L4 are superposed.

Using constant vaccine efficacy profiles, we estimated similar PPD for nirsevimab immunisations programs evaluated. From a healthcare perspective (**Table 4**), the maximum PPD for a positive NMB was $610 in L1, and reduced to $370 in L2, $295 in L3, and $215 in L4 (**Figure 3E**). Corresponding to these PPDs, the probabilities of L1, L2, L3, and L4 being cost-effective were 54%, 69%, 90%, and 86%, respectively. For MI, the maximum PPD was $185 (**Figure 3E**), with cost-effectiveness probability of 81%. From a societal perspective (**Table 4**) with constant vaccine efficacy profiles, the maximum PPD for a positive NMB was estimated to be $700 in L1, $450 in L2, $380 in L3, and $285 in L4 (**Figure 3D**). The probabilities of these programs being cost-effective at their maximum PPD were 55%, 58%, 82%, and 78% in L1, L2, L3, and L4, respectively. MI was cost-effective for a PPD up to $235, with the probability of 83%.

**Table 4.** Model estimates of cost-effectiveness analyses associated with infant and maternal immunisation programs as standalone prevention strategies from healthcare and societal perspectives at the WTP of $50,000 using constant vaccine efficacy profiles. All strategies were compared to the baseline with no intervention.

| Prevention strategy | Maximum PPD, \$ | Incremental costs, \$<br>(95% CI) | QALYs gained<br>(95% CI) | ICER, \$/QALY<br>(95% CI) | Budget impact, \$ |
| --- | --- | --- | --- | --- | --- |
| *Healthcare perspective* |  |  |  |  |  |
| L1 | 610 | 1,117<br>(-397 to 2,642) | 0.024<br>(0.018 to 0.031) | 46,135<br>(-15,407 to 123,253) | 1,144 |
| L2 | 370 | 1,303<br>(-443 to 3,025) | 0.036<br>(0.028 to 0.045) | 36,306<br>(-11,975 to 91,544) | 1,330 |
| L3 | 295 | 2,311<br>(-1,131 to 5,714) | 0.094<br>(0.082 to 0.106) | 24,716<br>(-11,905 to 63,068) | 2,354 |
| L4 | 215 | 3,017<br>(-1,327 to 7,221) | 0.110<br>(0.098 to 0.123) | 27,348<br>(-11,810 to 67,888) | 3,050 |
| MI | 185 | 3,991<br>(-45 to 8,032) | 0.117<br>(0.104 to 0.131) | 34,041<br>(-362 to 71,355) | 4,010 |
| *Societal perspective* |  |  |  |  |  |
| L1 | 700 | 1,114<br>(-507 to 2,722) | 0.024<br>(0.018 to 0.031) | 45,987<br>(-19,668 to 127,807) | 4,525 |
| L2 | 450 | 1,584<br>(-332 to 3,462) | 0.036<br>(0.028 to 0.045) | 44,162<br>(-8,803 to 105,052) | 9,637 |
| L3 | 380 | 2,751<br>(-1242 to 6,664) | 0.094<br>(0.082 to 0.106) | 29,422<br>(-13,081 to 73,612) | 51,790 |
| L4 | 285 | 3,439<br>(-1,753 to 8,664) | 0.110<br>(0.098 to 0.123) | 31,187<br>(-15,679 to 80,821) | 80,960 |
| MI | 235 | 3,554<br>(-1,100 to 8,303) | 0.117<br>(0.104 to 0.131) | 30,317<br>(-9,310 to 73,014) | 59,660 |

### Cost-effectiveness of a combined nirsevimab and RSVpreF prevention program

LMI was cost-effective for various combinations of PPD values for nirsevimab and RSVpreF (**Figure 4**). Here, we considered maximum PPDs derived for L1 and L4 programs in combination with MI at which LMI program was cost-effective (**Tables 5 and 6**). From a healthcare perspective, at PPD of $615 for nirsevimab with sigmoidal vaccine efficacy profiles, LMI was cost-effective (NMB>0) for a PPD up to $140 for RSVpreF, with probability of 100% at the WTP threshold of $50,000 per QALY gained (**Table 5**). Reducing PPD for nirsevimab to $215, LMI was cost-effective for a PPD up to $155 for RSVpreF with the probability of 96%. From a societal perspective, LMI with a PPD of $705 for nirsevimab and $180 for RSVpreF was cost-effective with the probability of 98% (**Table 5**). LMI was also cost-effective for a combination PPD of $290 and $195 for nirsevimab and RSVpreF, respectively, with the probability of 95%.

**Figure 4.**
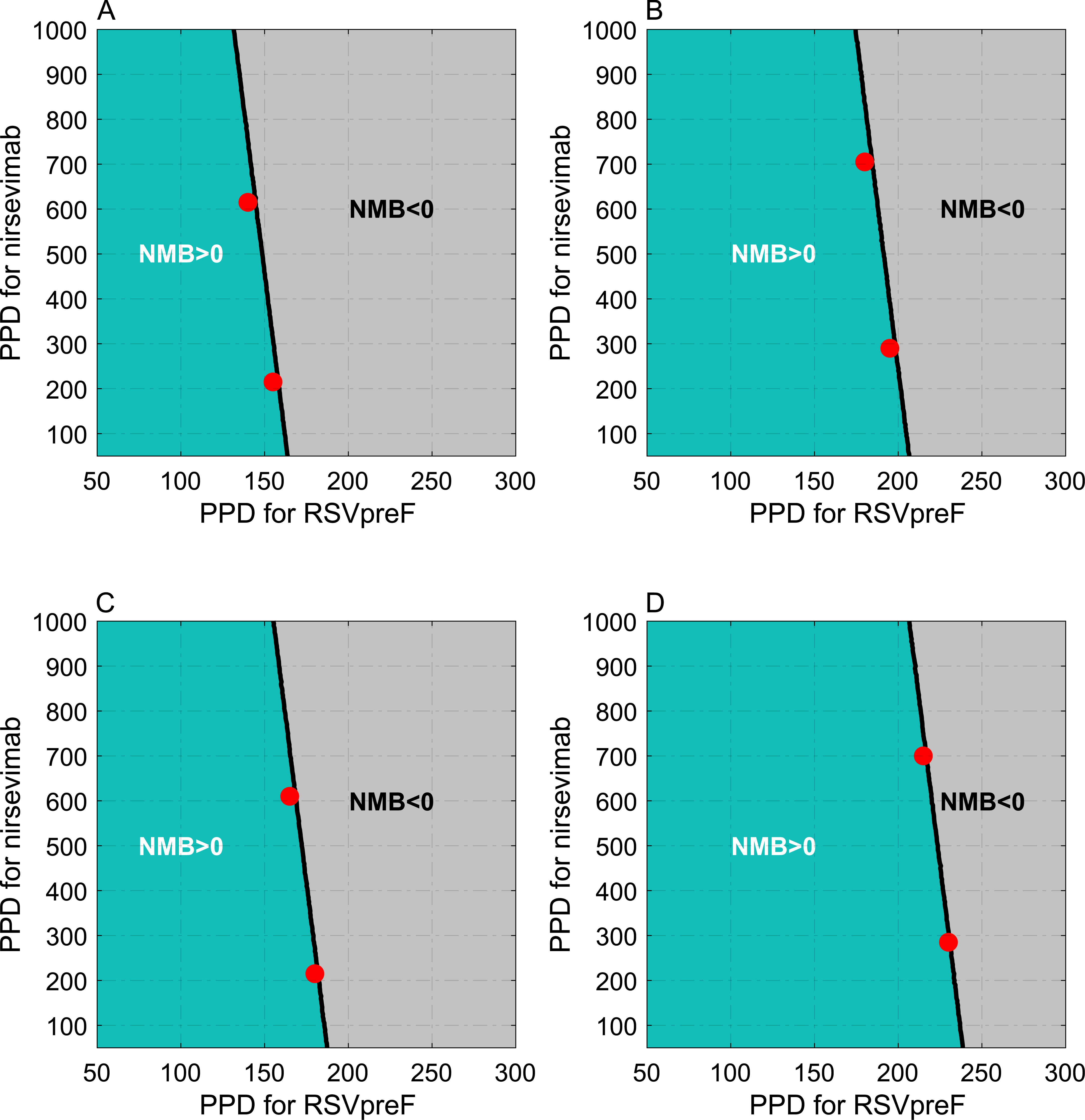
Net monetary benefit (NMB) of the combined infant and maternal immunisation program at the WTP of $50,000 per QALY gained as a function of PPD for nirsevimab and RSVpreF. Panels (A) and (B) correspond to the analysis from healthcare and societal perspectives, respectively, with sigmoidal vaccine efficacy profiles. Panels (C) and (D) correspond to the analysis from the healthcare and societal perspectives, respectively, with constant vaccine efficacy profiles. Red circles correspond to the PPD values in Tables 5 and 6.

**Table 5.** Model estimates of cost-effectiveness analyses associated with the combined infant and maternal immunisation program from healthcare and societal perspectives at the WTP of $50,000 using sigmoidal vaccine efficacy profiles. All strategies were compared to the baseline with no intervention.

| Nirsevimab<br>PPD, \$ | RSVpreF<br>PPD, \$ | Incremental costs, \$<br>(95% CI) | QALYs gained<br>(95% CI) | ICER, \$/QALY<br>(95% CI) | Budget impact per<br>100,000 population, \$ |
| --- | --- | --- | --- | --- | --- |
| *Healthcare perspective* |  |  |  |  |  |
| 615 | 140 | 432<br>(-3,496 to 4,245) | 0.112<br>(0.099 to 0.126) | 3,853<br>(-30,616 to 38,887) | 467 |
| 215 | 155 | 2,041<br>(-1,846 to 5,892) | 0.112<br>(0.099 to 0.126) | 18,193<br>(-16,004 to 54,135) | 2,135 |
| *Societal perspective* |  |  |  |  |  |
| 705 | 180 | 650<br>(-3854 to 5042) | 0.112<br>(0.099 to 0.126) | 5797<br>(-33894 to 46014) | 48,368 |
| 290 | 195 | 1,738<br>(-2,653 to 6,132) | 0.112<br>(0.099 to 0.126) | 15,511<br>(-23,306 to 56,333) | 49,473 |

**Table 6.** Model estimates of cost-effectiveness analyses associated with the combined infant and maternal immunisation program from healthcare and societal perspectives at the WTP of $50,000 using constant vaccine efficacy profiles. All strategies were compared to the baseline with no intervention.

| **Nirsevimab**<br>**PPD, \$** | **RSVpreF**<br>**PPD, \$** | **Incremental costs, \$**<br>**(95% CI)** | **QALYs gained**<br>**(95% CI)** | **ICER, \$/QALY**<br>**(95% CI)** | **Budget impact per**<br>**100,000 population, \$** |
| --- | --- | --- | --- | --- | --- |
| *Healthcare perspective* |  |  |  |  |  |
| 610 | 165 | 2,046<br>(-1,994 to 6,095) | 0.119<br>(0.106 to 0.133) | 17,243<br>(-16,725 to 53,016) | 2,099 |
| 215 | 180 | 3,911<br>(-272 to 7,989) | 0.119<br>(0.106 to 0.133) | 32,932<br>(-2,311 to 69,494) | 3,954 |
| *Societal perspective* |  |  |  |  |  |
| 700 | 215 | 3,990<br>(-697 to 8,678) | 0.119<br>(0.106 to 0.133) | 33,598<br>(-5,820 to 75,142) | 61,130 |
| 285 | 230 | 5,083<br>(266 to 9,849) | 0.119<br>(0.106 to 0.133) | 42,805<br>(2,217 to 85,356) | 62,234 |

With constant vaccine efficacy profiles, LMI was cost-effective from a healthcare perspective at PPD of $610 for nirsevimab and $165 for RSVpreF, with the probability of 0.96% at the WTP threshold of $50,000 per QALY gained (**Table 6**). Reducing PPD for nirsevimab to $215, LMI was cost-effective at a PPD of $180 for RSVpreF with the probability of 82%. From a societal perspective, LMI with constant vaccine efficacy profiles was cost-effective at a PPD of $700 for nirsevimab and $215 for RSVpreF, with the probability of 78% (**Table 6**). LMI was also cost-effective for a combination PPD of $285 and $230 for nirsevimab and RSVpreF, respectively, with the probability of 63%.

### Budget impact

The total number of nirsevimab doses per 100,000 population was 38, 104, 582, and 1113 in L1, L2, L3, and L4, respectively (**Figure 3A**). For sigmoidal vaccine efficacy profiles, the annual budget impact of these interventions to the healthcare system would be $1,225 in L1, $1,668 in L2, $3,303 in L3, and $503 in L4 at the maximum PPD estimated for each program to be cost-effective (**Table 3**). For MI, the total number of RSVpreF vaccine doses was 1113 per 100,000 population (**Figure 3A)**, resulting in an annual budget impact of $4,546 to the healthcare system. From a societal perspective, the annual budget impact was estimated at $4,606, $9,976, $52,738, and $83,978 for PPD of $705 in L1, $455 in L2, $385 in L3, and $290 in L4, respectively (**Table 3**). The annual budget impact for MI with a PPD of $200 would be $49,066.

For the combined immunisation program with sigmoidal vaccine efficacy profiles, LMI was associated with an annual budget impact of $467 per 100,000 population with PPD of $615 and $140 for nirsevimab and RSVpreF, respectively (**Table 5**). When the PPD for nirsevimab and RSVpreF changed to $215 and $155, respectively, the budget impact of LMI was estimated at $2,135. From a societal perspective, the budget impact of MLI was estimated at $48,368 with a PPD of $705 and $180 for nirsevimab and RSVpreF, respectively (**Table 5**). Changing the corresponding PPDs to $290 and $195 resulted in a similar budget impact of $49,473.

Using constant vaccine efficacy profiles, we estimated the annual budget impact of infant immunisation programs with nirsevimab to the healthcare system to be $1,144 in L1, $1,330 in L2, $2,354 in L3, and $3,050 in L4 per 100,000 population at the maximum PPD estimated for each program to be cost-effective (**Table 4**). MI resulted in a total annual budget impact of $4,010 to the healthcare system at the maximum PPD. From a societal perspective, the annual budget impact was estimated at $4,525, $9,637, $51,790, and $80,960 for PPD of $700 in L1, $450 in L2, $380 in L3, and $285 in L4, respectively (**Table 4**). The annual budget impact for MI with a PPD of $235 would be $59,660 per 100,000 population.

For the combined immunisation program with constant vaccine efficacy profiles, LMI was associated with an annual budget impact of $2,099 per 100,000 population with PPD of $610 and $165 for nirsevimab and RSVpreF, respectively (**Table 6**). When the PPD for nirsevimab and RSVpreF changed to $215 and $180, respectively, the budget impact of LMI was estimated at $3,954. From a societal perspective, the budget impact of MLI was estimated at $61,130 with a PPD of $700 for nirsevimab and $215 for RSVpreF (**Table 6**). Changing the corresponding PPDs to $285 and $230 resulted in a similar annual budget impact of $62,234.

### Secondary analyses

The results of secondary analyses for reduced coverage of nirsevimab and RSFpreF using both sigmoidal and constant vaccine profiles, without and with monetary loss of life due to RSV-related infant mortality, are provided in the Supplementary Material. We also estimated the reduction of direct healthcare costs (outpatient and inpatient care) and indirect costs (loss of productivity and out-of-pocket expenses) achieved from interventions (**Supplementary Material, Tables A4 and A5**). Our results show that PPD for cost-effective programs with nirsevimab is sensitive to the target groups among the infant population, but remained relatively robust with respect to the efficacy profiles of nirsevimab and the coverage of immunisation.

## Discussion

In this study, we evaluated the cost-effectiveness of infant and maternal immunisation programs against RSV disease using nirsevimab and RSVpreF as new preventive measures. Seasonal administration of nirsevimab to the entire birth cohort could be cost-effective at a sufficiently low PPD. However, this strategy would entail a substantial annual budget impact from a societal perspective. We found that a combined program of year-round vaccination of pregnant women with RSVpreF, followed by immunising those infants at high-risk of severe RSV disease with nirsevimab was comparable to an extended nirsevimab-only program for the entire birth cohort in reducing RSV-related mortality among infants, but required a lower annual budget impact.

Our results remained qualitatively consistent at different WTP thresholds, with the target population being an important factor in determining the range of PPD for cost-effective immunisation strategies. Previous studies have evaluated the cost-effectiveness of prevention strategies against RSV disease in infants, including long-acting monoclonal antibody, maternal vaccination, and potential active vaccination.^50,52,58–63^ These studies have been conducted in different population settings including the United States,^62^ England and Wales,^61,63^ Norway,^64^ other European countries,^59^ and low- and middle-income countries,^58,60^ indicating the potential for cost-effective immunisation programs. However, no previous work has evaluated cost-effectiveness of these interventions in Canada, except one study that is specific to Nunavik, a small population in the Canadian Arctic region, with significant burden of RSV disease.^50^ Furthermore, published studies evaluating cost-effectiveness of long-acting monoclonal antibody and maternal vaccination have relied on early efficacy estimates of these products with varying assumptions across population and epidemiological contexts. Our study provides a comprehensive cost-effectiveness analysis of these RSV preventive measures, with the most recent efficacy estimates, in a population setting reflective of the Canadian south. Moreover, we have provided a comparison between various programs using nirsevimab and RSVpreF vaccine, as well as a combined strategy for vaccination of pregnant women followed by immunisation of high-risk infants.

Published studies have employed different approaches including cohort, decision-tree, and transmission dynamic models.^52,58–63^ Our analysis is based on a discrete-event simulation model, following a birth cohort up to one year of age, without consideration of RSV transmission dynamics. Employing transmission dynamic models could allow for the evaluation of population-wide benefits of immunisation programs. However, since the effect of nirsevimab and RSVpreF in reducing RSV infection or transmission is not yet known, estimating the indirect benefits of immunisation, including herd effects, may be difficult.

### Limitations

A strength of our study is the stratification of the infant population by wGA and critical risk factors of CLD and CHD, which allowed us to utilise available estimates associated with RSV outcomes in infants. However, our model has several limitations. First, for efficacy of nirsevimab against RSV disease outcomes, we relied on reported estimates for infants of ≥29 wGA.^39^ If the efficacy among preterm infants <29 wGA is lower than those ≥29 wGA, the maximum PPD for cost-effectiveness may be lower than our estimates. Second, the efficacy of a single dose of nirsevimab may also depend on weight-based dosing.^10,11^ We assumed that PPD is not affected by the dosage. Third, the model includes only CLD and CHD as risk factors; however, other risk factors may be considered such as cystic fibrosis, Down syndrome, and immunocompromise,^21^ which were not considered in our analysis due to the lack of specific estimates. Furthermore, the National Advisory Committee on Immunisation recommends only hemodynamically significant CHD infants for use of palivizumab, as opposed to all infants with congenital heart disease.^21^ Although the proportion of CHD infants who are hemodynamically significant could be as high as 79% (95% CI: 62% to 91%),^65^ in the absence of such estimates in Canada, we considered all CHD infants in the basecase analysis and 80% of them in the secondary analysis of combined nirsevimab and RSVpreF immunisation program. Fourth, we note that maternal vaccination is recommended during the third trimester of pregnancy and therefore a proportion of preterm birth mothers may not receive RSVpreF prior to their infants’ birth, which could be considered under our secondary analysis with 60% vaccine coverage of pregnant women.

Finally, we recognize that the feasibility of different immunisation programs to deliver interventions to pregnant women and infants seasonally are not considered here, and will impact decision making.

## Conclusion

Our study shows that prevention strategies against RSV disease in infants using nirsevimab and RSVpreF could be cost-effective. Passive immunisation of all infants experiencing their first RSV season would require a PPD under $290 to become cost-effective without considering the monetary loss of life due to RSV-related infant mortality. However, this program would incur a higher budget impact to the healthcare system than a cost-effective strategy that combines year-round maternal vaccination with seasonal administration of nirsevimab to high-risk infants who are currently eligible for palivizumab.

## Data Availability

The computational model is available at https://github.com/affans/rsv_costeffectiveness

## Contributors

Seyed Moghadas and Joanne Langley conceived the study; Seyed Moghadas designed the model framework; Seyed Moghadas and Elaheh Abdollahi collected input parameters; Affan Shoukat developed the computational model and performed simulations; Seyed Moghadas analysed the simulated data and wrote the first draft of the manuscript; Scott Halperin and Alison Galvani provided insights into the analysis and interpretation of the results; all authors contributed to the writing.

## Data sharing agreement

The computational model is available at https://github.com/affans/rsv_costeffectiveness

## References

1. Shi, T. et al. Global, regional, and national disease burden estimates of acute lower respiratory infections due to respiratory syncytial virus in young children in 2015: a systematic review and modelling study. The Lancet 390, 946–958 (2017).

2. Borchers, A. T., Chang, C., Gershwin, M. E. & Gershwin, L. J. Respiratory Syncytial Virus— A Comprehensive Review. Clin. Rev. Allergy Immunol. 45, 331–379 (2013).

3. Li, Y. et al. Global, regional, and national disease burden estimates of acute lower respiratory infections due to respiratory syncytial virus in children younger than 5 years in 2019: a systematic analysis. Lancet Lond. Engl. 399, 2047–2064 (2022).

4. Thampi, N. et al. Health care costs of hospitalization of young children for respiratory syncytial virus infections: a population-based matched cohort study. CMAJ Open 9, E948–E956 (2021).

5. Mitchell, I., Defoy, I. & Grubb, E. Burden of Respiratory Syncytial Virus Hospitalizations in Canada. Can. Respir. J. 2017, 1–9 (2017).

6. McLaurin, K. K., Farr, A. M., Wade, S. W., Diakun, D. R. & Stewart, D. L. Respiratory syncytial virus hospitalization outcomes and costs of full-term and preterm infants. J. Perinatol. 36, 990–996 (2016).

7. Rafferty, E. et al. Evaluating the Individual Healthcare Costs and Burden of Disease Associated with RSV Across Age Groups. PharmacoEconomics 40, 633–645 (2022).

8. National Advisory Committee on Immunization (NACI), Moore, D., Sinilaite, A. & Killikelly, A. Summary of the National Advisory Committee on Immunization (NACI) statement update on the recommended use of palivizumab to reduce complications of respiratory syncytial virus infection in infants. Can. Commun. Dis. Rep. 48, 363–366 (2022).

9. Graham, B. S. The Journey to RSV Vaccines — Heralding an Era of Structure-Based Design. N. Engl. J. Med. 388, 579–581 (2023).

10. Griffin, M. P. et al. Single-Dose Nirsevimab for Prevention of RSV in Preterm Infants. N. Engl. J. Med. 383, 415–425 (2020).

11. Hammitt, L. L. et al. Nirsevimab for Prevention of RSV in Healthy Late-Preterm and Term Infants. N. Engl. J. Med. 386, 837–846 (2022).

12. Pfizer. Pfizer Announces Positive Top-Line Data of Phase 3 Global Maternal Immunization Trial for its Bivalent Respiratory Syncytial Virus (RSV) Vaccine Candidate. https://www.pfizer.com/news/press-release/press-release-detail/pfizer-announces-positive-top-line-data-phase-3-global (2022).

13. Statistics Canada. Census Profile. 2021 Census of Population. (2023).

14. Johnston, K. M. et al. The economic burden of prematurity in Canada. BMC Pediatr. 14, 93 (2014).

15. Born Ontario. Number of Live Births and Stillbirths in Ontario, by month: March 2021 - February 2022. https://www.bornontario.ca/en/news/stillbirths-in-ontario-recent-data.aspx.

16. Statistics Canada. Live births, by month. doi:10.25318/1310041501-ENG.

17. Isayama, T. et al. Comparison of Mortality and Morbidity of Very Low Birth Weight Infants Between Canada and Japan. Pediatrics 130, e957–e965 (2012).

18. Liu, S. et al. Effect of Folic Acid Food Fortification in Canada on Congenital Heart Disease Subtypes. Circulation 134, 647–655 (2016).

19. Zheng, Z., Pitzer, V. E., Shapiro, E. D., Bont, L. J. & Weinberger, D. M. Estimation of the Timing and Intensity of Reemergence of Respiratory Syncytial Virus Following the COVID-19 Pandemic in the US. JAMA Netw. Open 4, e2141779 (2021).

20. Nduaguba, S. O., Tran, P. T., Choi, Y. & Winterstein, A. G. Respiratory syncytial virus reinfections among infants and young children in the United States, 2011–2019. PLOS ONE 18, e0281555 (2023).

21. National Advisory Committee on Immunization. Recommended use of palivizumab to reduce complications of respiratory syncytial virus infection in infants. https://www.canada.ca/en/public-health/services/publications/vaccines-immunization/palivizumab-respiratory-syncitial-virus-infection-infants.html#a5.5 (2022).

22. Ratti, C., Greca, A. D., Bertoncelli, D., Rubini, M. & Tchana, B. Prophylaxis protects infants with congenital heart disease from severe forms of RSV infection: an Italian observational retrospective study: Palivizumab prophylaxis in children with congenital heart disease. Ital. J. Pediatr. 49, 4 (2023).

23. Paes, B. et al. Defining the Risk and Associated Morbidity and Mortality of Severe Respiratory Syncytial Virus Infection Among Infants with Chronic Lung Disease. Infect. Dis. Ther. 5, 453–471 (2016).

24. Lanctôt, K. L. et al. The cost-effectiveness of palivizumab for respiratory syncytial virus prophylaxis in premature infants with a gestational age of 32–35 weeks: a Canadian-based analysis. Curr. Med. Res. Opin. 24, 3223–3237 (2008).

25. Anderson, E. et al. SENTINEL1: An Observational Study of Respiratory Syncytial Virus Hospitalizations among U.S. Infants Born at 29 to 35 Weeks’ Gestational Age Not Receiving Immunoprophylaxis. Am. J. Perinatol. 34, 51–61 (2016).

26. Bozaykut. Evaluation of Risk Factors for Recurrent Wheezing Episodes. J. Clin. Med. Res. (2013) doi:10.4021/jocmr1543w.

27. Schauer, U. et al. RSV bronchiolitis and risk of wheeze and allergic sensitisation in the first year of life. Eur. Respir. J. 20, 1277–1283 (2002).

28. Zhou, Y. et al. Recurrent Wheezing and Asthma After Respiratory Syncytial Virus Bronchiolitis. Front. Pediatr. 9, 649003 (2021).

29. Welliver, R. C. et al. Fatality rates in published reports of RSV hospitalizations among high-risk and otherwise healthy children. Curr. Med. Res. Opin. 26, 2175–2181 (2010).

30. Checchia, P. A. et al. Mortality and morbidity among infants at high risk for severe respiratory syncytial virus infection receiving prophylaxis with palivizumab: A systematic literature review and meta-analysis: *Pediatr*. Crit. Care Med. 12, 580–588 (2011).

31. Hall, C. B. et al. Respiratory Syncytial Virus–Associated Hospitalizations Among Children Less Than 24 Months of Age. Pediatrics 132, e341–e348 (2013).

32. Sampalis, J. S. Morbidity and mortality after RSV-associated hospitalizations among premature Canadian infants. J. Pediatr. 143, 150–156 (2003).

33. Szabo, S. M. et al. The risk of mortality among young children hospitalized for severe respiratory syncytial virus infection. Paediatr. Respir. Rev. 13, S1–S8 (2013).

34. Tam, J. et al. Pediatric Investigators Collaborative Network on Infections in Canada Study of Respiratory Syncytial Virus–associated Deaths in Pediatric Patients in Canada, 2003–2013. Clin. Infect. Dis. 68, 113–119 (2019).

35. Wingert, A. et al. Burden of illness in infants and young children hospitalized for respiratory syncytial virus: A rapid review. Can. Commun. Dis. Rep. 47, 381–396 (2021).

36. Statistics Canada. Gross domestic product (GDP) at basic prices, by industry, provinces and territories. doi:10.25318/3610040201-ENG.

37. CADTH. Guidelines for the economic evaluation of health technologies: Canada. 4th ed. Ottawa https://www.cadth.ca/sites/default/files/pdf/guidelines_for_the_economic_evaluation_of_health_technologies_canada_4th_ed.pdf (2017).

38. Government of Canada. Results of the Survey on Vaccination during Pregnancy 2021. https://www.canada.ca/en/public-health/services/publications/vaccines-immunization/survey-vaccination-during-pregnancy-2021.html (2022).

39. Simões, E. A. F. et al. Efficacy of nirsevimab against respiratory syncytial virus lower respiratory tract infections in preterm and term infants, and pharmacokinetic extrapolation to infants with congenital heart disease and chronic lung disease: a pooled analysis of randomised controlled trials. Lancet Child Adolesc. Health 7, 180–189 (2023).

40. Ortega-Sanchez, I. R. Economics of Preventing Respiratory Syncytial Virus Lower Respiratory Tract Infections (RSV-LRTI) among US Infants with Nirsevimab. (2023).

41. Kampmann, B. et al. Bivalent Prefusion F Vaccine in Pregnancy to Prevent RSV Illness in Infants. N. Engl. J. Med. 388, 1451–1464 (2023).

42. Munjal, I. Safety and Efficacy of RSV Bivalent PreF Maternal Vaccine. (2023).

43. Fleming-Dutra, K. Evidence to Recommendations Framework: Pfizer Maternal RSVpreF Vaccine. (2023).

44. Papenburg, J. et al. Cost-analysis of Withdrawing Immunoprophylaxis for Respiratory Syncytial Virus in Infants Born at 33–35 Weeks Gestational Age in Quebec. Pediatr. Infect. Dis. J. 39, 694–699 (2020).

45. McGirr, A. A., Schwartz, K. L., Allen, U., Solomon, M. & Sander, B. The cost-effectiveness of palivizumab in infants with cystic fibrosis in the Canadian setting: A decision analysis model. Hum. Vaccines Immunother. 13, 599–606 (2017).

46. Stinnett, A. A. & Mullahy, J. Net Health Benefits: A New Framework for the Analysis of Uncertainty in Cost-Effectiveness Analysis. Med. Decis. Making 18, S68–S80 (1998).

47. Pichon-Riviere, A., Drummond, M., Palacios, A., Garcia-Marti, S. & Augustovski, F. Determining the efficiency path to universal health coverage: cost-effectiveness thresholds for 174 countries based on growth in life expectancy and health expenditures. Lancet Glob. Health 11, e833–e842 (2023).

48. Ochalek, J., Lomas, J. & Claxton, K. Assessing health opportunity costs for the Canadian health care systems. 2018. http://www.pmprb-cepmb.gc.ca/CMFiles/Consultations/new_guidelines/Canada_report_2018-03-14_Final.pdf (2018).

49. GDP per capita (current US$) - Canada. World Bank Open Data https://data.worldbank.org.

50. Nourbakhsh, S. et al. Effectiveness and cost-effectiveness of RSV infant and maternal immunization programs: A case study of Nunavik, Canada. eClinicalMedicine 41, 101141 (2021).

51. Tam, D. Y. et al. The cost effectiveness of palivizumab in term Inuit infants in the Eastern Canadian Arctic. J. Med. Econ. 12, 361–370 (2009).

52. Pouwels, K. B. et al. Potential Cost-Effectiveness of RSV Vaccination of Infants and Pregnant Women in Turkey: An Illustration Based on Bursa Data. PLOS ONE 11, e0163567 (2016).

53. Roy, L. M. Deriving health utility weights for infants with Respiratory Syncytial Virus (RSV). (British Columbia, 2014).

54. Weiner, L. B., Masaquel, A. S., Polak, M. J. & Mahadevia, P. J. Cost-effectiveness analysis of palivizumab among pre-term infant populations covered by Medicaid in the United States. J. Med. Econ. 15, 997–1018 (2012).

55. Greenough, A. Health care utilisation of prematurely born, preschool children related to hospitalisation for RSV infection. Arch. Dis. Child. 89, 673–678 (2004).

56. Hodgson, D. et al. Estimates for quality of life loss due to Respiratory Syncytial Virus. Influenza Other Respir. Viruses 14, 19–27 (2020).

57. Husereau, D. et al. Correction to: Consolidated Health Economic Evaluation Reporting Standards 2022 (CHEERS 2022) Statement: Updated Reporting Guidance for Health Economic Evaluations. Appl. Health Econ. Health Policy. 20, 781–782 (2022).

58. Laufer, R. S. et al. Cost-effectiveness of infant respiratory syncytial virus preventive interventions in Mali: A modeling study to inform policy and investment decisions. Vaccine 39, 5037–5045 (2021).

59. Getaneh, A. M. et al. Cost-effectiveness of monoclonal antibody and maternal immunization against respiratory syncytial virus (RSV) in infants: Evaluation for six European countries. Vaccine 41, 1623–1631 (2023).

60. Koltai, M. et al. Estimating the cost-effectiveness of maternal vaccination and monoclonal antibodies for respiratory syncytial virus in Kenya and South Africa. BMC Med. 21, 120 (2023).

61. Hodgson, D., Pebody, R., Panovska-Griffiths, J., Baguelin, M. & Atkins, K. E. Evaluating the next generation of RSV intervention strategies: a mathematical modelling study and cost-effectiveness analysis. BMC Med. 18, 348 (2020).

62. Kieffer, A. et al. Expected Impact of Universal Immunization With Nirsevimab Against RSV-Related Outcomes and Costs Among All US Infants in Their First RSV Season: A Static Model. J. Infect. Dis. 226, S282–S292 (2022).

63. Hodgson, D. et al. Optimal Respiratory Syncytial Virus intervention programmes using Nirsevimab in England and Wales. Vaccine 40, 7151–7157 (2022).

64. Li, X. et al. Cost-effectiveness of Respiratory Syncytial Virus Disease Prevention Strategies: Maternal Vaccine Versus Seasonal or Year-Round Monoclonal Antibody Program in Norwegian Children. J. Infect. Dis. 226, S95–S101 (2022).

65. Bergman, G., Hærskjold, A., Stensballe, L. G., Kieler, H. & Linder, M. Children with hemodynamically significant congenital heart disease can be identified through population-based registers. Clin. Epidemiol. 7, 119–127 (2015).

66. Lanari, M. et al. Burden of respiratory syncytial virus hospitalisation among infants born at 32–35 weeks’ gestational age in the Northern Hemisphere: pooled analysis of seven studies. Epidemiol. Infect. 148, e170 (2020).

67. Canadian Institute for Health Information. Hospital spending. https://www.cihi.ca/sites/default/files/document/hospital-spending-highlights-2020-en.pdf (2020).

68. CIHI: Canadian Institute for Health Information. Care in Canadian ICUs. 1–36 (2016).

