## Supplementary Material for "Cost-Effectiveness Analysis of Nirsevimab and RSVpreF Vaccine Prevention Strategies for Respiratory Syncytial Virus Disease in infants: A Canadian Immunisation Research Network (CIRN) Study"

This Supplemental provides additional information for parameterisation of the model, additional figures supporting results reported in the main text, and the results of secondary analyses with willingness-to-pay thresholds of $30,000 and $70,000.

#### 1. Model scenarios and inputs

Scenarios evaluated in the model are summarised in Table A1 below:

**Table A1. Immunisation programs, target populations, and immunisation coverages.**

| Immunisation Program | Target population | Immunisation coverage | |
|---|---|---|---|
| | | Basecase analysis | Secondary analysis |
| L1 | Infants ≤32 wGA, and infants with CLD or CHD | 100% | 80% |
| L2 | Infants ≤36 wGA, and infants with CLD or CHD | 100% | 80% |
| L3 | Infants ≤36 wGA, and infants with CLD or CHD, and infants ≥37 wGA born during the RSV season | 100% | 80% |

| | | | |
|---|---|---|---|
| L3 | Entire birth cohort | 100% | 80% |
| MI | Pregnant women | 100% | 60% |
| LMI | Infants ≤32 wGA, and infants with CLD or CHD | 100% | 80% |
| | pregnant women | 100% | 60% |

The birth cohort included 1113 infants with approximately 93 births per month, of which ~2% were born at less than 33 wGA and ~6% were preterm of 33-35 wGA. The population size of infants targeted in L1, L2, L3, and L4 were 38, 104, 582, and 1113 per 100,000 population.

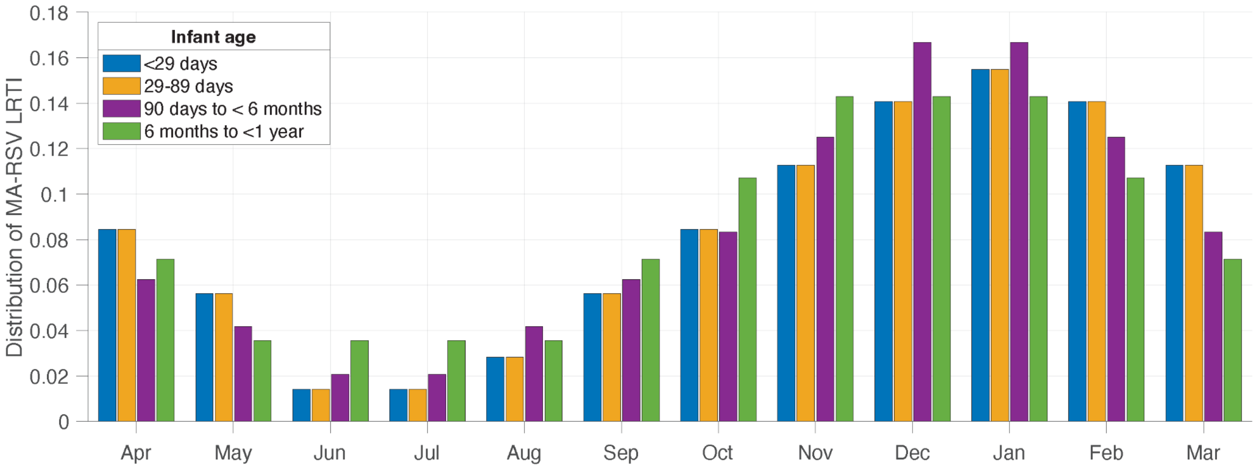

**Figure A1.** Seasonal distribution of MA-RSV LRTI for infants under 1 year of age. To generate the distribution, we considered the estimated medically attended RSV infection, on a monthly basis, for 9 seasons from 2010-11 to 2018-2019.[1] Applying the rates of MA RSV reported for infants under 1 year of age (Table A2), monthly distribution of MA RSV LRTI for different age groups among infants was generated by averaging each month over the 9 seasons.

**Table A2.** Rates of MA RSV LRTI among infants under 1 year of age.[1] Rates were derived from previously published study, estimating the rate of MA RSV among all age groups in the population (Table 1 in Ref. 1).

| **Age** | **Rate** |
|---|---|
| < 29 days | 5.1% |
| 29–89 days | 14.7% |

| | |
|---|---|
| 90 days to < 6 months | 11.12% |
| 6 months to < 1 year | 11.9% |

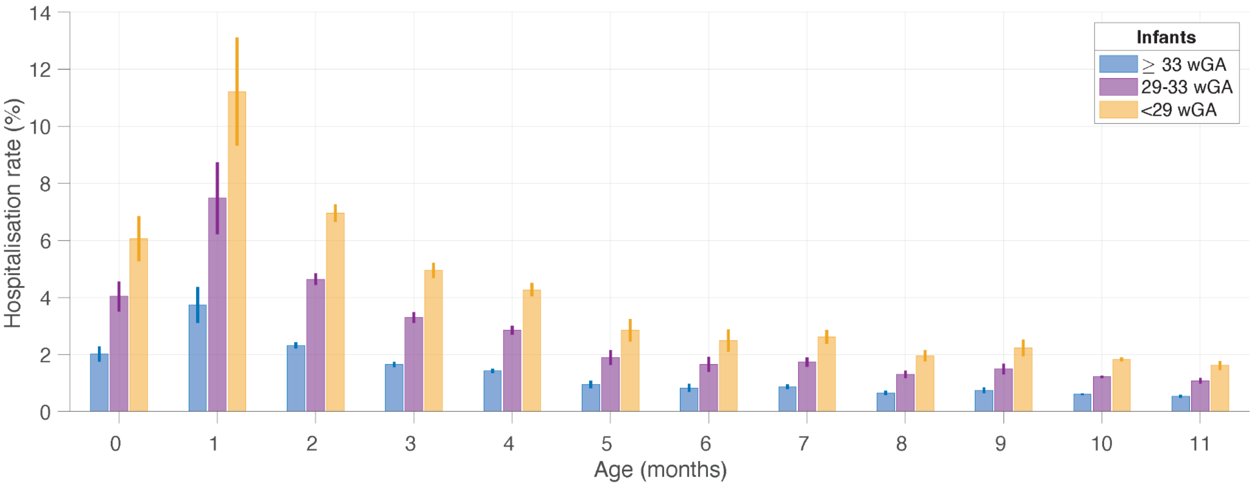

**Figure A2.** Hospitalisation rates of MA RSV LRT infants based on wGA and age at incidence derived from previously published studies.[2,3] The range of rates are reported in Table A3.

**Table A3.** Hospitalisation rates (%) by wGA and age at incidence.

| **wGA** | **Age at incidence (months)** | | | | | | | | | | | |
|---|---|---|---|---|---|---|---|---|---|---|---|---|
| **≥33** | **0** | **1** | **2** | **3** | **4** | **5** | **6** | **7** | **8** | **9** | **10** | **11** |
| Low | 1.76 | 3.11 | 2.22 | 1.56 | 1.35 | 0.82 | 0.70 | 0.79 | 0.59 | 0.65 | 0.59 | 0.49 |
| High | 2.28 | 4.37 | 2.42 | 1.74 | 1.50 | 1.08 | 0.96 | 0.95 | 0.72 | 0.84 | 0.63 | 0.59 |
| mean | 2.02 | 3.74 | 2.32 | 1.65 | 1.425 | 0.95 | 0.83 | 0.87 | 0.655 | 0.745 | 0.61 | 0.54 |
| **29-32** | | | | | | | | | | | | |
| Low | 3.52 | 6.22 | 4.44 | 3.12 | 2.70 | 1.64 | 1.40 | 1.58 | 1.18 | 1.30 | 1.18 | 0.98 |
| High | 4.56 | 8.74 | 4.84 | 3.48 | 3.00 | 2.16 | 1.92 | 1.90 | 1.44 | 1.68 | 1.26 | 1.18 |
| mean | 4.04 | 7.48 | 4.64 | 3.3 | 2.85 | 1.9 | 1.66 | 1.74 | 1.31 | 1.49 | 1.22 | 1.08 |
| **<29** | | | | | | | | | | | | |
| Low | 5.28 | 9.33 | 6.66 | 4.68 | 4.05 | 2.46 | 2.10 | 2.37 | 1.77 | 1.95 | 1.77 | 1.47 |

| | | | | | | | | | | | | |
|---|---|---|---|---|---|---|---|---|---|---|---|---|
| High | 6.84 | 13.11 | 7.26 | 5.22 | 4.50 | 3.24 | 2.88 | 2.85 | 2.16 | 2.52 | 1.89 | 1.77 |
| mean | 6.06 | 11.22 | 6.96 | 4.95 | 4.275 | 2.85 | 2.49 | 2.61 | 1.965 | 2.235 | 1.83 | 1.62 |

The probability of hospitalisation ($p_c$) for infants with CLD or CHD condition was determined by

$$p_c(a) = \frac{p_0(a)L}{1 + p_0(a)L - p_0(a)},$$

where $p_0(a)$ is the probability of hospitalisation for infants without CLD or CHD at age $a$ in months (Table A3), and $L$ is the likelihood of hospitalisation for infants with CLD or CHD compared to those without these conditions.[4,5]

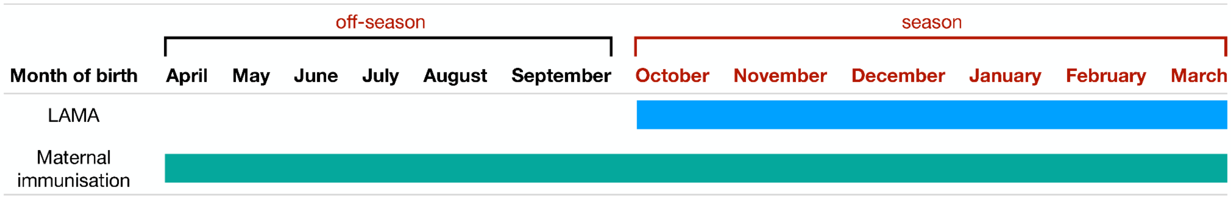

**Figure A3.** Timelines of immunisation. Nirsevimab is offered at birth to infants born during RSV season from the start of October to the end of March. Those who are born off-season are immunised with nirsevimab at the start of season in October. Maternal immunisation is offered to pregnant women year-round, during the third trimester before gestation week 33.

#### 2. Temporal decline of efficacy

To parameterize the model with temporal efficacy of nirsevimab and RSVpreF, we considered a sigmoidal decay function over a 10-month period, given by

$$V_e(t) = \mathrm{VE}_{\mathrm{max}} - \frac{(\mathrm{VE}_{\mathrm{max}} - \mathrm{VE}_{\mathrm{min}})a}{b + e^{-ct}}$$

where $\mathrm{VE}_{\mathrm{mean}}$ is the mean efficacy estimated during the follow-up period post vaccination, $\mathrm{VE}_{\mathrm{max}}$ and $\mathrm{VE}_{\mathrm{min}}$ are the maximum and minimum efficacy estimates during the study period. Assuming that the vaccine efficacy reduced to zero at 10 months after vaccination,[6] we estimated the parameters $a > 0$, $b > 0$, and $c > 0$ (using curve fitting function in Matlab) to derive estimates with the same average residual protections as reported in clinical trials for the first 5 months post immunisation. Figure A4 (A,B) illustrates the decline of protection efficacy over a 10-month period post-dose for different outcomes.

We also considered constant vaccine efficacy profiles for each outcome with the mean estimates reported in clinical trials over the study period, followed by a linear decline to a zero protection at 10 months after immunisation (Figure A4, C-D).

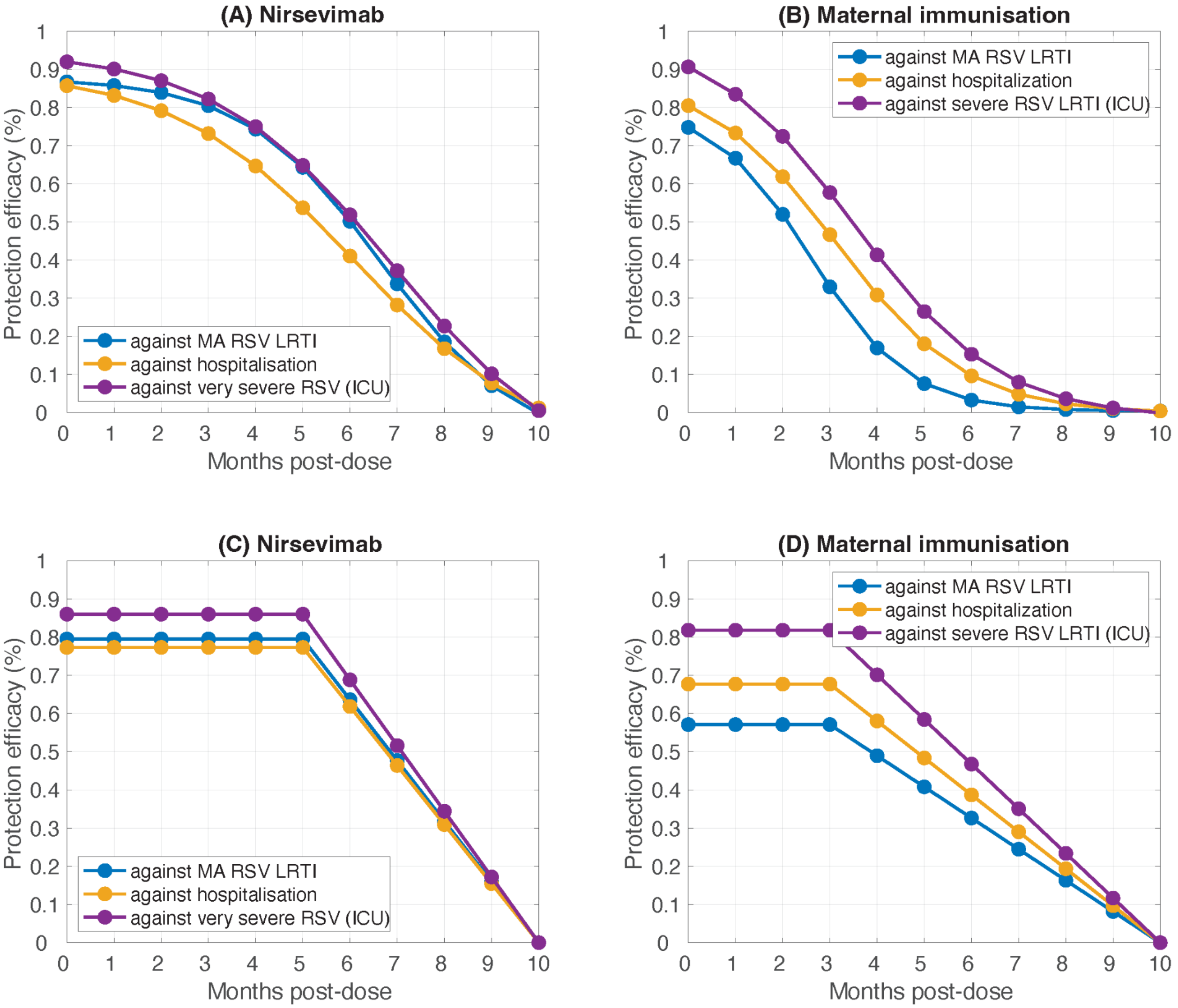

**Figure A4.** Vaccine efficacy profiles. Panel (A) and (B) correspond to sigmoidal decay of protection for each outcome after a single dose of nirsevimab and RSVpreF, respectively. Panel (C) and (D) correspond to constant protection efficacy followed by a linear decline to zero after a single dose of nirsevimab and RSVpreF, respectively.

#### 3. Reduction of direct and indirect costs

**Table A4.** Reduction of direct (outpatient and inpatient) costs and indirect costs using intervention scenarios evaluated with 100% coverage of nirsevimab and 100% coverage of maternal vaccination, compared to no intervention.

| | Reduction of costs (%) | | |
|---|---|---|---|
| **Intervention** | **Outpatient (95% CI)** | **Inpatient (95% CI)** | **indirect (95% CI)** |
| *Sigmoidal vaccine efficacy profiles* | | | |
| L1 | 2.40<br>(2.36 to 2.44) | 11.24<br>(10.36 to 12.11) | 2.80<br>(2.73 to 2.87) |
| L2 | 6.31<br>(6.25 to 6.38) | 16.86<br>(15.96 to 17.74) | 6.73<br>(6.64 to 6.83) |
| L3 | 41.30<br>(41.20 to 41.42) | 63.21<br>(62.17 to 64.25) | 41.29<br>(41.18 to 41.42) |
| L4 | 64.73<br>(64.58 to 64.90) | 79.93<br>(79.04 to 80.78) | 64.99<br>(64.82 to 65.15) |
| MI | 38.13<br>(37.98 to 38.28) | 76.13<br>(75.23 to 76.95) | 38.05<br>(37.89 to 38.21) |
| LMI | 39.26<br>(39.11 to 39.41) | 78.11<br>(77.28 to 78.91) | 39.28<br>(39.13 to 39.45) |
| *Constant vaccine efficacy profiles* | | | |
| L1 | 2.37<br>(2.33 to 2.41) | 11.21<br>(10.30 to 12.03) | 2.77<br>(2.70 to 2.84) |
| L2 | 6.21<br>(6.14 to 6.28) | 16.87<br>(15.90 to 17.75) | 6.63<br>(6.54 to 6.72) |
| L3 | 40.52<br>(40.42 to 40.63) | 63.12<br>(62.07 to 64.12) | 40.55<br>(40.44 to 40.69) |
| L4 | 63.66<br>(63.53 to 63.80) | 79.83<br>(78.91 to 80.64) | 63.97<br>(63.82 to 64.12) |
| MI | 45.93<br>(45.78 to 46.09) | 84.24<br>(83.47 to 84.88) | 46.23<br>(46.07 to 46.39) |
| LMI | 46.75<br>(46.61 to 46.91) | 85.10<br>(84.38 to 85.75) | 47.09<br>(46.94 to 47.26) |

**Table A5.** Reduction of direct (outpatient and inpatient) costs and indirect costs using intervention scenarios evaluated with 80% coverage of nirsevimab and 60% coverage of maternal vaccination, compared to no intervention.

| Intervention | Reduction of costs (%) | | |
|---|---|---|---|
| | **Outpatient (95% CI)** | **Inpatient (95% CI)** | **indirect (95% CI)** |
| *Sigmoidal vaccine efficacy profiles* | | | |
| L1 | 1.92 (1.88 to 1.95) | 9.01 (8.19 to 9.72) | 2.25 (2.18 to 2.31) |
| L2 | 5.06 (5.00 to 5.12) | 13.70 (12.83 to 14.57) | 5.42 (5.33 to 5.50) |
| L3 | 33.03 (32.92 to 33.14) | 50.63 (49.61 to 51.73) | 33.03 (32.90 to 33.14) |
| L4 | 51.80 (51.64 to 51.94) | 63.80 (62.76 to 64.87) | 52.00 (51.84 to 52.15) |
| MI | 22.83 (22.70 to 22.95) | 45.43 (44.33 to 46.54) | 22.83 (22.70 to 22.98) |
| LMI | 24.15 (24.02 to 24.27) | 49.90 (48.83 to 50.96) | 24.32 (24.18 to 24.45) |
| *Constant vaccine efficacy profiles* | | | |
| L1 | 1.89 (1.85 to 1.93) | 8.97 (8.18 to 9.71) | 2.22 (2.15 to 2.28) |
| L2 | 4.98 (4.92 to 5.04) | 13.64 (12.78 to 14.50) | 5.34 (5.26 to 5.43) |
| L3 | 32.41 (32.30 to 32.52) | 50.51 (49.45 to 51.58) | 32.43 (32.31 to 32.54) |
| L4 | 50.94 (50.79 to 51.07) | 63.72 (62.66 to 64.72) | 51.18 (51.03 to 51.32) |
| MI | 27.51 (27.38 to 27.65) | 50.02 (48.93 to 51.07) | 27.73 (27.58 to 27.88) |
| LMI | 28.67 (28.55 to 28.81) | 54.07 (52.96 to 55.10) | 29.04 (28.89 to 29.18) |

#### 4. Secondary analyses with a WTP of $50,000 per QALY gained

##### 4.1. 100% coverage of nirsevimab and 100% coverage of maternal vaccination

**Table A6.** Reduction of RSV-related outcomes (%) in intervention programs compared to no intervention with 100% coverage of nirsevimab and 100% coverage of maternal vaccination, corresponding to Figure 2 of the main text.

| Intervention | Outpatient (95% CI) | Inpatient (95% CI) | Death (95% CI) |
|---|---|---|---|
| *Sigmoidal vaccine efficacy profiles* | | | |
| L1 | 2.0 (2.0 to 2.1) | 6.2 (5.8 to 6.6) | 24.3 (16.0 to 33.2) |
| L2 | 5.9 (5.8 to 5.9) | 11.1 (10.6 11.6) | 36.3 (26.8 to 46.5) |
| L3 | 38.9 (38.8 to 39.0) | 61.2 (60.4 to 62.1) | 67.9 (58.8 to 77.3) |
| L4 | 63.4 (63.2 to 63.5) | 79.3 (78.7 to 80.1) | 77.8 (69.6 to 82.3) |
| MI | 34.0 (33.9 to 34.2) | 72.8 (72.1 to 73.5) | 72.4 (62.5 to 81.9) |
| LMI | 35.2 (35.0 to 35.3) | 74.2 (73.5 to 74.9) | 76.8 (67.1 to 85.8) |
| *Constant vaccine efficacy profiles* | | | |
| L1 | 2.0 (2.0 to 2.0) | 6.1 (5.7 to 6.6) | 24.3 (16.0 to 33.2) |
| L2 | 5.8 (5.7 to 5.8) | 11.0 (10.5 to 11.6) | 36.3 (26.8 to 46.5) |
| L3 | 38.1 (38.0 to 38.2) | 60.8 (60.0 to 61.7) | 67.9 (58.4 to 77.4) |
| L4 | 62.3 (62.1 to 62.4) | 78.9 (78.2 to 79.6) | 77.9 (69.3 to 85.4) |
| MI | 42.3 (42.1 to 42.4) | 80.6 (80.0 to 81.2) | 80.1 (71.4 to 88.9) |
| LMI | 43.1 (43.0 to 43.3) | 81.3 (80.6 to 81.9) | 82.3 (74.0 to 90.2) |

The results in this section include the monetary loss of life as an indirect cost in the cost-effectiveness analysis from a societal perspective.

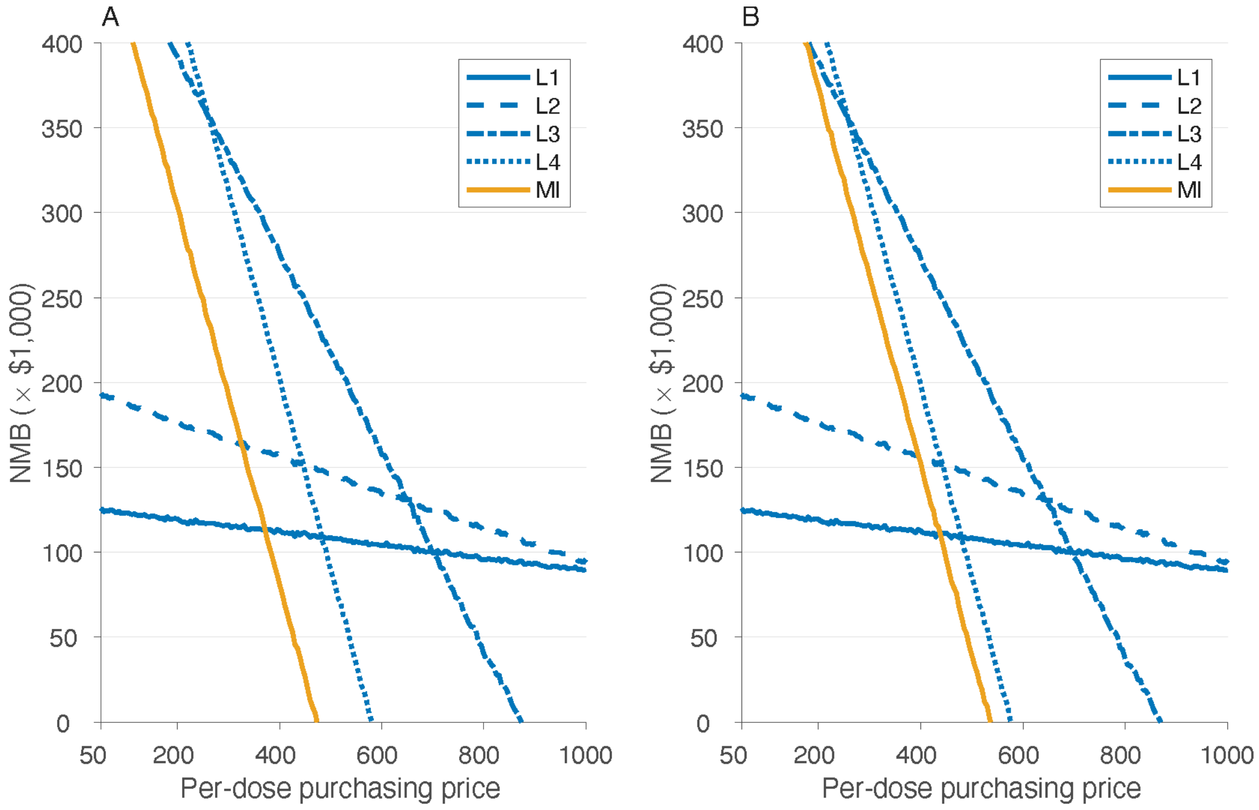

**Figure A5.** Estimated net monetary benefit (NMB) as a function of price per dose at the WTP threshold of $50,000 per QALY gained with the inclusion of monetary loss of life due to RSV-related infant mortality. Panels (A) and (B) correspond to the analysis from a societal perspective using sigmoidal and constant vaccine efficacy profiles, respectively. Note that the inclusion of monetary loss due to infant mortality (as an indirect cost) does not affect cost-effectiveness analysis from a healthcare perspective.

**Table A7.** Model estimates of cost-effectiveness analyses associated with infant and maternal immunisation programs as standalone prevention strategies from a societal perspective at the WTP of $50,000 per QALY gained. All strategies were compared to the baseline with no intervention, and with the inclusion of monetary loss of life due to RSV-related infant mortality. Since the monetary loss of life, as an indirect cost, does not affect cost-effectiveness from a healthcare perspective, the results of Table 3 and 4 of the main text are applicable for the healthcare perspective.

| Prevention strategy | Maximum PPD, $ | Incremental costs, $ (95% CI) | QALYs gained (95% CI) | ICER, $/QALY (95% CI) | Probability of being cost-effective | Budget impact per 100,000 population, $ |
|---|---|---|---|---|---|---|
| *Societal perspective, sigmoidal vaccine efficacy profiles* | | | | | | |
| L1 | 1,000 | -38,282 (-60,190 to -17,798) | 1.023 (0.607 to 1.479) | -37,439 (-41,487 to-28,904) | Cost-saving | 15,688 |
| L2 | 1,000 | -17,393 (-44,625 to 7,446) | 1.533 (1.025 to 2.083) | -11,345 (-21,523 to 7,164) | 100% | 66,571 |
| L3 | 870 | 142,571 (107,290 to 176,328) | 2.904 (2.256 to 3.595) | 49,091 (29,780 to 78,778) | 52% | 334,811 |
| L4 | 580 | 165,298 (126,772 to 201,367) | 3.324 (2.635 to 4.065) | 49,721 (31,088 to 76,585) | 50% | 406,748 |
| MI | 470 | 151,898 (113,226 to 188,026) | 3.101 (2.406 to 3.882) | 48,980 (29,229 to 78,462) | 52% | 349,576 |
| *Societal perspective, constant vaccine efficacy profiles* | | | | | | |
| L1 | 1,000 | -38,129 (-60,033 to -17,630) | 1.022 (0.607 to 1.479) | -37,291 (-41,376 to -28,707) | Cost-saving | 15,795 |
| L2 | 1,000 | -17,089 (-44,330 to 7,796) | 1.533 (1.025 to 2.083) | -11,147 (-21,381 to 7,553) | 100% | 66,752 |

| | | | | | | |
|---|---|---|---|---|---|---|
| L3 | 865 | 143,040<br>(107,283 to 176,698) | 2.895<br>(2.212 to 3.595) | 49,406<br>(29,797 to 79,337) | 52% | 333,862 |
| L4 | 575 | 163,397<br>(125,353 to 198,966) | 3.327<br>(2.635 to 4.064) | 49,118<br>(30,763 to 75,788) | 53% | 403,730 |
| MI | 535 | 170,325<br>(129,747 to 208,487) | 3.419<br>(2.684 to 4.209) | 49,813<br>(30,798 to 78,090) | 49% | 321,215 |

**Table A8.** Model estimates of cost-effectiveness analyses associated with the combined infant and maternal immunisation program from a societal perspective at the WTP of $50,000 per QALY gained. All strategies were compared to the baseline with no intervention, and with the inclusion of monetary loss of life due to RSV-related infant mortality. Since the monetary loss of life, as an indirect cost, does not affect cost-effectiveness from a healthcare perspective, the results of Table 5 and 6 of the main text are applicable for the healthcare perspective.

| Nirsevimab PPD, $ | RSVpreF PPD, $ | Incremental costs, $ (95% CI) | QALYs gained (95% CI) | ICER, $/QALY (95% CI) | Probability of being cost-effective | Budget impact per 100,000 population, $ |
|---|---|---|---|---|---|---|
| *Societal perspective, sigmoidal vaccine efficacy profiles* | | | | | | |
| 1,000 | 460 | 162,923<br>(125,144 to 198,706) | 3.282<br>(2.589 to 4.023) | 49,639<br>(3,1034 to 77,219) | 50% | 371,090 |
| 580 | 475 | 163,549<br>(125,058 to 200,848) | 3.282<br>(2.589 to 4.023) | 49,735<br>(30,880 to 78,871) | 50% | 370,880 |
| *Societal perspective, constant vaccine efficacy profiles* | | | | | | |
| 1,000 | 510 | 171,165<br>(130,776 to 208,772) | 3.526<br>(2.778 to 4.344) | 48,549<br>(30,289 to 75,213) | 55% | 400,735 |

| | | | | | | |
|---|---|---|---|---|---|---|
| 575 | 525 | 172,376 (132,556 to 210,729) | 3.526 (2.778 to 4.344) | 49,035 (30,833 to 76,584) | 53% | 401,464 |

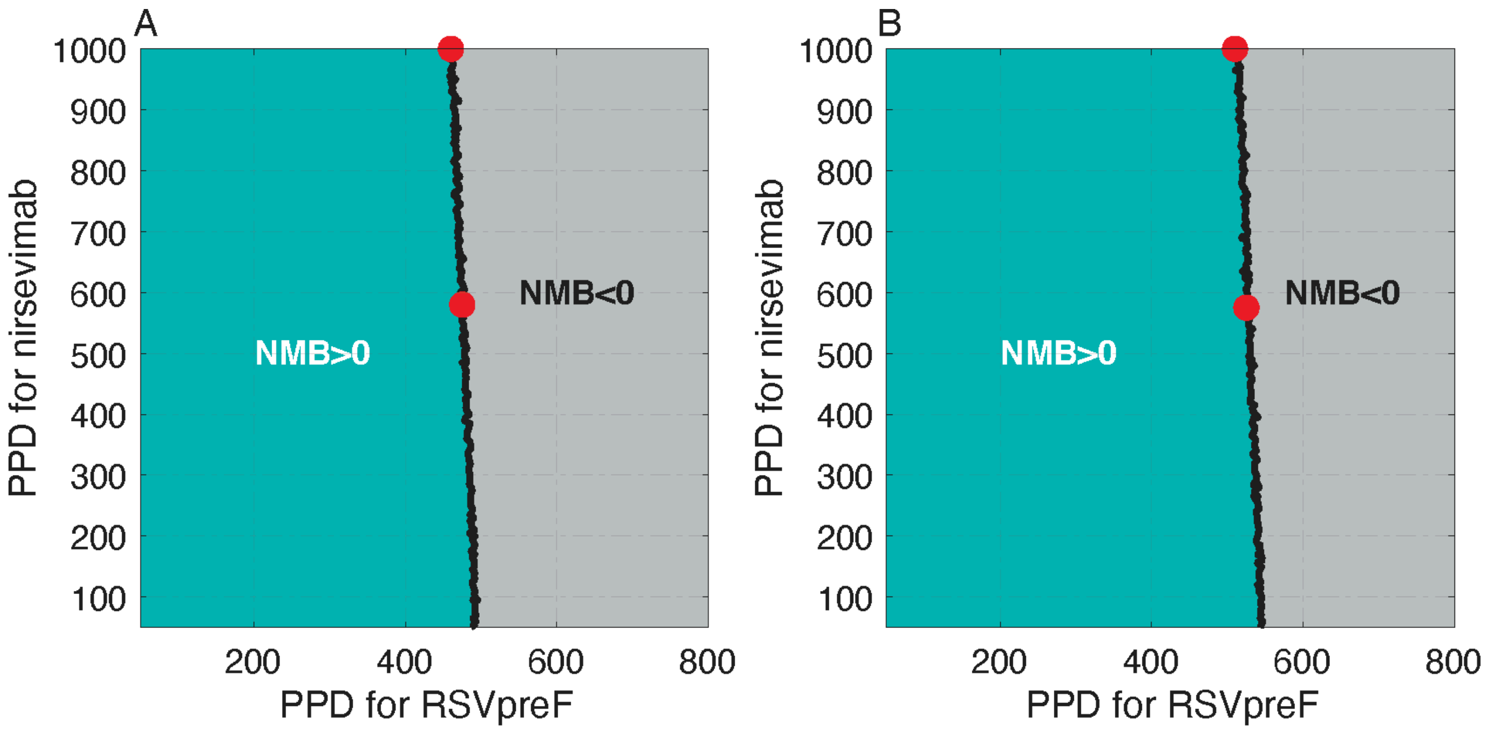

**Figure A6.** Net monetary benefit (NMB) of the combined infant and maternal immunisation program at the WTP of $50,000 per QALY gained as a function of PPD for nirsevimab and RSVpreF. Panels (A) and (B) correspond to the analysis from a societal perspective with sigmoidal and constant vaccine efficacy profiles, respectively. Red circles correspond to the PPD values in Table A8.

**4.2. 80% coverage of nirsevimab and 60% coverage of maternal vaccination**

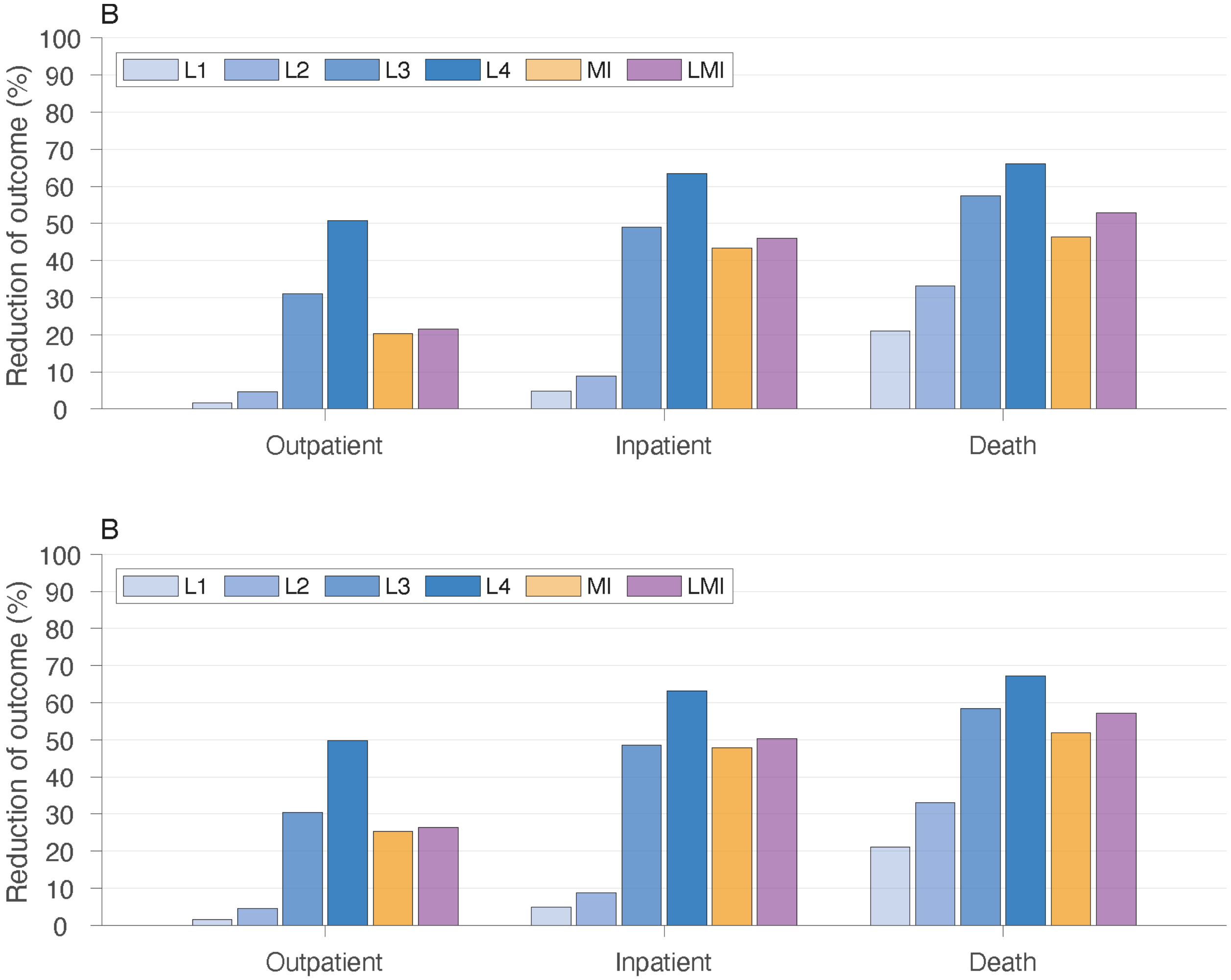

**Figure A7.** Overall reduction of RSV-related outpatient care (office and ED visits), inpatient care (paediatric ward and ICU admissions), and death among infants under one year of age for standalone immunisation programs with nirsevimab (L1, L2, L3, L4) and RSVpreF (MI), and combined nirsevimab and RSV-preF immunisation program (LMI), compared to the scenario without any prevention strategy. Panel (A) and (B) correspond to the sigmoidal and constant vaccine efficacy profiles, respectively.

**Reduction of health outcomes presented in Figure A7 are summarised in Table A9.**

**Table A9.** Reduction of RSV-related outcomes (%) in intervention programs compared to no intervention with 80% coverage of nirsevimab and 60% coverage of maternal vaccination.

| Intervention | Outpatient (95% CI) | Inpatient (95% CI) | Death (95% CI) |
|---|---|---|---|
| *Sigmoidal vaccine efficacy profiles* | | | |
| L1 | 1.6 (1.6 to 1.7) | 4.9 (4.5 to 5.3) | 21.1 (13.0 to 29.8) |
| L2 | 4.7 (4.7 to 4.8) | 9.0 (8.5 to 9.5) | 33.1 (23.5 to 43.3) |
| L3 | 31.1 (31.0 to 31.2) | 49.0 (48.1 to 49.9) | 57.1 (47.3 to 67.0) |
| L4 | 50.7 (50.5 to 50.8) | 63.5 (62.7 to 64.5) | 65.9 (56.0 to 75.6) |
| MI | 20.4 (20.3 to 20.5) | 43.4 (42.5 to 44.2) | 46.0 (35.4 to 57.8) |
| LMI | 21.6 (21.4 to 21.7) | 46.1 (45.2 to 46.9) | 52.6 (42.0 to 63.3) |
| *Sigmoidal vaccine efficacy profiles* | | | |
| L1 | 1.6 (1.6 to 1.6) | 4.9 (4.5 to 5.2) | 21.1 (13.0 to 29.8) |
| L2 | 4.6 (4.6 to 4.7) | 8.9 (8.4 to 9.4) | 33.1 (23.5 to 43.3) |
| L3 | 30.5 (30.4 to 30.6) | 48.7 (47.8 to 49.6) | 58.2 (48.6 to 67.9) |
| L4 | 49.8 (49.7 to 49.9) | 63.2 (62.4 to 64.1) | 67.1 (58.1 to 76.9) |
| MI | 25.3 (25.2 to 25.4) | 47.9 (47.0 to 48.7) | 51.6 (41.4 to 62.4) |
| LMI | 26.4 (26.3 to 26.5) | 50.3 (49.4 to 51.2) | 57.1 (46.9 to 67.7) |

##### 4.2.1. ***Cost-effectiveness analysis without monetary loss of life***

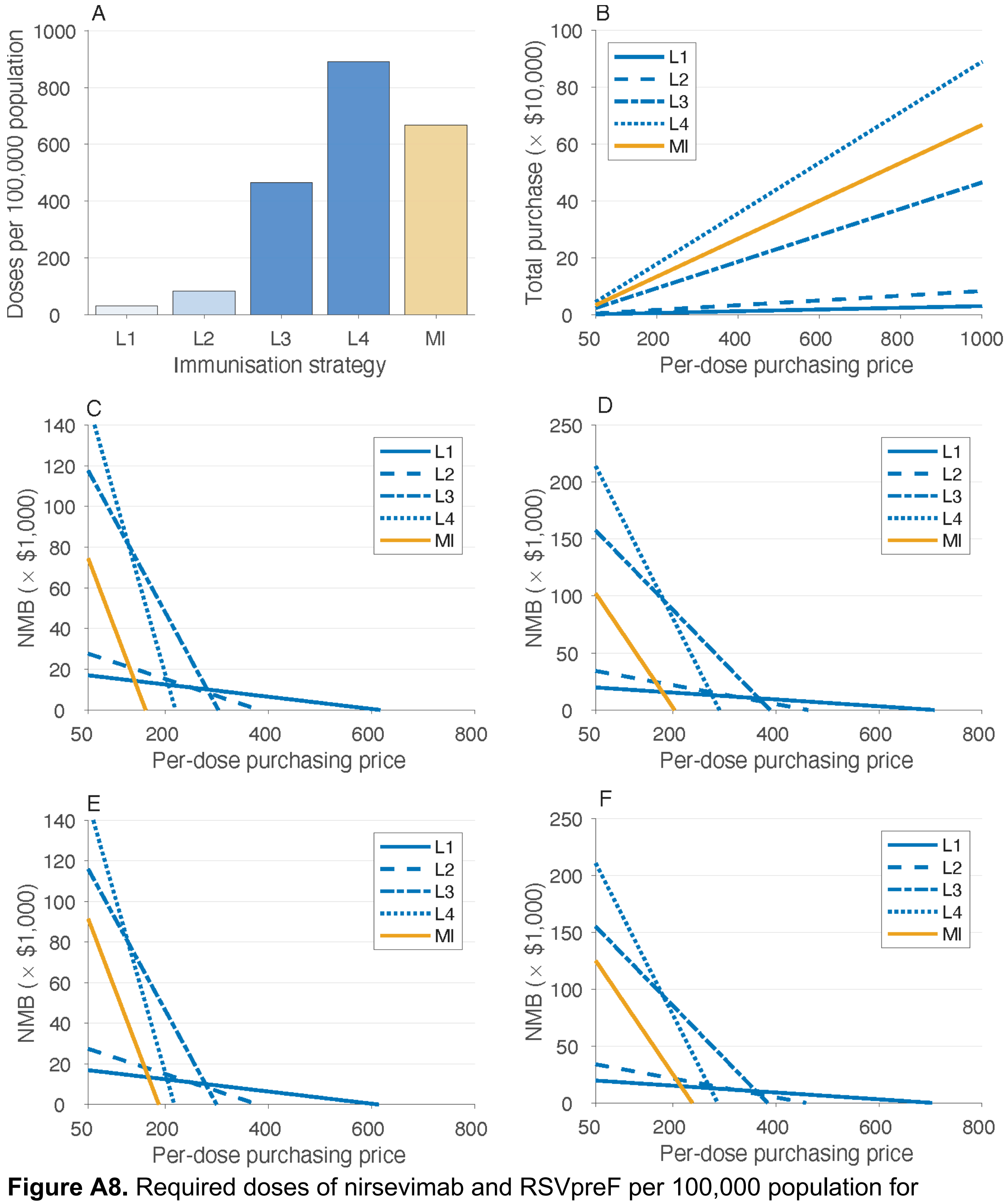

**Figure A8.** Required doses of nirsevimab and RSVpreF per 100,000 population for immunisation strategies (A), with total purchasing costs (B), and the estimated net monetary benefit (NMB) as a function of price per dose at the WTP threshold of $50,000 per QALY gained. Panels (C) and (D) correspond to the analysis from healthcare and societal perspectives, respectively, with sigmoidal vaccine efficacy profiles. Panels (E) and (F) correspond to the analysis from the healthcare and societal perspectives, respectively, with constant vaccine efficacy profiles.

**Table A10.** Model estimates of cost-effectiveness analyses associated with infant and maternal immunisation programs as standalone prevention strategies from healthcare and societal perspectives at the WTP of $50,000 per QALY gained, using sigmoidal vaccine efficacy profiles. All strategies were compared to the baseline with no intervention.

| Prevention strategy | Maximum PPD, $ | Incremental costs, $ (95% CI) | QALYs gained (95% CI) | ICER, $/QALY (95% CI) | Probability of being cost-effective | Budget impact per 100,000 population, $ |
|---|---|---|---|---|---|---|
| *Healthcare perspective* | | | | | | |
| L1 | 615 | 1,001 (-368 to 2,312) | 0.020 (0.014 to 0.027) | 49,074 (-17,909 to 13,0531) | 50% | 1,029 |
| L2 | 380 | 1,338 (-249 to 2,871) | 0.031 (0.024 to 0.040) | 42,543 (-7,968 to 100,737) | 61% | 1,368 |
| L3 | 300 | 2,612 (-274 to 5,414) | 0.077 (0.066 to 0.088) | 33,985 (-3,586 to 73,369) | 79% | 2,692 |
| L4 | 215 | 648 (-2,982 to 4,230) | 0.091 (0.080 to 0.103) | 7,136 (-32,462 to 47,950) | 98% | 687 |
| MI | 160 | 2,389 (-318 to 5,013) | 0.068 (0.058 to 0.080) | 34,976 (-4,485 to 77,612) | 76% | 2,551 |
| *Societal perspective* | | | | | | |
| L1 | 705 | 956 (-485 to 2,355) | 0.020 (0.014 to 0.028) | 46,730 (-22,840 to 132,578) | 53% | 3,734 |
| L2 | 460 | 1,420 (-233 to 3,041) | 0.031 (0.024 to 0.040) | 45,175 (-7,152 to 106,659) | 56% | 8,015 |
| L3 | 385 | 2,203 (-1,187 to 5,577) | 0.077 (0.066 to 0.089) | 28,577 (-15,159 to 74,875) | 83% | 42,242 |

| | | | | | | |
|---|---|---|---|---|---|---|
| L4 | 290 | 4,424<br>(215 to 8,673) | 0.091<br>(0.080 to 0.102) | 48,691<br>(2,268 to 98,420) | 52% | 67,469 |
| MI | 200 | 1,334<br>(-1,762 to 4,364) | 0.068<br>(0.058 to 0.080) | 19,525<br>(-25,258 to 66,318) | 90% | 29,245 |

**Table A11.** Model estimates of cost-effectiveness analyses associated with infant and maternal immunisation programs as standalone prevention strategies from healthcare and societal perspectives at the WTP of $50,000 per QALY gained, using constant vaccine efficacy profiles. All strategies were compared to the baseline with no intervention.

| Prevention strategy | Maximum PPD, $ | Incremental costs, $ (95% CI) | QALYs gained (95% CI) | ICER, $/QALY (95% CI) | Probability of being cost-effective | Budget impact per 100,000 population, $ |
|---|---|---|---|---|---|---|
| *Healthcare perspective* | | | | | | |
| L1 | 610 | 951<br>(-383 to 2,300) | 0.020<br>(0.014 to 0.027) | 46,605<br>(-17,996 to 128,074) | 54% | 982 |
| L2 | 375 | 1,172<br>(-385 to 2,683) | 0.031<br>(0.024 to 0.040) | 37,266<br>(-11,347 to 93,874) | 68% | 1,194 |
| L3 | 295 | 1,873<br>(-1,054 to 4,742) | 0.077<br>(0.066 to 0.088) | 24,335<br>(-13,546 to 64,696) | 90% | 1,929 |
| L4 | 215 | 2,681<br>(-927 to 6,227) | 0.091<br>(0.080 to 0.103) | 29,490<br>(-10,022 to 70,933) | 84% | 2,721 |
| MI | 185 | 2,309<br>(-599 to 5,097) | 0.073<br>(0.062 to 0.085) | 31,544<br>(-7,883 to 73,556) | 81% | 2,502 |
| *Societal perspective* | | | | | | |
| L1 | 700 | 945 | 0.020 | 46,279 | 53% | 3,688 |

| | | | | | | |
|---|---|---|---|---|---|---|
| | | (-470 to 2,348) | (0.014 to 0.027) | (-21,844 to 13,3081) | | |
| L2 | 455 | 1,337<br>(-297 to 2,947) | 0.031<br>(0.024 to 0.040) | 42,566<br>(-9,033 to 103,820) | 61% | 7,840 |
| L3 | 380 | 2,240<br>(-1,077 to 5,484) | 0.077<br>(0.066 to 0.088) | 29,101<br>(-13,567 to 73,876) | 82% | 41,479 |
| L4 | 285 | 3,011<br>(-1,284 to 7,328) | 0.091<br>(0.080 to 0.103) | 33,127<br>(-13,956 to 83,476) | 75% | 65,050 |
| MI | 235 | 2,030<br>(-1,207 to 5,259) | 0.073<br>(0.063 to 0.085) | 27,715<br>(-15,848 to 74,388) | 83% | 35,869 |

**Table A12.** Model estimates of cost-effectiveness analyses associated with the combined infant and maternal immunisation program from healthcare and societal perspectives at the WTP of $50,000 per QALY gained, using sigmoidal vaccine efficacy profiles. All strategies were compared to the baseline with no intervention.

| Nirsevimab PPD, $ | RSVpreF PPD, $ | Incremental costs, $ (95% CI) | QALYs gained (95% CI) | ICER, $/QALY (95% CI) | Probability of being cost-effective | Budget impact per 100,000 population, $ |
|---|---|---|---|---|---|---|
| *Healthcare perspective* | | | | | | |
| 615 | 145 | 1854<br>(-1090 to 4734) | 0.075<br>(0.064 to 0.087) | 24,640<br>(-14,443 to 66,016) | 89% | 1,880 |
| 215 | 165 | 3,173<br>(221 to 6,074) | 0.075<br>(0.064 to 0.087) | 42,208<br>(2,806 to 84,576) | 65% | 3,204 |
| *Societal perspective* | | | | | | |

| | | | | | | |
|---|---|---|---|---|---|---|
| 705 | 185 | 1,733<br>(-1,543 to 4,974) | 0.075<br>(0.064 to 0.087) | 23,028<br>(-19,838 to 69,035) | 88% | 31,279 |
| 290 | 205 | 2,582<br>(-650 to 5,822) | 0.075<br>(0.064 to 0.087) | 34,267<br>(-8,500 to 81,059) | 75% | 32,152 |

**Table A13.** Model estimates of cost-effectiveness analyses associated with the combined infant and maternal immunisation program from healthcare and societal perspectives at the WTP of $50,000 per QALY gained, using constant vaccine efficacy profiles. All strategies were compared to the baseline with no intervention.

| Nirsevimab PPD, $ | RSVpreF PPD, $ | Incremental costs, $ (95% CI) | QALYs gained (95% CI) | ICER, $/QALY (95% CI) | Probability of being cost-effective | Budget impact per 100,000 population, $ |
|---|---|---|---|---|---|---|
| *Healthcare perspective* | | | | | | |
| 610 | 170 | 2,524<br>(-492 to 5,541) | 0.079<br>(0.068 to 0.091) | 31,790<br>(-6,126 to 72,815) | 81% | 2,567 |
| 215 | 185 | 688<br>(-2,365 to 3,668) | 0.079<br>(0.068 to 0.091) | 8,660<br>(-29,622 to 48,272) | 98% | 705 |
| *Societal perspective* | | | | | | |
| 700 | 220 | 3,445<br>(-91 to 6,901) | 0.079<br>(0.068 to 0.091) | 43,375<br>(-1,130 to 90,441) | 61% | 38,639 |
| 285 | 235 | 933<br>(-2,474 to 4,439) | 0.079<br>(0.068 to 0.091) | 11,750<br>(-30,796 to 57,765) | 95% | 36,175 |

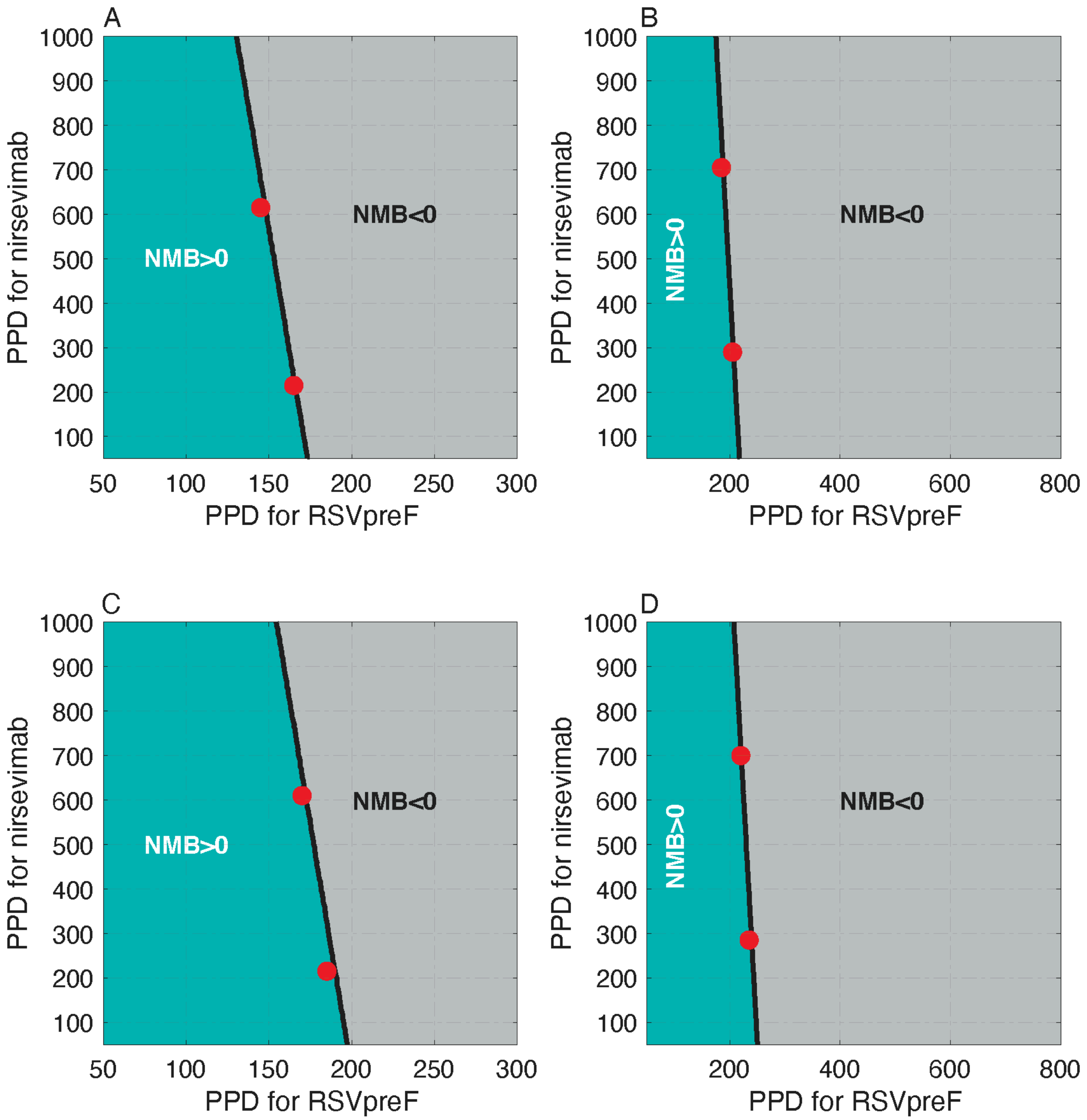

**Figure A9.** Net monetary benefit (NMB) of the combined infant and maternal immunisation program at the WTP of $50,000 per QALY gained as a function of PPD for nirsevimab and RSVpreF. Panels (A) and (B) correspond to the analysis from healthcare and societal perspectives, respectively, with sigmoidal vaccine efficacy profiles. Panels (C) and (D) correspond to the analysis from the healthcare and societal perspectives, respectively, with constant vaccine efficacy profiles. Red circles correspond to the PPD values in Tables A12 and A13.

###### 4.2.2. ***Cost-effectiveness analysis with monetary loss of life***

Cost-effectiveness analyses in this section include the monetary loss of life as an indirect cost from a societal perspective. Reductions of RSV-related outcomes remain the same as reported in Figure A7 and Table A6.

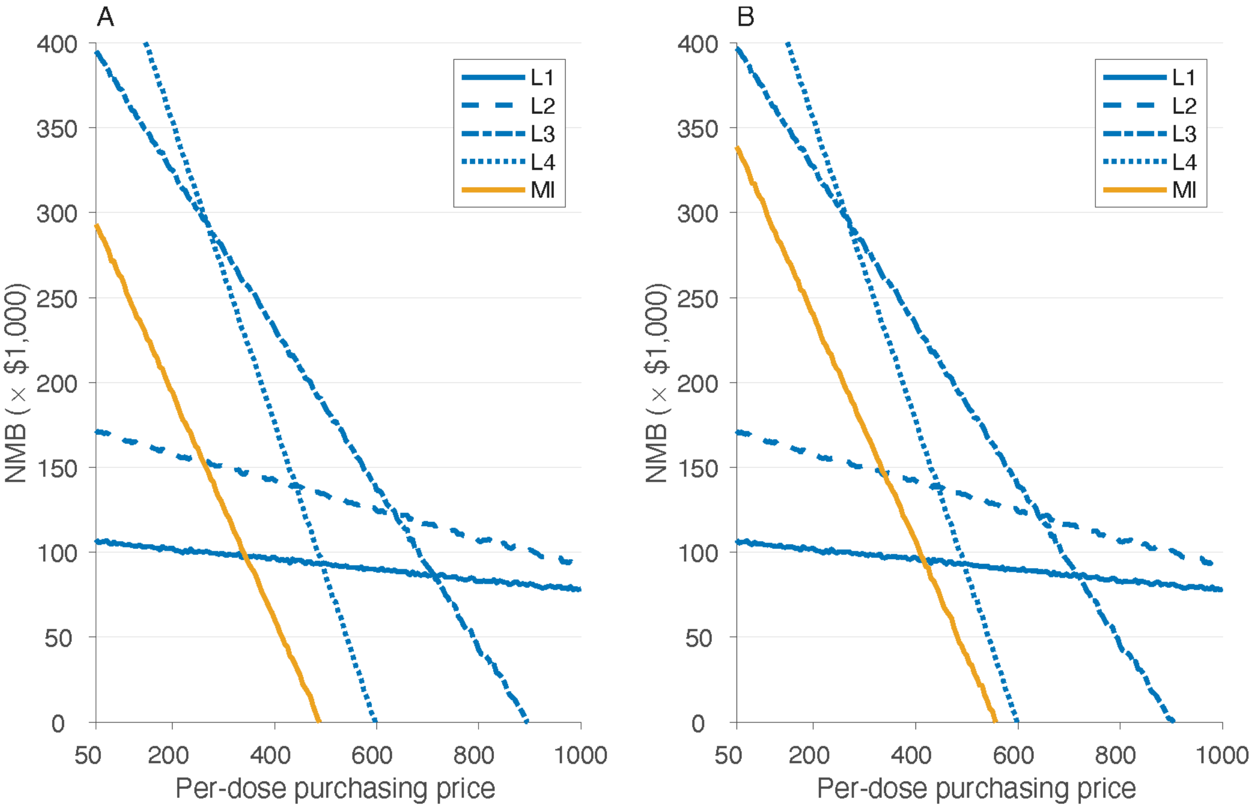

**Figure A10.** Estimated net monetary benefit (NMB) as a function of price per dose at the WTP threshold of $50,000 per QALY gained with the inclusion of monetary loss of life due to RSV-related infant mortality. Panels (A) and (B) correspond to the analysis from a societal perspective using sigmoidal and constant vaccine efficacy profiles, respectively. Note that the inclusion of monetary loss due to infant mortality (as an indirect cost) does not affect cost-effectiveness analysis from a healthcare perspective.

**Table A14.** Model estimates of cost-effectiveness analyses associated with infant and maternal immunisation programs as standalone prevention strategies from a societal perspective at the WTP of $50,000 per QALY gained. All strategies were compared to the baseline with no intervention with the inclusion of monetary loss of life due to RSV-related infant mortality. Since the monetary loss of life, as an indirect cost, does not affect cost-effectiveness from a healthcare perspective, the results of Table A10 and A11 are applicable for the healthcare perspective.

| Prevention strategy | Maximum PPD, $ | Incremental costs, $ (95% CI) | QALYs gained (95% CI) | ICER, $/QALY (95% CI) | Probability of being cost-effective | Budget impact per 100,000 population, $ |
|---|---|---|---|---|---|---|
| *Societal perspective, sigmoidal vaccine efficacy profiles* | | | | | | |
| L1 | 1,000 | -33,700<br>(-54,217 to -15,243) | 0.881<br>(0.513 to 1.295) | -38,274<br>(-42,304 to -29,416) | Cost-saving | 12,601 |
| L2 | 1,000 | -22,491<br>(-48,607 to 1,060) | 1.390<br>(0.930 to 1.898) | -16,175<br>(-25,432 to 1,140) | 100% | 52,877 |
| L3 | 895 | 120,358<br>(86,526 to 151,470) | 2.432<br>(1.832 to 3.122) | 49,485<br>(27,795 tp 83,415) | 50% | 279,546 |
| L4 | 595 | 138,753<br>(104,526 to 172,047) | 2.803<br>(2.163 to 3.499) | 49,509<br>(30,004 to 80,034) | 51% | 339,048 |
| MI | 490 | 98,474<br>(67,486 to 126,963) | 1.973<br>(1.417 to 2.572) | 49,907<br>(26,051 to 89,830) | 49% | 222,614 |
| *Societal perspective, constant vaccine efficacy profiles* | | | | | | |
| L1 | 1,000 | -33,560<br>(-54,101 to -15,073) | 0.880<br>(0.513 to 1.295) | -38,115<br>(-42,186 to -29,129) | Cost-saving | 12,705 |
| L2 | 1,000 | -22,153<br>(-48,236 to 1,425) | 1.390<br>(0.930 to 1.898) | -15,932<br>(-25,280 to 1,532) | 100% | 53,118 |
| L3 | 900 | 122,582 | 2.480 | 49,419 | 51% | 283,436 |

| | | | | | | |
|---|---|---|---|---|---|---|
| | | (89,659 to 153,435) | (1.878 to 3.124) | (28,662 to 81,814) | | |
| L4 | 595 | 139,555 (104,894 to 173,201) | 2.847 (2.208 to 3.544) | 49,014 (29,618 to 78,922) | 53% | 341,082 |
| MI | 555 | 107,807 (75,203 to 138,403) | 2.203 (1.602 to 2.849) | 48,936 (26,452 to 86,427) | 52% | 205,850 |

**Table A15.** Model estimates of cost-effectiveness analyses associated with the combined infant and maternal immunisation program from a societal perspective at the WTP of $50,000 per QALY gained. All strategies were compared to the baseline with no intervention with the inclusion of monetary loss of life due to RSV-related infant mortality. Since the monetary loss of life, as an indirect cost, does not affect cost-effectiveness from a healthcare perspective, the results of Table A12 and A13 of the previous section are applicable for the healthcare perspective.

| Nirsevimab PPD, $ | RSVpreF PPD, $ | Incremental costs, $ (95% CI) | QALYs gained (95% CI) | ICER, $/QALY (95% CI) | Probability of being cost-effective | Budget impact per 100,000 population, $ |
|---|---|---|---|---|---|---|
| *Societal perspective, sigmoidal vaccine efficacy profiles* | | | | | | |
| 1,000 | 500 | 110,671 (78,640 to 140,196) | 2.251 (1.651 to 2.895) | 49,160 (27,185 to 84,425) | 51% | 250,357 |
| 595 | 520 | 111,898 (79,222 to 142,298) | 2.251 (1.651 to 2.895) | 49,723 (27,292 to 86,213) | 50% | 251,530 |
| *Societal perspective, constant vaccine efficacy profiles* | | | | | | |
| 1,000 | 560 | 120,062 (86,604 to 151,712) | 2.438 (1.790 to 3.081) | 49,284 (27,996 to 83,419) | 51% | 274,550 |
| 595 | 580 | 121,126 | 2.438 | 49,688 | 51% | 275,724 |

| | | | | |
|---|---|---|---|---|
| | | (88,039 to 153,428) | (1.790 to 3.081) | (28,559 to 85,448) |

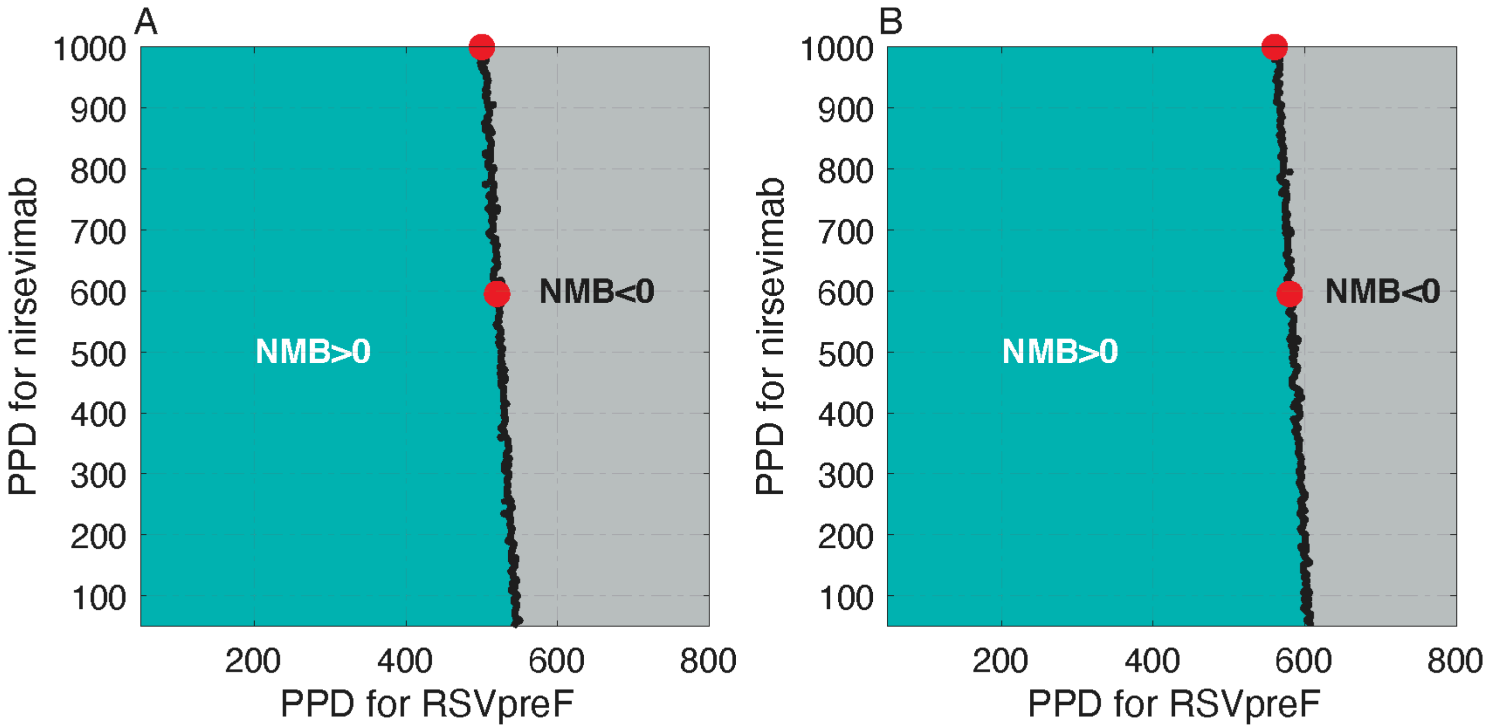

**Figure A11.** Net monetary benefit (NMB) of the combined infant and maternal immunisation program at the WTP of $50,000 per QALY gained as a function of PPD for nirsevimab and RSVpreF. Panels (A) and (B) correspond to the analysis from a societal perspective with sigmoidal and constant vaccine efficacy profiles, respectively. Red circles correspond to the PPD values in Table A15.

#### 5. Secondary analyses with a WTP of $30,000 per QALY gained

##### 5.1. 100% coverage of nirsevimab and 100% coverage of maternal vaccination

The reductions of health outcomes for this scenario are the same as those reported in the main text at the WTP of $50,000 per QALY gained.

###### 5.1.1. *Cost-effectiveness analysis without monetary loss of life*

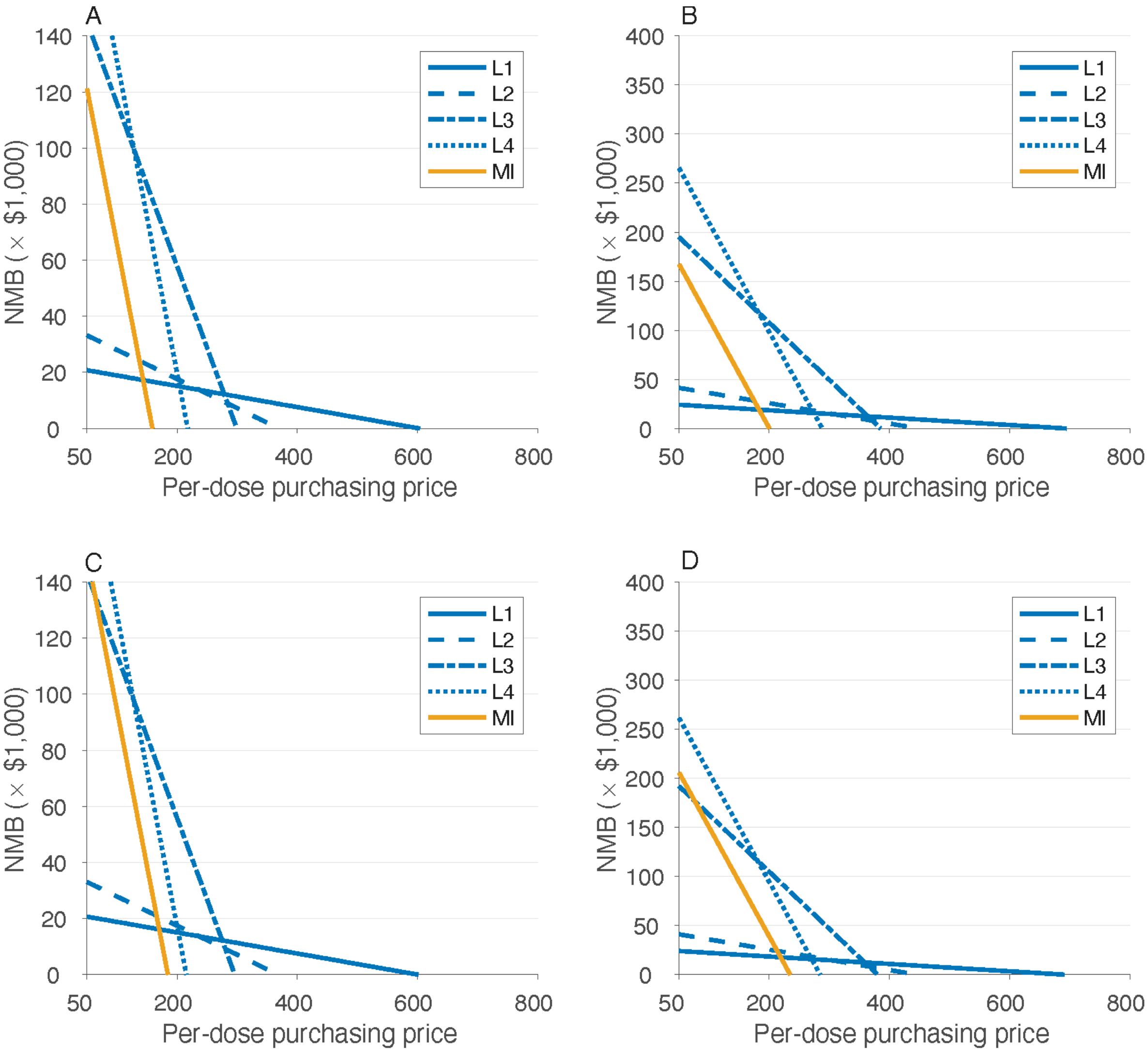

**Figure A12.** Estimates of the net monetary benefit (NMB) as a function of price per dose at the WTP threshold of $30,000 per QALY gained. Panels (A) and (B) correspond to the analysis from healthcare and societal perspectives, respectively, with sigmoidal vaccine efficacy profiles. Panels (C) and (D) correspond to the analysis from the healthcare and societal perspectives, respectively, with constant vaccine efficacy profiles.

**Table A16.** Model estimates of cost-effectiveness analyses associated with infant and maternal immunisation programs as standalone prevention strategies from healthcare and societal perspectives at the WTP of $30,000 per QALY gained, using sigmoidal vaccine efficacy profiles. All strategies were compared to the baseline with no intervention.

| Prevention strategy | Maximum PPD, $ | Incremental costs, $ (95% CI) | QALYs gained (95% CI) | ICER, $/QALY (95% CI) | Probability of being cost-effective | Budget impact per 100,000 population, $ |
|---|---|---|---|---|---|---|
| *Healthcare perspective* | | | | | | |
| L1 | 600 | 629<br>(-924 to 2,110) | 0.024<br>(0.018 to 0.032) | 25,985<br>(-36,445 to 97,260) | 54% | 661 |
| L2 | 365 | 597<br>(-1,164 to 2,349) | 0.036<br>(0.028 to 0.045) | 16,639<br>(-32,325 to 70,184) | 69% | 630 |
| L3 | 295 | 348<br>(-3,130 to 3,814) | 0.094<br>(0.082 to 0.107) | 3,695<br>(-33,149 to 41,600) | 92% | 395 |
| L4 | 215 | 467<br>(-3,878 to 4,708) | 0.111<br>(0.099 to 0.124) | 4,200<br>(-34,697 to 43,384) | 90% | 503 |
| MI | 155 | -1,115<br>(-4,932 to 2,682) | 0.109<br>(0.096 to 0.123) | -10,214<br>(-44,180 to 25,029) | 99% | -1,019 |
| *Societal perspective* | | | | | | |
| L1 | 690 | 603<br>(-1,029 to 2,214) | 0.024<br>(0.018 to 0.032) | 24,866<br>(-41,045 to 102,508) | 55% | 4,043 |
| L2 | 445 | 740<br>(-1,160 to 2,568) | 0.036<br>(0.028 to 0.045) | 20,674<br>(-32,467 to 77,558) | 63% | 8,937 |
| L3 | 385 | 2,705<br>(-1,342 to 6,703) | 0.094<br>(0.082 to 0.107) | 28,634<br>(-13,811 to 73,395) | 52% | 52,738 |

| | | | | | | |
|---|---|---|---|---|---|---|
| L4 | 285 | -378<br>(-5,674 to 4,892) | 0.111<br>(0.099 to 0.124) | -3403<br>(-50,404 to 45,184) | 91% | 78,413 |
| MI | 200 | 2,816<br>(-1,495 to 7,037) | 0.109<br>(0.096 to 0.123) | 25,815<br>(-13,217 to 66,816) | 58% | 49,066 |

**Table A17.** Model estimates of cost-effectiveness analyses associated with infant and maternal immunisation programs as standalone prevention strategies from healthcare and societal perspectives at the WTP of $30,000 per QALY gained, using constant vaccine efficacy profiles. All strategies were compared to the baseline with no intervention.

| Prevention strategy | Maximum PPD, $ | Incremental costs, $ (95% CI) | QALYs gained (95% CI) | ICER, $/QALY (95% CI) | Probability of being cost-effective | Budget impact per 100,000 population, $ |
|---|---|---|---|---|---|---|
| *Healthcare perspective* | | | | | | |
| L1 | 595 | 541<br>(-989 to 2,036) | 0.024<br>(0.018 to 0.032) | 22,417<br>(-40,159 to 94,033) | 59% | 581 |
| L2 | 365 | 776<br>(-976 to 2,547) | 0.036<br>(0.028 to 0.045) | 21,640<br>(-27,237 to 76,304) | 62% | 811 |
| L3 | 295 | 2,311<br>(-1,131 to 5,714) | 0.094<br>(0.082 to 0.106) | 24,716<br>(-11,905 to 63,068) | 61% | 2,354 |
| L4 | 215 | 3,017<br>(-1,327 to 7,221) | 0.110<br>(0.098 to 0.123) | 27,348<br>(-11,810 to 67,888) | 55% | 3,050 |
| MI | 180 | -1,603<br>(-5,725 to 2,439) | 0.117<br>(0.104 to 0.131) | -13,677<br>(-48,261 to 21,179) | 99% | -1,555 |
| *Societal perspective* | | | | | | |

| | | | | | | |
|---|---|---|---|---|---|---|
| L1 | 685 | 562<br>(-1,085 to 2,133) | 0.024<br>(0.018 to 0.031) | 23,259<br>(-43,323 to 98,999) | 57% | 3,962 |
| L2 | 445 | 1,046<br>(-865 to 2,885) | 0.036<br>(0.028 to 0.045) | 29,231<br>(-23,894 to 87,190) | 51% | 9,118 |
| L3 | 380 | 2,751<br>(-1,242 to 6,664) | 0.094<br>(0.082 to 0.106) | 29,422<br>(-13,081 to 73,612) | 51% | 51,790 |
| L4 | 280 | -2,162<br>(-7,370 to 3,042) | 0.110<br>(0.098 to 0.123) | -19,596<br>(-66,256 to 27,983) | 98% | 75,395 |
| MI | 230 | -2,014<br>(-6,770 to 2,707) | 0.117<br>(0.104 to 0.131) | -17,184<br>(-56,975 to 23,531) | 99% | 54,095 |

**Table A18.** Model estimates of cost-effectiveness analyses associated with the combined infant and maternal immunisation program from healthcare and societal perspectives at the WTP of $30,000 per QALY gained, using sigmoidal vaccine efficacy profiles. All strategies were compared to the baseline with no intervention.

| Nirsevimab PPD, $ | RSVpreF PPD, $ | Incremental costs, $ (95% CI) | QALYs gained (95% CI) | ICER, $/QALY (95% CI) | Probability of being cost-effective | Budget impact per 100,000 population, $ |
|---|---|---|---|---|---|---|
| *Healthcare perspective* | | | | | | |
| 600 | 140 | -132<br>(-4,058 to 3,681) | 0.112<br>(0.099 to 0.126) | -1,174<br>(-35,712 to 33,741) | 96% | -97 |
| 215 | 155 | 2,041<br>(-1,846 to 5,892) | 0.112<br>(0.099 to 0.126) | 18,193<br>(-16,004 to 54,135) | 75% | 2,135 |
| *Societal perspective* | | | | | | |

| | | | | | | |
|---|---|---|---|---|---|---|
| 690 | 180 | 86<br>(-4,417 to 4,482) | 0.112<br>(0.099 to 0.126) | 768<br>(-38,946 to 40,828) | 92% | 47,804 |
| 285 | 195 | 1,551<br>(-2,842 to 5,944) | 0.112<br>(0.099 to 0.126) | 13,835<br>(-24,884 to 54,520) | 79% | 45,528 |

**Table 19.** Model estimates of cost-effectiveness analyses associated with the combined infant and maternal immunisation program from healthcare and societal perspectives at the WTP of $30,000 per QALY gained, using constant vaccine efficacy profiles. All strategies were compared to the baseline with no intervention.

| Nirsevimab PPD, $ | RSVpreF PPD, $ | Incremental costs, $ (95% CI) | QALYs gained (95% CI) | ICER, $/QALY (95% CI) | Probability of being cost-effective | Budget impact per 100,000 population, $ |
|---|---|---|---|---|---|---|
| *Healthcare perspective* | | | | | | |
| 595 | 165 | 1,483<br>(-2,557 to 5,532) | 0.119<br>(0.106 to 0.133) | 12,495<br>(-21,445 to 48,068) | 83% | 27,541 |
| 215 | 175 | -1,657<br>(-5,873 to 2,300) | 0.119<br>(0.106 to 0.133) | -13,954<br>(-48,719 to 19,878) | 99% | 24,395 |
| *Societal perspective* | | | | | | |
| 685 | 215 | 3,426<br>(-1,262 to 8,114) | 0.119<br>(0.106 to 0.133) | 28,853<br>(-10,603 to 70,149) | 52% | 60,566 |
| 280 | 225 | -615<br>(-5,364 to 4,115) | 0.119<br>(0.106 to 0.133) | -5,187<br>(-44,579 to 35,417) | 95% | 56481 |

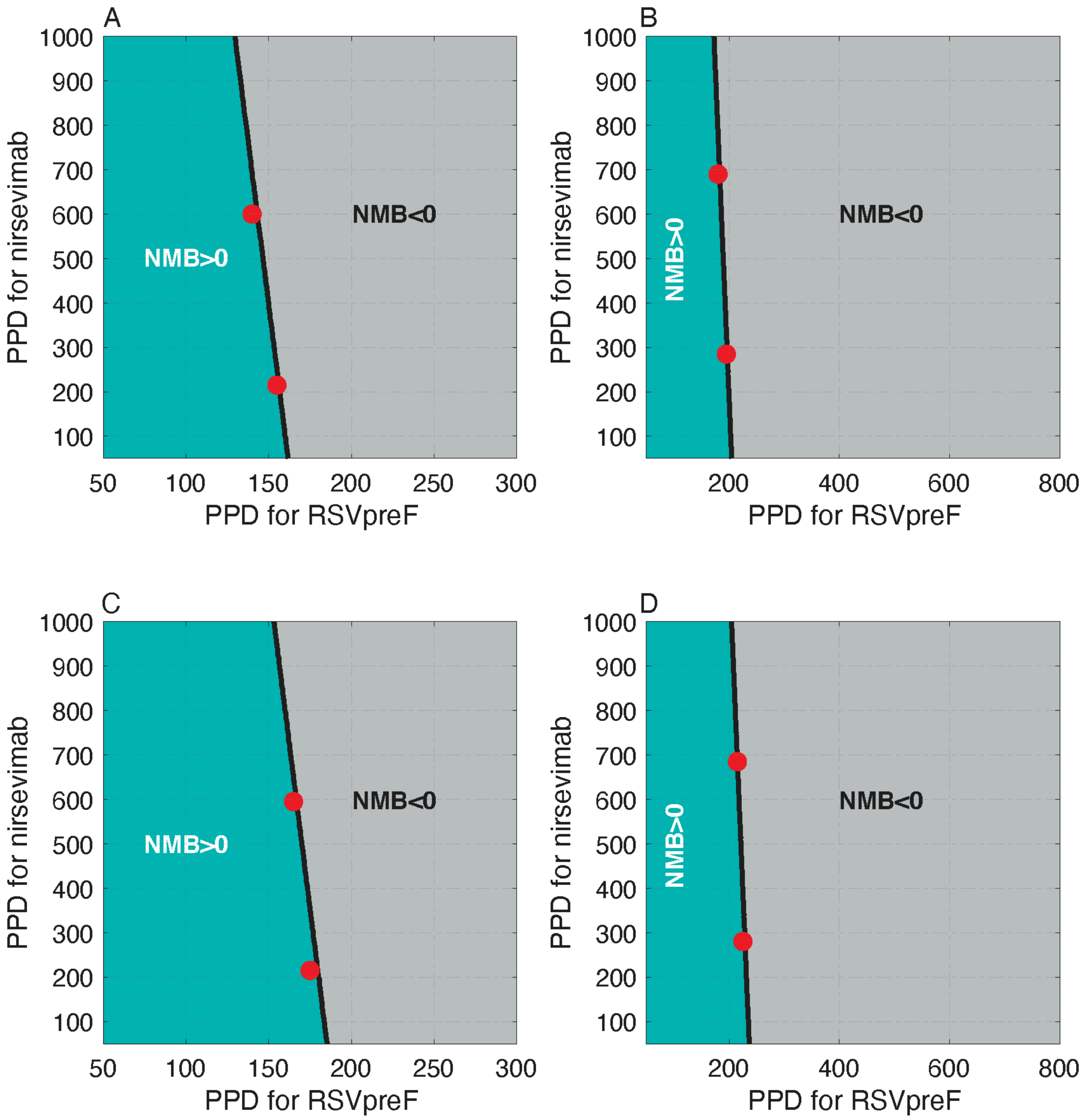

**Figure A13.** Net monetary benefit (NMB) of the combined infant and maternal immunisation program at the WTP of $30,000 per QALY gained as a function of PPD for nirsevimab and RSVpreF. Panels (A) and (B) correspond to the analysis from healthcare and societal perspectives, respectively, with sigmoidal vaccine efficacy profiles. Panels (C) and (D) correspond to the analysis from the healthcare and societal perspectives, respectively, with constant vaccine efficacy profiles. Red circles correspond to the PPD values in Tables A18 and A19.

##### 5.1.2. ***Cost-effectiveness analysis with monetary loss of life***

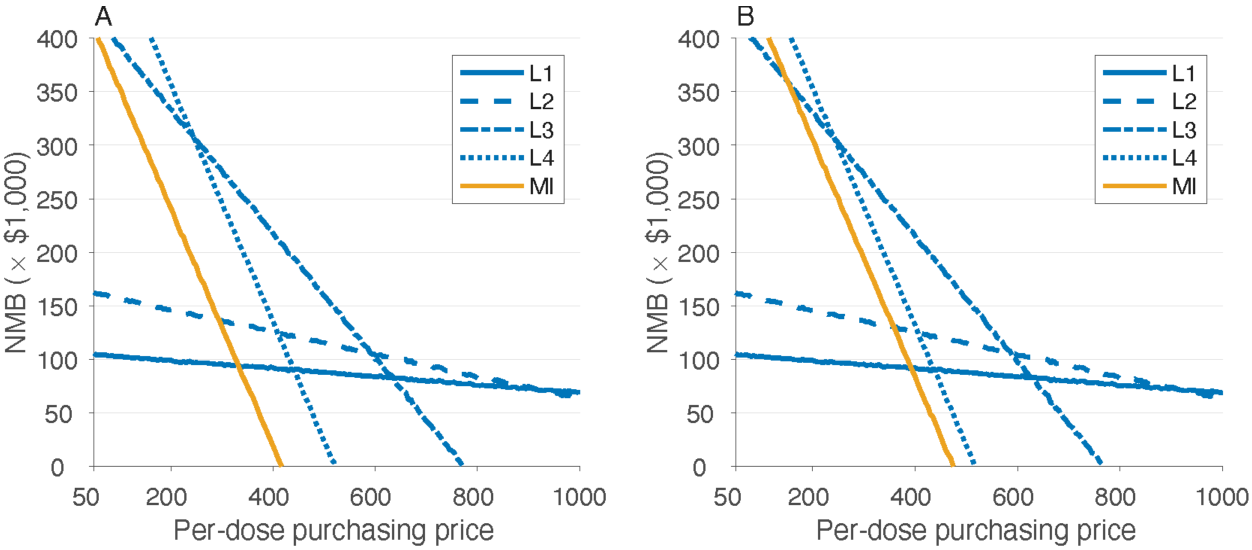

**Figure A14.** Estimated net monetary benefit (NMB) as a function of price per dose at the WTP threshold of $30,000 per QALY gained with the inclusion of monetary loss of life due to RSV-related infant mortality. Panels (A) and (B) correspond to the analysis from a societal perspective using sigmoidal and constant vaccine efficacy profiles, respectively. Note that the inclusion of monetary loss due to infant mortality (as an indirect cost) does not affect cost-effectiveness analysis from a healthcare perspective.

**Table A20.** Model estimates of cost-effectiveness analyses associated with infant and maternal immunisation programs as standalone prevention strategies from a societal perspective at the WTP of $30,000 per QALY gained. All strategies were compared to the baseline with no intervention, and with the inclusion of monetary loss of life due to RSV-related infant mortality. Since the monetary loss of life, as an indirect cost, does not affect cost-effectiveness from a healthcare perspective, the results of Table A16 and A17 of the previous section are applicable for the healthcare perspective.

| Prevention strategy | Maximum PPD, $ | Incremental costs, $ (95% CI) | QALYs gained (95% CI) | ICER, $/QALY (95% CI) | Probability of being cost-effective | Budget impact per 100,000 population, $ |
|---|---|---|---|---|---|---|
| *Societal perspective, sigmoidal vaccine efficacy profiles* | | | | | | |
| L1 | 1,000 | -38,282 (-60,190 to -17,798) | 1.023 (0.607 to 1.479) | -37,439 (-41,487 to -28,904) | Cost-saving | 15,688 |
| L2 | 1,000 | -17,393 (-44,625 to 7,446) | 1.533 (1.025 to 2.083) | -11,345 (-21,523 to 7,164) | 100% | 66,571 |
| L3 | 770 | 84,322 (47,373 to 118,185) | 2.904 (2.256 to 3.595) | 29,000 (13,073 to 53,082) | 53% | 276,652 |
| L4 | 520 | 98,492 (61,202 to 134,922) | 3.324 (2.635 to 4.065) | 29,614 (15,047 to 51,601) | 52% | 339,968 |
| MI | 415 | 91,165 (53,096 to 127,298) | 3.101 (2.406 to 3.882) | 29,485 (13,805 to 53,519) | 51% | 288,361 |
| *Societal perspective, constant vaccine efficacy profiles* | | | | | | |
| L1 | 1,000 | -38,129 (-60,033 to -17,630) | 1.022 (0.607 to 1.479) | -37,291 (-41,376 to -28,707) | Cost-saving | 15,795 |
| L2 | 1,000 | -17,089 (-44,330 to 7,796) | 1.533 (1.025 to 2.083) | -11,147 (-21,381 to 7,553) | 100% | 66,752 |
| L3 | 770 | 87,177 | 2.895 | 29,982 | 49% | 278,611 |

| | | | | | | |
|---|---|---|---|---|---|---|
| | | (50,566 to 120,777) | (2.212 to 3.595) | (13,957 to 53,808) | | |
| L4 | 515 | 96,103<br>(58,229 to 132,136) | 3.327<br>(2.635 to 4.064) | 28,798<br>(14,288 to 50,361) | 55% | 336,950 |
| MI | 470 | 97,623<br>(56,455 to 135,039) | 3.419<br>(2.684 to 4.209) | 28,488<br>(13,359 to 50,188) | 56% | 321,215 |

**Table A21.** Model estimates of cost-effectiveness analyses associated with the combined infant and maternal immunisation program from a societal perspective at the WTP of $30,000 per QALY gained. All strategies were compared to the baseline with no intervention, and with the inclusion of monetary loss of life due to RSV-related infant mortality. Since the monetary loss of life, as an indirect cost, does not affect cost-effectiveness from a healthcare perspective, the results of Table A18 and A19 of the previous section are applicable for the healthcare perspective.

| Nirsevimab PPD, $ | RSVpreF PPD, $ | Incremental costs, $ (95% CI) | QALYs gained (95% CI) | ICER, $/QALY (95% CI) | Probability of being cost-effective | Budget impact per 100,000 population, $ |
|---|---|---|---|---|---|---|
| *Societal perspective, sigmoidal vaccine efficacy profiles* | | | | | | |
| 1,000 | 400 | 96,038<br>(57,312 to 132,076) | 3.282<br>(2.589 to 4.023) | 29,238<br>(14,182 to 51,228) | 52% | 304,310 |
| 520 | 415 | 95119 56690<br>131762 | 3.282<br>(2.589 to 4.023) | 29033 14013<br>51677 | 54% | 302,973 |
| *Societal perspective, constant vaccine efficacy profiles* | | | | | | |
| 1,000 | 450 | 105,185<br>(65,889 to 143,091) | 3.526<br>(2.778 to 4.344) | 29,961<br>(15,346 to 51,799) | 49% | 333,955 |
| 515 | 465 | 103,318 | 3.526 | 29,372 | 52% | 332,430 |

| | | | | |
|---|---|---|---|---|
| | | (63,082 to 141,771) | (2.778 to 4.344) | (14,656 to 51,232) |

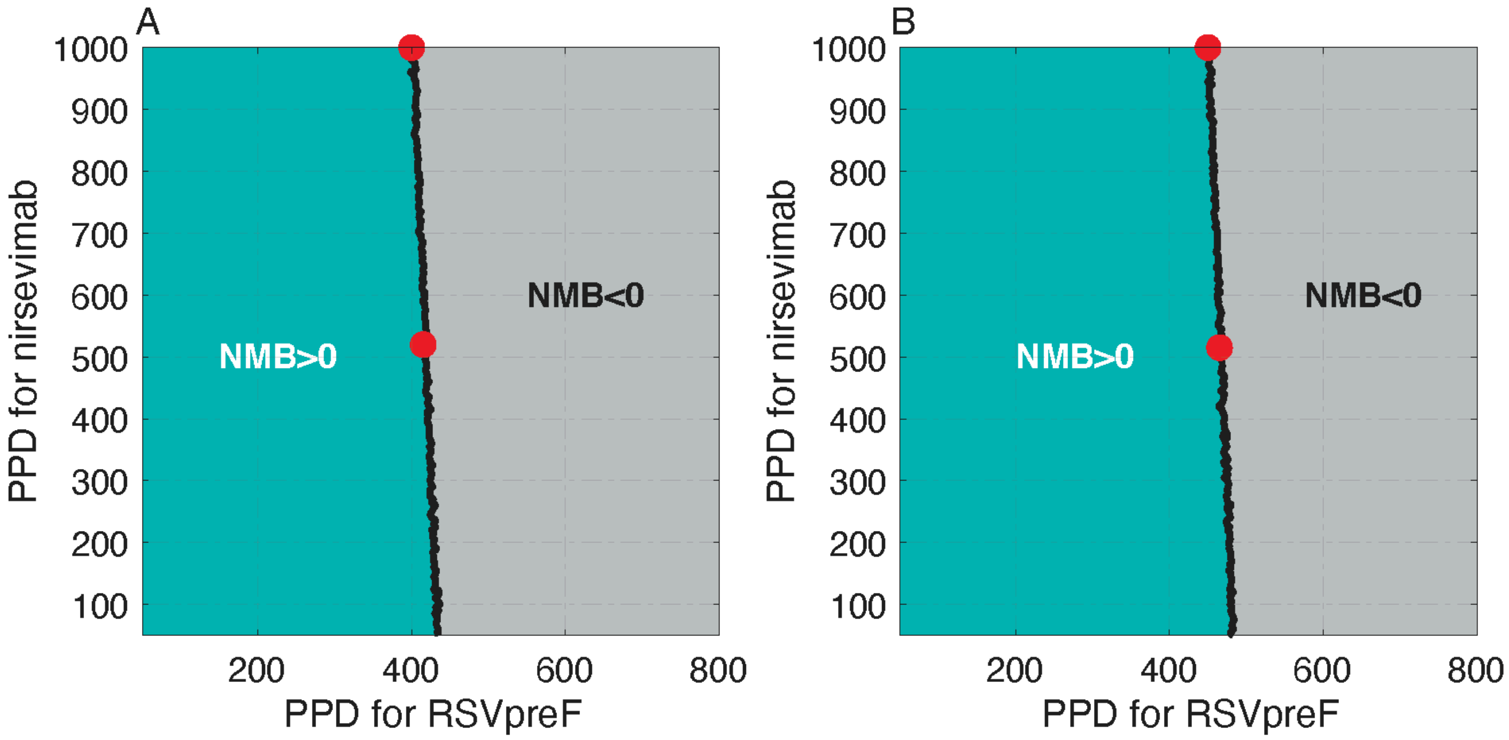

**Figure A15.** Net monetary benefit (NMB) of the combined infant and maternal immunisation program at the WTP of $30,000 per QALY gained as a function of PPD for nirsevimab and RSVpreF. Panels (A) and (B) correspond to the analysis from a societal perspective with sigmoidal and constant vaccine efficacy profiles, respectively. Red circles correspond to the PPD values in Table A21.

##### 5.2. 80% coverage of nirsevimab and 60% coverage of maternal vaccination

Reduction of health outcomes are the same as presented in section 3.2 of this Supplemental.

###### 5.2.1. *Cost-effectiveness analysis without monetary loss of life*

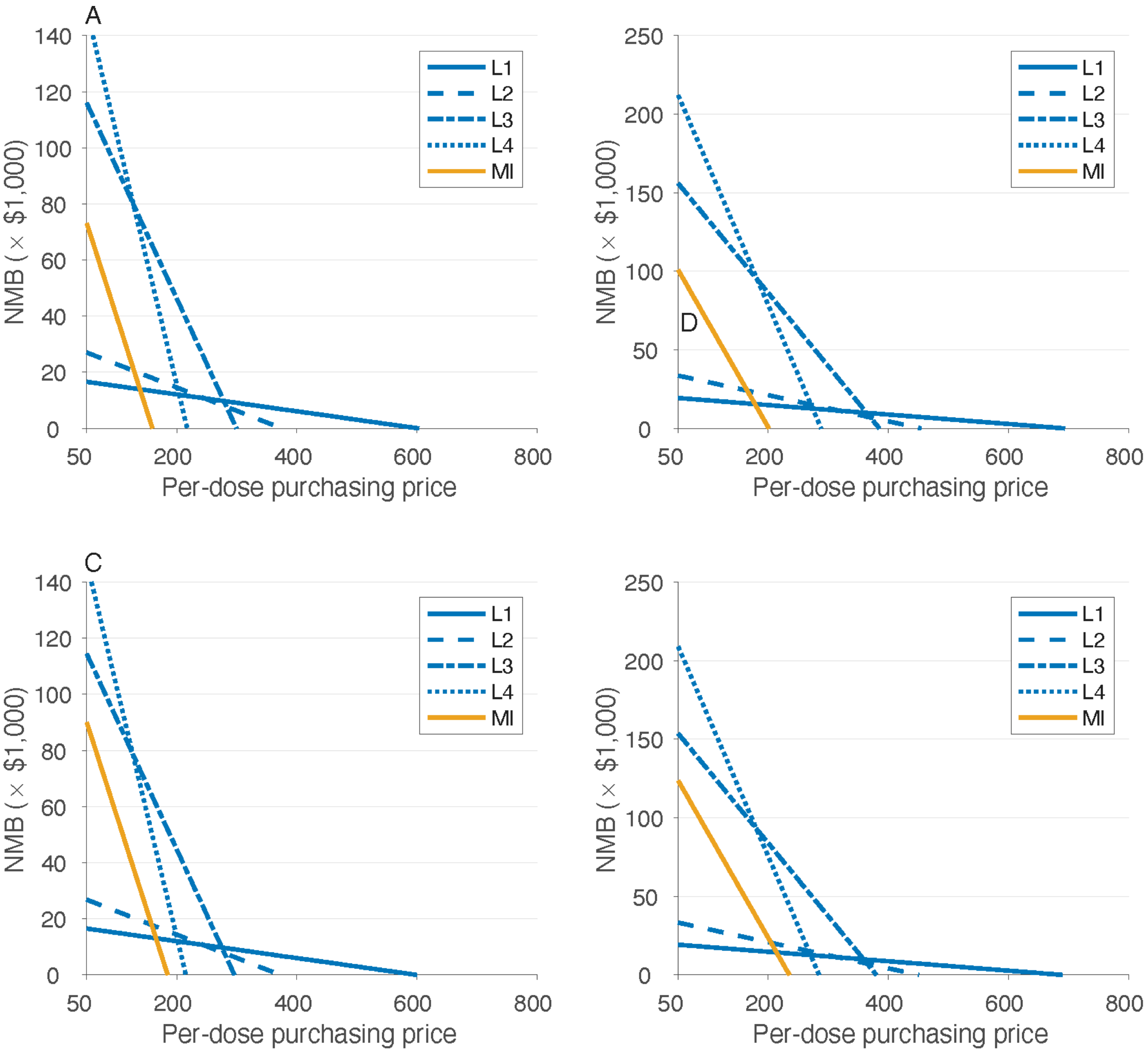

**Figure A16.** Estimates of the net monetary benefit (NMB) as a function of price per dose at the WTP threshold of $30,000 per QALY gained. Panels (A) and (B) correspond to the analysis from healthcare and societal perspectives, respectively, with sigmoidal vaccine efficacy profiles. Panels (C) and (D) correspond to the analysis from the healthcare and societal perspectives, respectively, with constant vaccine efficacy profiles.

**Table A22.** Model estimates of cost-effectiveness analyses associated with infant and maternal immunisation programs as standalone prevention strategies from healthcare and societal perspectives at the WTP of $30,000 per QALY gained, using sigmoidal vaccine efficacy profiles. All strategies were compared to the baseline with no intervention.

| Prevention strategy | Maximum PPD, $ | Incremental costs, $ (95% CI) | QALYs gained (95% CI) | ICER, $/QALY (95% CI) | Probability of being cost-effective | Budget impact per 100,000 population, $ |
|---|---|---|---|---|---|---|
| *Healthcare perspective* | | | | | | |
| L1 | 600 | 549 (-817 to 1,864) | 0.020 (0.014 to 0.027) | 26,966 (-38,162 to 103,237) | 52% | 578 |
| L2 | 375 | 936 (-624 to 2,437) | 0.031 (0.024 to 0.040) | 29,719 (-18,876 to 84,742) | 50% | 953 |
| L3 | 295 | 303 (-2,612 to 3,197) | 0.077 (0.066 to 0.088) | 3,937 (-34,141 to 43,287) | 90% | 365 |
| L4 | 215 | 648 (-2,982 to 4,230) | 0.091 (0.080 to 0.103) | 7,136 (-32,462 to 47,950) | 87% | 687 |
| MI | 155 | -975 (-3,720 to 1,717) | 0.068 (0.058 to 0.080) | -14,269 (-53,712 to 25,789) | 98% | -944 |
| *Societal perspective* | | | | | | |
| L1 | 690 | 515 (-914 to 1,938) | 0.020 (0.014 to 0.028) | 25,188 (-42,664 to 108,526) | 55% | 3,284 |
| L2 | 450 | 610 (-1,029 to 2,226) | 0.031 (0.024 to 0.040) | 19,393 (-3,1978 to 75,957) | 65% | 7,184 |
| L3 | 385 | 2203 (-1,187 to 5,577) | 0.077 (0.066 to 0.089) | 28,577 (-15,159 to 74,875) | 53% | 42,242 |

| | | | | | | |
|---|---|---|---|---|---|---|
| L4 | 285 | -43<br>(-4,411 to 4,321) | 0.091<br>(0.080 to 0.102) | -474<br>(-48,457 to 48,740) | 89% | 63,017 |
| MI | 200 | 1,334<br>(-1,762 to 4,364) | 0.068<br>(0.058 to 0.080) | 19,525<br>(-25,258 to 66,318) | 67% | 29,086 |

**Table A23.** Model estimates of cost-effectiveness analyses associated with infant and maternal immunisation programs as standalone prevention strategies from healthcare and societal perspectives at the WTP of $30,000 per QALY gained, using constant vaccine efficacy profiles. All strategies were compared to the baseline with no intervention.

| Prevention strategy | Maximum PPD, $ | Incremental costs, $ (95% CI) | QALYs gained (95% CI) | ICER, $/QALY (95% CI) | Probability of being cost-effective | Budget impact per 100,000 population, $ |
|---|---|---|---|---|---|---|
| *Healthcare perspective* | | | | | | |
| L1 | 595 | 496<br>(-878 to 1,813) | 0.020<br>(0.014 to 0.027) | 24,389<br>(-41,470 to 100,863) | 56% | 532 |
| L2 | 370 | 754<br>(-783 to 2,253) | 0.031<br>(0.024 to 0.040) | 23,960<br>(-24,526 to 78,262) | 59% | 779 |
| L3 | 295 | 1,873<br>(-1,054 to 4,742) | 0.077<br>(0.066 to 0.088) | 24,335<br>(-13,546 to 64,696) | 61% | 1,929 |
| L4 | 215 | 2,681<br>(-927 to 6,227) | 0.091<br>(0.080 to 0.103) | 29,490<br>(-100,22 to 70,933) | 51% | 2,721 |
| MI | 180 | -1,045<br>(-3,914 to 1,802) | 0.073<br>(0.062 to 0.085) | -14,266<br>(-52,698 to 25,657) | 99% | -1,025 |
| *Societal perspective* | | | | | | |
| L1 | 685 | 502 | 0.020 | 24,697 | 56% | 3,237 |

| | | | | | | |
|---|---|---|---|---|---|---|
| | | (-922 to 1,859) | (0.014 to 0.027) | (-45,414 to 105,796) | | |
| L2 | 445 | 507<br>(-1,140 to 2,089) | 0.031<br>(0.024 to 0.040) | 16,153<br>(-35,575 to 73,124) | 69% | 7,010 |
| L3 | 380 | 2,240<br>(-1,077 to 5,484) | 0.077<br>(0.066 to 0.088) | 29,101<br>(-13,567 to 73,876) | 51% | 41,479 |
| L4 | 280 | -1,462<br>(-5,700 to 2,837) | 0.091<br>(0.080 to 0.103) | -16,076<br>(-62,970 to 31,800) | 97% | 60,598 |
| MI | 235 | 2,030<br>(-1,207 to 5,259) | 0.073<br>(0.063 to 0.085) | 27,715<br>(-15,848 to 74,388) | 54% | 35,679 |

**Table A24.** Model estimates of cost-effectiveness analyses associated with the combined infant and maternal immunisation program from healthcare and societal perspectives at the WTP of $30,000 per QALY gained, using sigmoidal vaccine efficacy profiles. All strategies were compared to the baseline with no intervention.

| Nirsevimab PPD, $ | RSVpreF PPD, $ | Incremental costs, $ (95% CI) | QALYs gained (95% CI) | ICER, $/QALY (95% CI) | Probability of being cost-effective | Budget impact per 100,000 population, $ |
|---|---|---|---|---|---|---|
| *Healthcare perspective* | | | | | | |
| 600 | 145 | 1,404<br>(-1,541 to 4,283) | 0.075<br>(0.064 to 0.087) | 18,649<br>(-20,449 to 59,587) | 72% | 1,430 |
| 215 | 160 | -153<br>(-3,053 to 2,727) | 0.075<br>(0.064 to 0.087) | -2,030<br>(-40,074 to 37,601) | 94% | -132 |
| *Societal perspective* | | | | | | |

| | | | | | | |
|---|---|---|---|---|---|---|
| 690 | 185 | 1,282<br>(-1,995 to 4,522) | 0.075<br>(0.064 to 0.087) | 17,036<br>(-25,638 to 62,765) | 72% | 30,828 |
| 285 | 200 | -901<br>(-4,135 to 2,324) | 0.075<br>(0.064 to 0.087) | -11,959<br>(-54,870 to 32,037) | 97% | 28,665 |

**Table A25.** Model estimates of cost-effectiveness analyses associated with the combined infant and maternal immunisation program from healthcare and societal perspectives at the WTP of $30,000 per QALY gained, using constant vaccine efficacy profiles. All strategies were compared to the baseline with no intervention.

| Nirsevimab PPD, $ | RSVpreF PPD, $ | Incremental costs, $ (95% CI) | QALYs gained (95% CI) | ICER, $/QALY (95% CI) | Probability of being cost-effective | Budget impact per 100,000 population, $ |
|---|---|---|---|---|---|---|
| *Healthcare perspective* | | | | | | |
| 595 | 170 | 2,074<br>(-942 to 5,089) | 0.079<br>(0.068 to 0.091) | 26,112<br>(-11,594 to 66,756) | 57% | 2,116 |
| 215 | 185 | 688<br>(-2,365 to 3,668) | 0.079<br>(0.068 to 0.091) | 8,660<br>(-29,622 to 48,272) | 86% | 705 |
| *Societal perspective* | | | | | | |
| 685 | 215 | -391<br>(-3,835 to 3,040) | 0.079<br>(0.068 to 0.091) | -4,921<br>(-47,341 to 39,482) | 94% | 34,851 |
| 280 | 235 | 783<br>(-2,624 to 4,289) | 0.079<br>(0.068 to 0.091) | 9,859<br>(-32,671 to 55,729) | 81% | 36,025 |

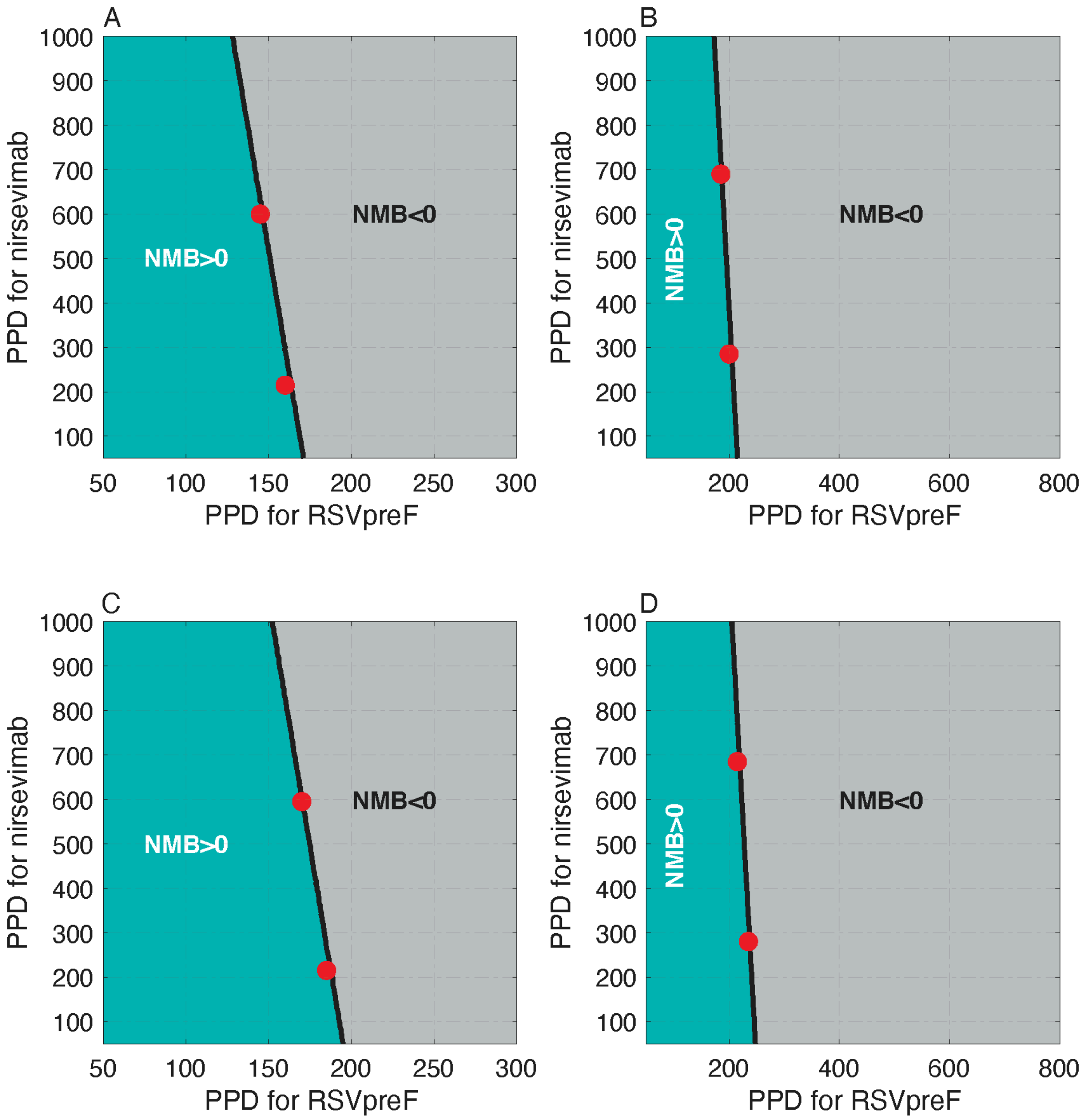

**Figure A17.** Net monetary benefit (NMB) of the combined infant and maternal immunisation program at the WTP of $30,000 per QALY gained as a function of PPD for nirsevimab and RSVpreF. Panels (A) and (B) correspond to the analysis from healthcare and societal perspectives, respectively, with sigmoidal vaccine efficacy profiles. Panels (C) and (D) correspond to the analysis from the healthcare and societal perspectives, respectively, with constant vaccine efficacy profiles. Red circles correspond to the PPD values in Tables A24 and A25.

###### 5.2.2. *Cost-effectiveness analysis with monetary loss of life*

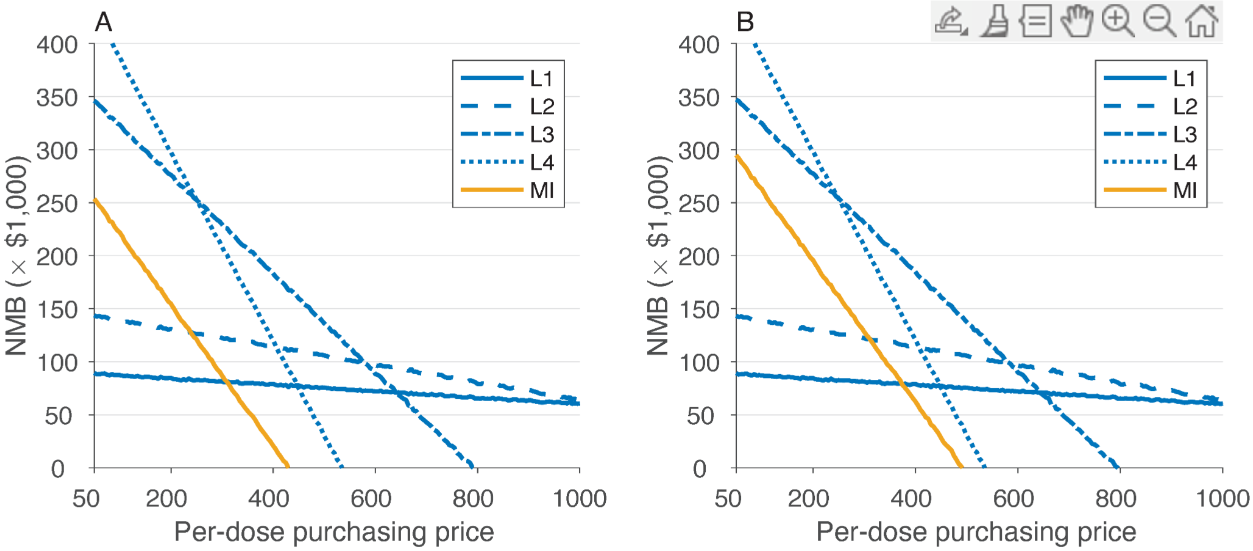

**Figure A18.** Estimated net monetary benefit (NMB) as a function of price per dose at the WTP threshold of $30,000 per QALY gained with the inclusion of monetary loss of life due to RSV-related infant mortality. Panels (A) and (B) correspond to the analysis from a societal perspective using sigmoidal and constant vaccine efficacy profiles, respectively. Note that the inclusion of monetary loss due to infant mortality (as an indirect cost) does not affect cost-effectiveness analysis from a healthcare perspective.

**Table A26.** Model estimates of cost-effectiveness analyses associated with infant and maternal immunisation programs as standalone prevention strategies from a societal perspective at the WTP of $30,000 per QALY gained. All strategies were compared to the baseline with no intervention, and with the inclusion of monetary loss of life due to RSV-related infant mortality. Since the monetary loss of life, as an indirect cost, does not affect cost-effectiveness from a healthcare perspective, the results of Table A22 and A23 of the previous section are applicable for the healthcare perspective.

| Prevention strategy | Maximum PPD, $ | Incremental costs, $ (95% CI) | QALYs gained (95% CI) | ICER, $/QALY (95% CI) | Probability of being cost-effective | Budget impact per 100,000 population, $ |
|---|---|---|---|---|---|---|
| *Societal perspective, sigmoidal vaccine efficacy profiles* | | | | | | |
| L1 | 1,000 | -33,700 (-54,217 to -15,243) | 0.881 (0.513 to 1.295) | -38,274 (-42,304 to -29,416) | Cost-saving | 12,601 |
| L2 | 1,000 | -22,491 (-48,607 to 1,060) | 1.390 (0.930 to 1.898) | -16,175 (-25,432 to 1,140) | 100% | 52,877 |
| L3 | 790 | 71,513 (38,525 to 102,736) | 2.432 (1.832 to 3.122) | 29,408 (12,477 to 56,626) | 52% | 230,689 |
| L4 | 530 | 80,634 (45,069 to 112,940) | 2.803 (2.163 to 3.499) | 28,722 (12,828 to 52,115) | 54% | 281,171 |
| MI | 430 | 58,616 (27,535 to 86,877) | 1.973 (1.417 to 2.572) | 29,772 (10,633 to 61,444) | 50% | 182,573 |
| *Societal perspective, constant vaccine efficacy profiles* | | | | | | |
| L1 | 1,000 | -33,560 (-54,101 to -15,073) | 0.880 (0.513 to 1.295) | -38,115 (-42,186 to -29,129) | Cost-saving | 12,705 |
| L2 | 1,000 | -22,153 (-48,236 to 1,425) | 1.390 (0.930 to 1.898) | -15,932 (-25,280 to 1,532) | 100% | 53,118 |
| L3 | 795 | 73,592 | 2.480 | 29,639 | 51% | 234,579 |

| | | | | | | |
|---|---|---|---|---|---|---|
| | | (40,529 to 104,645) | (1.878 to 3.124) | (12,894 to 56,205) | | |
| L4 | 530 | 81,418<br>(45,889 to 114,324) | 2.847<br>(2.208 to 3.544) | 28,543<br>(12,924 to 51,753) | 55% | 283,204 |
| MI | 490 | 64,379<br>(31,594 to 94,526) | 2.203<br>(1.602 to 2.849) | 29,208<br>(11,019 to 58,847) | 52% | 205,850 |

**Table A27.** Model estimates of cost-effectiveness analyses associated with the combined infant and maternal immunisation program from a societal perspective at the WTP of $30,000 per QALY gained. All strategies were compared to the baseline with no intervention, and with the inclusion of monetary loss of life due to RSV-related infant mortality. Since the monetary loss of life, as an indirect cost, does not affect cost-effectiveness from a healthcare perspective, the results of Table A24 and A25 of the previous section are applicable for the healthcare perspective.

| Nirsevimab PPD, $ | RSVpreF PPD, $ | Incremental costs, $ (95% CI) | QALYs gained (95% CI) | ICER, $/QALY (95% CI) | Probability of being cost-effective | Budget impact per 100,000 population, $ |
|---|---|---|---|---|---|---|
| *Societal perspective, sigmoidal vaccine efficacy profiles* | | | | | | |
| 1,000 | 435 | 67,379<br>(34,766 to 97,791) | 2.251<br>(1.651 to 2.895) | 29,953<br>(12,021 to 59,212) | 49% | 206,980 |
| 530 | 455 | 66,558<br>(34,166 to 96,844) | 2.251<br>(1.651 to 2.895) | 29,567<br>(11,793 to 58,652) | 51% | 206,200 |
| *Societal perspective, constant vaccine efficacy profiles* | | | | | | |
| 1,000 | 485 | 69,866<br>(36,677 to 102,388) | 2.438<br>(1.790 to 3.081) | 28,657<br>(11,850 to 57,103) | 54% | 224,500 |
| 530 | 510 | 72,319 | 2.438 | 29,631 | 51% | 227,057 |

| | | | | |
|---|---|---|---|---|
| | | (39,406 to 104,324) | (1.790 to 3.081) | (12,789 to 57,314) |

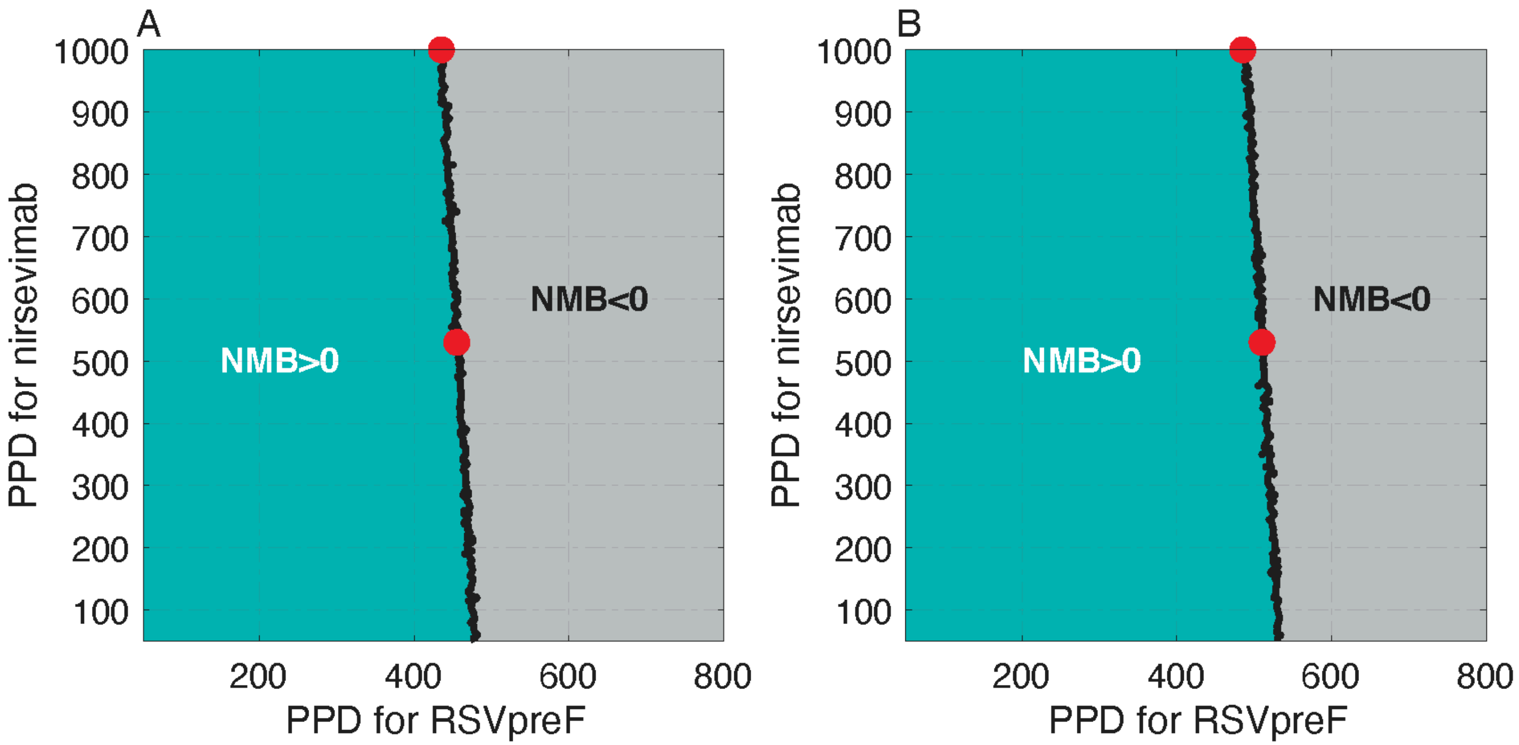

**Figure A19.** Net monetary benefit (NMB) of the combined infant and maternal immunisation program at the WTP of $30,000 per QALY gained as a function of PPD for nirsevimab and RSVpreF. Panels (A) and (B) correspond to the analysis from a societal perspective with sigmoidal and constant vaccine efficacy profiles, respectively. Red circles correspond to the PPD values in Table A27.

### 6. Secondary analyses with a WTP of $70,000 per QALY gained

#### 6.1. 100% coverage of nirsevimab and 100% coverage of maternal vaccination

The reductions of health outcomes for this scenario are the same as those reported in the main text with the WTP of $50,000 per QALY gained.

##### 6.1.1. *Cost-effectiveness analysis without monetary loss of life*

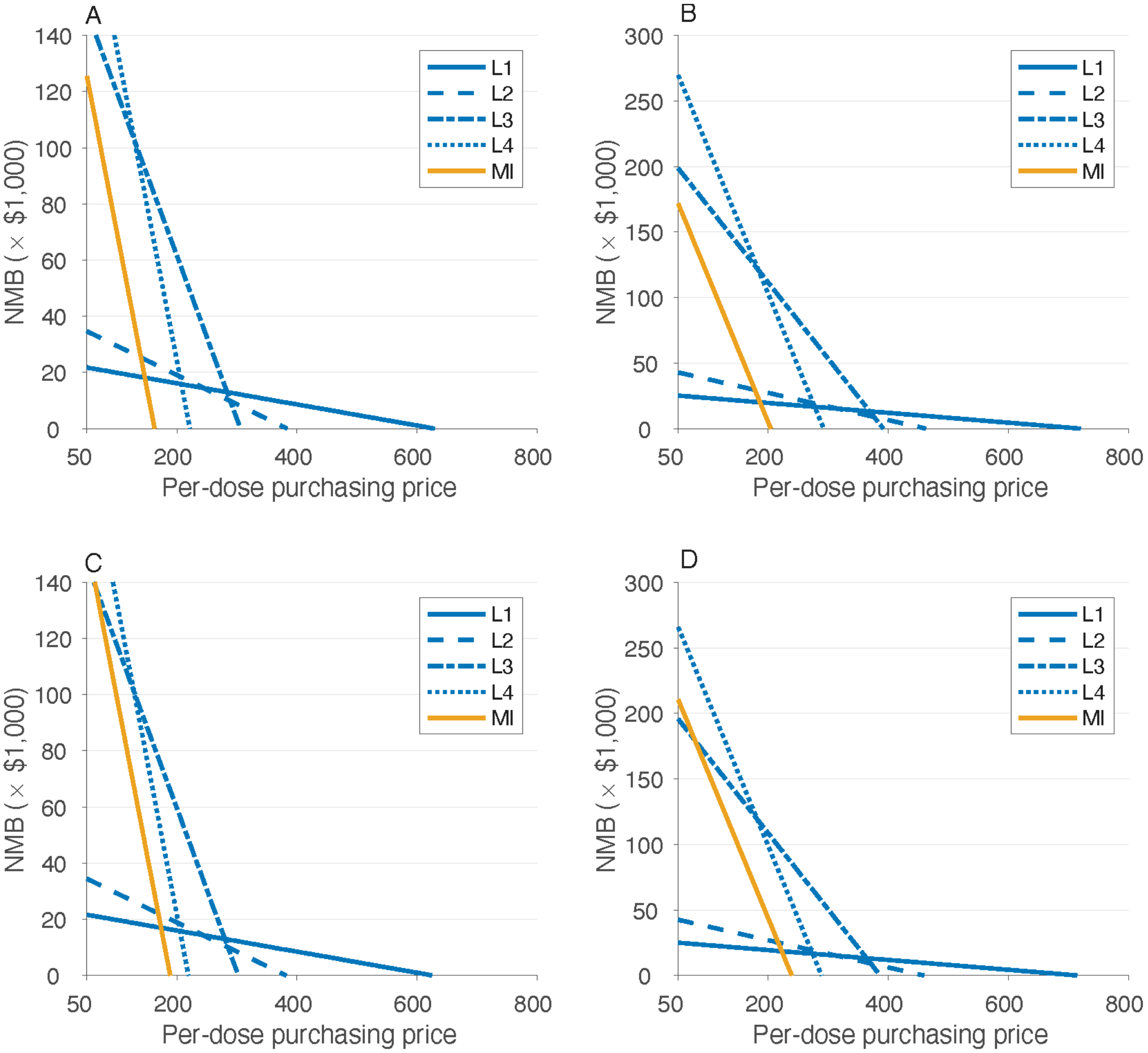

**Figure A20.** Estimates of the net monetary benefit (NMB) as a function of price per dose at the WTP threshold of $70,000 per QALY gained. Panels (A) and (B) correspond to the analysis from healthcare and societal perspectives, respectively, with sigmoidal vaccine efficacy profiles. Panels (C) and (D) correspond to the analysis from the healthcare and societal perspectives, respectively, with constant vaccine efficacy profiles.

**Table A28.** Model estimates of cost-effectiveness analyses associated with infant and maternal immunisation programs as standalone prevention strategies from healthcare and societal perspectives at the WTP of $70,000 per QALY gained, using sigmoidal vaccine efficacy profiles. All strategies were compared to the baseline with no intervention.

| Prevention strategy | Maximum PPD, $ | Incremental costs, $ (95% CI) | QALYs gained (95% CI) | ICER, $/QALY (95% CI) | Probability of being cost-effective | Budget impact per 100,000 population, $ |
|---|---|---|---|---|---|---|
| *Healthcare perspective* | | | | | | |
| L1 | 625 | 1,546 (27 to 3,022) | 0.024 (0.018 to 0.032) | 63,636 (10,22 to 144,239) | 57% | 1,601 |
| L2 | 380 | 2,151 (331 to 3,895) | 0.036 (0.028 to 0.045) | 60,007 (8,944 to 120,389) | 64% | 2,188 |
| L3 | 305 | 6,164 (2,795 to 9,563) | 0.094 (0.082 to 0.107) | 65,322 (28,701 to 106,456) | 60% | 6,211 |
| L4 | 220 | 6,083 (1,638 to 10,445) | 0.111 (0.099 to 0.124) | 54,742 (14,437 to 97,296) | 76% | 6,068 |
| MI | 160 | 4,501 (764 to 8,262) | 0.109 (0.096 to 0.123) | 41,321 (6,800 to 78,174) | 94% | 4,546 |
| *Societal perspective* | | | | | | |
| L1 | 715 | 1,543 (-92 to 3,116) | 0.024 (0.018 to 0.032) | 63,630 (-3,464 to 147,418) | 56% | 4,982 |
| L2 | 460 | 2,303 (402 to 4,176) | 0.036 (0.028 to 0.045) | 64,271 (10,477 to 128,675) | 57% | 10,495 |
| L3 | 390 | 5,694 (1,656 to 9,585) | 0.094 (0.082 to 0.107) | 60,317 (17,139 to 105,805) | 66% | 55,646 |

| | | | | | | |
|---|---|---|---|---|---|---|
| L4 | 290 | 5,195<br>(68 to 10,285) | 0.111<br>(0.099 to 0.124) | 46,749<br>(597 to 95,262) | 83% | 83,978 |
| MI | 200 | 2,816<br>(-1,495 to 7,037) | 0.109<br>(0.096 to 0.123) | 25,815<br>(-13,217 to 66,816) | 98% | 49,066 |

**Table A29.** Model estimates of cost-effectiveness analyses associated with infant and maternal immunisation programs as standalone prevention strategies from healthcare and societal perspectives at the WTP of $70,000 per QALY gained, using constant vaccine efficacy profiles. All strategies were compared to the baseline with no intervention.

| Prevention strategy | Maximum PPD, $ | Incremental costs, $ (95% CI) | QALYs gained (95% CI) | ICER, $/QALY (95% CI) | Probability of being cost-effective | Budget impact per 100,000 population, $ |
|---|---|---|---|---|---|---|
| *Healthcare perspective* | | | | | | |
| L1 | 625 | 1,652<br>(126 to 3,116) | 0.024<br>(0.018 to 0.031) | 68,144<br>(5,105 to 149,616) | 52% | 1,708 |
| L2 | 380 | 2,329<br>(516 to 4,063) | 0.036<br>(0.028 to 0.045) | 64,973<br>(1,3921 to 126,237) | 57% | 2,368 |
| L3 | 300 | 5,198<br>(1,802 to 8,538) | 0.094<br>(0.082 to 0.106) | 55,623<br>(18,734 to 95,404) | 77% | 5,262 |
| L4 | 215 | 3,017<br>(-1,327 to 7,221) | 0.110<br>(0.098 to 0.123) | 27,348<br>(-11,810 to 67,888) | 98% | 3,050 |
| MI | 185 | 3,991<br>(-45 to 8,032) | 0.117<br>(0.104 to 0.131) | 34,041<br>(-362 to 71,355) | 97% | 4,010 |
| *Societal perspective* | | | | | | |
| L1 | 715 | 1,694 | 0.024 | 69,993 | 49% | 5,089 |

| | | | | | | |
|---|---|---|---|---|---|---|
| | | (78 to 3,276) | (0.018 to 0.031) | (3,063 to 155,201) | | |
| L2 | 455 | 2,083<br>(180 to 3,894) | 0.036<br>(0.028 to 0.045) | 58,105<br>(4,851 to 120,342) | 66% | 10,157 |
| L3 | 385 | 5,602<br>(1,624 to 9,559) | 0.094<br>(0.082 to 0.106) | 59,775<br>(16,760 to 105,812) | 67% | 54,698 |
| L4 | 285 | 3,439<br>(-1,753 to 8,664) | 0.110<br>(0.098 to 0.123) | 31,187<br>(-15,679 to 80,821) | 94% | 80,960 |
| MI | 235 | 3,554<br>(-1,100 to 8,303) | 0.117<br>(0.104 to 0.131) | 30,317<br>(-9,310 to 73,014) | 96% | 59,660 |

**Table A30.** Model estimates of cost-effectiveness analyses associated with the combined infant and maternal immunisation program from healthcare and societal perspectives at the WTP of $70,000 per QALY gained, using sigmoidal vaccine efficacy profiles. All strategies were compared to the baseline with no intervention.

| Nirsevimab PPD, $ | RSVpreF PPD, $ | Incremental costs, $ (95% CI) | QALYs gained (95% CI) | ICER, $/QALY (95% CI) | Probability of being cost-effective | Budget impact per 100,000 population, $ |
|---|---|---|---|---|---|---|
| *Healthcare perspective* | | | | | | |
| 625 | 145 | 6,366<br>(2,515 to 10,154) | 0.112<br>(0.099 to 0.126) | 56,818<br>(21,806 to 94,480) | 76% | 6,408 |
| 220 | 155 | 2,228<br>(-1,658 to 6,080) | 0.112<br>(0.099 to 0.126) | 19,867<br>(-14,409 to 55,964) | 100% | 2,323 |
| *Societal perspective* | | | | | | |

| | | | | | | |
|---|---|---|---|---|---|---|
| 715 | 185 | 6,612<br>(2,225 to 11,049) | 0.112<br>(0.099 to 0.126) | 58,980<br>(19,448 to 103,015) | 70% | 54,309 |
| 290 | 200 | 7,300<br>(2,905 to 11,642) | 0.112<br>(0.099 to 0.126) | 65,090<br>(25,525 to 107,942) | 59% | 55,038 |

**Table A31.** Model estimates of cost-effectiveness analyses associated with the combined infant and maternal immunisation program from healthcare and societal perspectives at the WTP of $70,000 per QALY gained, using constant vaccine efficacy profiles. All strategies were compared to the baseline with no intervention.

| Nirsevimab PPD, $ | RSVpreF PPD, $ | Incremental costs, $ (95% CI) | QALYs gained (95% CI) | ICER, $/QALY (95% CI) | Probability of being cost-effective | Budget impact per 100,000 population, $ |
|---|---|---|---|---|---|---|
| *Healthcare perspective* | | | | | | |
| 625 | 170 | 8,183<br>(4,141 to 12,247) | 0.119<br>(0.106 to 0.133) | 68,943<br>(34,000 to 107,073) | 52% | 8,227 |
| 215 | 180 | 3,911<br>(-272 to 7,989) | 0.119<br>(0.106 to 0.133) | 32,932<br>(-2,311 to 69,494) | 98% | 3,954 |
| *Societal perspective* | | | | | | |
| 715 | 215 | 4,553<br>(-133 to 9,241) | 0.119<br>(0.106 to 0.133) | 38,344<br>(-1,120 to 80,203) | 93% | 61,693 |
| 285 | 230 | 5,083<br>(266 to 9,849) | 0.119<br>(0.106 to 0.133) | 42,805<br>(2,217 to 85,356) | 90% | 62,234 |

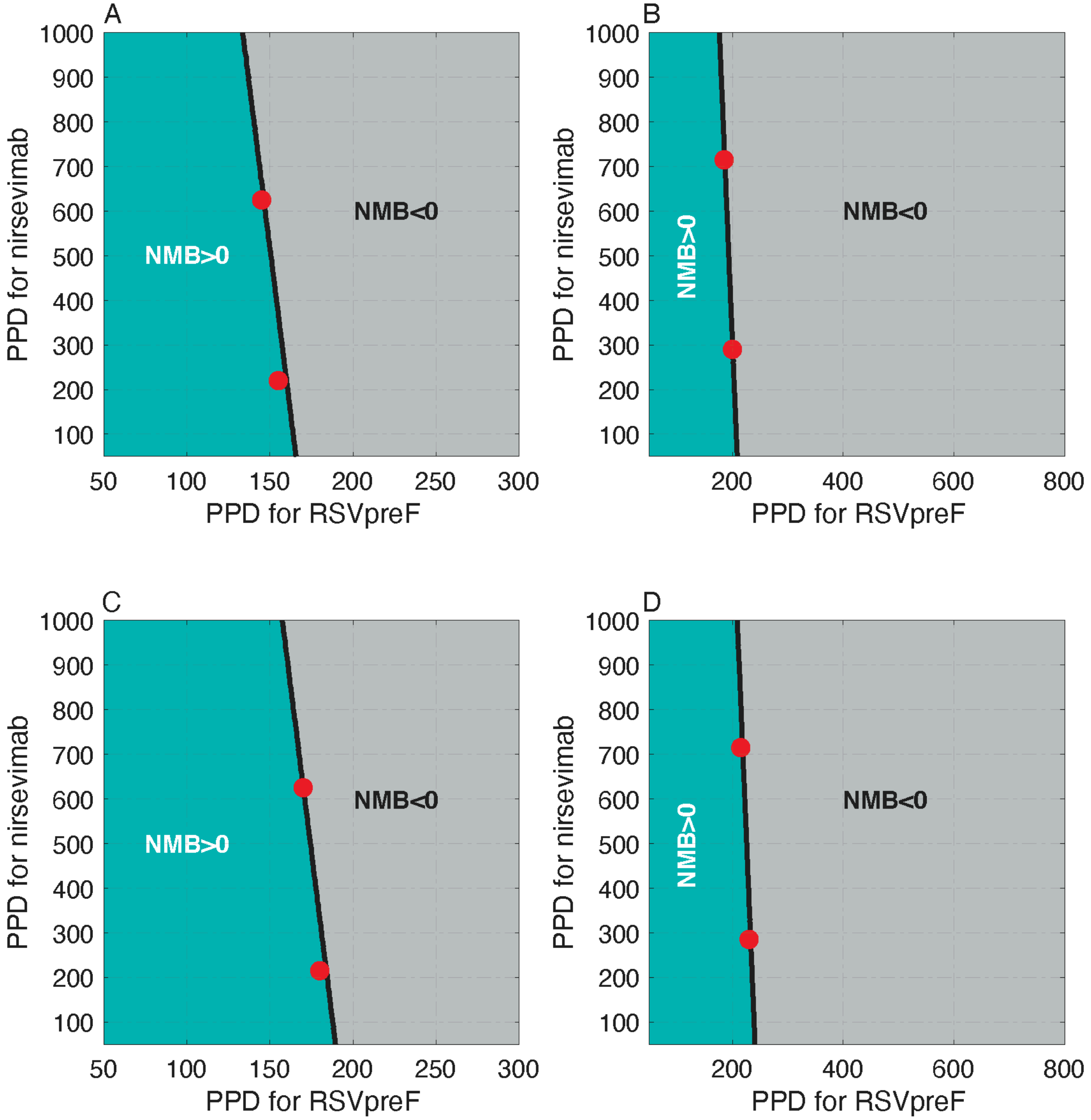

**Figure A21.** Net monetary benefit (NMB) of the combined infant and maternal immunisation program at the WTP of $70,000 per QALY gained as a function of PPD for nirsevimab and RSVpreF. Panels (A) and (B) correspond to the analysis from healthcare and societal perspectives, respectively, with sigmoidal vaccine efficacy profiles. Panels (C) and (D) correspond to the analysis from the healthcare and societal perspectives, respectively, with constant vaccine efficacy profiles. Red circles correspond to the PPD values in Tables A30 and A31.

###### 6.1.2. *Cost-effectiveness analysis with monetary loss of life*

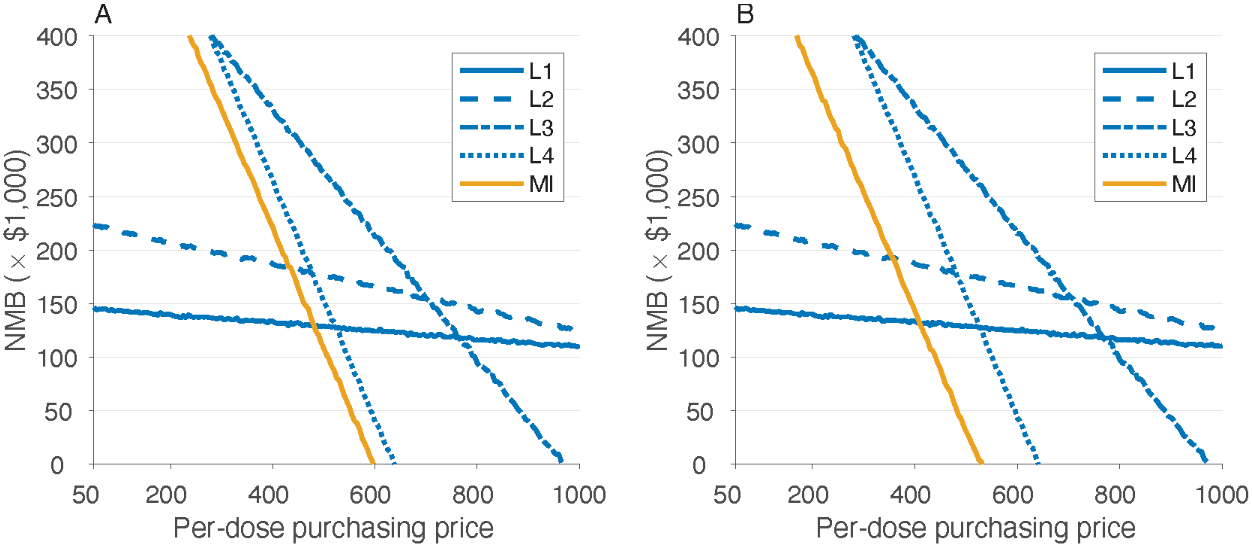

**Figure A22.** Estimated net monetary benefit (NMB) as a function of price per dose at the WTP threshold of $70,000 per QALY gained with the inclusion of monetary loss of life due to RSV-related infant mortality. Panels (A) and (B) correspond to the analysis from a societal perspective using sigmoidal and constant vaccine efficacy profiles, respectively. Note that the inclusion of monetary loss due to infant mortality (as an indirect cost) does not affect cost-effectiveness analysis from a healthcare perspective.

**Table A32.** Model estimates of cost-effectiveness analyses associated with LAMA and maternal immunisation programs as standalone prevention strategies from a societal perspective at the WTP of $70,000 per QALY gained. All strategies were compared to the baseline with no intervention, and with the inclusion of monetary loss of life due to RSV-related infant mortality. Since the monetary loss of life, as an indirect cost, does not affect cost-effectiveness from a healthcare perspective, the results of Table A28 and A29 are applicable for the healthcare perspective.

| Prevention strategy | Maximum PPD, $ | Incremental costs, $ (95% CI) | QALYs gained (95% CI) | ICER, $/QALY (95% CI) | Probability of being cost-effective | Budget impact per 100,000 population, $ |
|---|---|---|---|---|---|---|
| *Societal perspective, sigmoidal vaccine efficacy profiles* | | | | | | |
| L1 | 1,000 | -38,282<br>(-60,190 to -17,798) | 1.023<br>(0.607 to 1.479) | -37,439<br>(-41,487 to -28,904) | Cost-saving | 15,688 |
| L2 | 1,000 | -17,393<br>(-44,625 to 7,446) | 1.533<br>(1.025 to 2.083) | -11,345<br>(-21,523 to 7,164) | 100% | 66,571 |
| L3 | 975 | 203,472<br>(168,305 to 237,559) | 2.904<br>(2.256 to 3.595) | 69,951<br>(46,591 to 106,528) | 49% | 395,878 |
| L4 | 640 | 231,840<br>(194,127 to 268,248) | 3.324<br>(2.635 to 4.065) | 6,9641<br>(47,771 to 102,355) | 50% | 473,528 |
| MI | 525 | 213,258<br>(175,637 to 249,344) | 3.101<br>(2.406 to 3.882) | 68,850<br>(45,767 to 104,191) | 52% | 410,791 |
| *Societal perspective, constant vaccine efficacy profiles* | | | | | | |
| L1 | 1,000 | -38,129<br>(-60,033 to -17,630) | 1.022<br>(0.607 to 1.479) | -37,291<br>(-41,376 to -28,707) | Cost-saving | 15,795 |
| L2 | 1,000 | -17,089<br>(-44,330 to 7,796) | 1.533<br>(1.025 to 2.083) | -11,147<br>(-21,381 to 7,553) | 100% | 66,752 |
| L3 | 965 | 201,094 | 2.895 | 69,413 | 51% | 392,022 |

| | | | | | | |
|---|---|---|---|---|---|---|
| | | (165,494 to 234,697) | (2.212 to 3.595) | (45,971 to 104,848) | | |
| L4 | 635 | 230,156<br>(192,093 to 266,222) | 3.327<br>(2.635 to 4.064) | 69,170<br>(47,216 to 101,519) | 52% | 470,510 |
| MI | 595 | 237,119<br>(197,394 to 274,121) | 3.419<br>(2.684 to 4.209) | 69,340<br>(46,968 to 101,859) | 51% | 460,340 |

**Table A33.** Model estimates of cost-effectiveness analyses associated with the combined infant and maternal immunisation program from a societal perspective at the WTP of $70,000 per QALY gained. All strategies were compared to the baseline with no intervention, and with the inclusion of monetary loss of life due to RSV-related infant mortality. Since the monetary loss of life, as an indirect cost, does not affect cost-effectiveness from a healthcare perspective, the results of Table A30 and A31 are applicable for the healthcare perspective.

| Nirsevimab PPD, $ | RSVpreF PPD, $ | Incremental costs, $ (95% CI) | QALYs gained (95% CI) | ICER, $/QALY (95% CI) | Probability of being cost-effective | Budget impact per 100,000 population, $ |
|---|---|---|---|---|---|---|
| *Societal perspective, sigmoidal vaccine efficacy profiles* | | | | | | |
| 1,000 | 520 | 229,702<br>(190,522 to 267,562) | 3.282<br>(2.589 to 4.023) | 69,979<br>(46,880 to 105,196) | 49% | 437,870 |
| 640 | 530 | 227,291<br>(189,396 to 263,542) | 3.282<br>(2.589 to 4.023) | 69,229<br>(46,830 to 102,719) | 51% | 435,476 |
| *Societal perspective, constant vaccine efficacy profiles* | | | | | | |
| 1,000 | 570 | 243,784<br>(203,966 to 281,373) | 3.526<br>(2.778 to 4.344) | 69,257<br>(474,53 to 101,422) | 52% | 467,515 |
| 635 | 580 | 241,282<br>(201,386 to 279,445) | 3.526<br>(2.778 to 4.344) | 68,552<br>(46,789 to 100,640) | 53% | 464,933 |

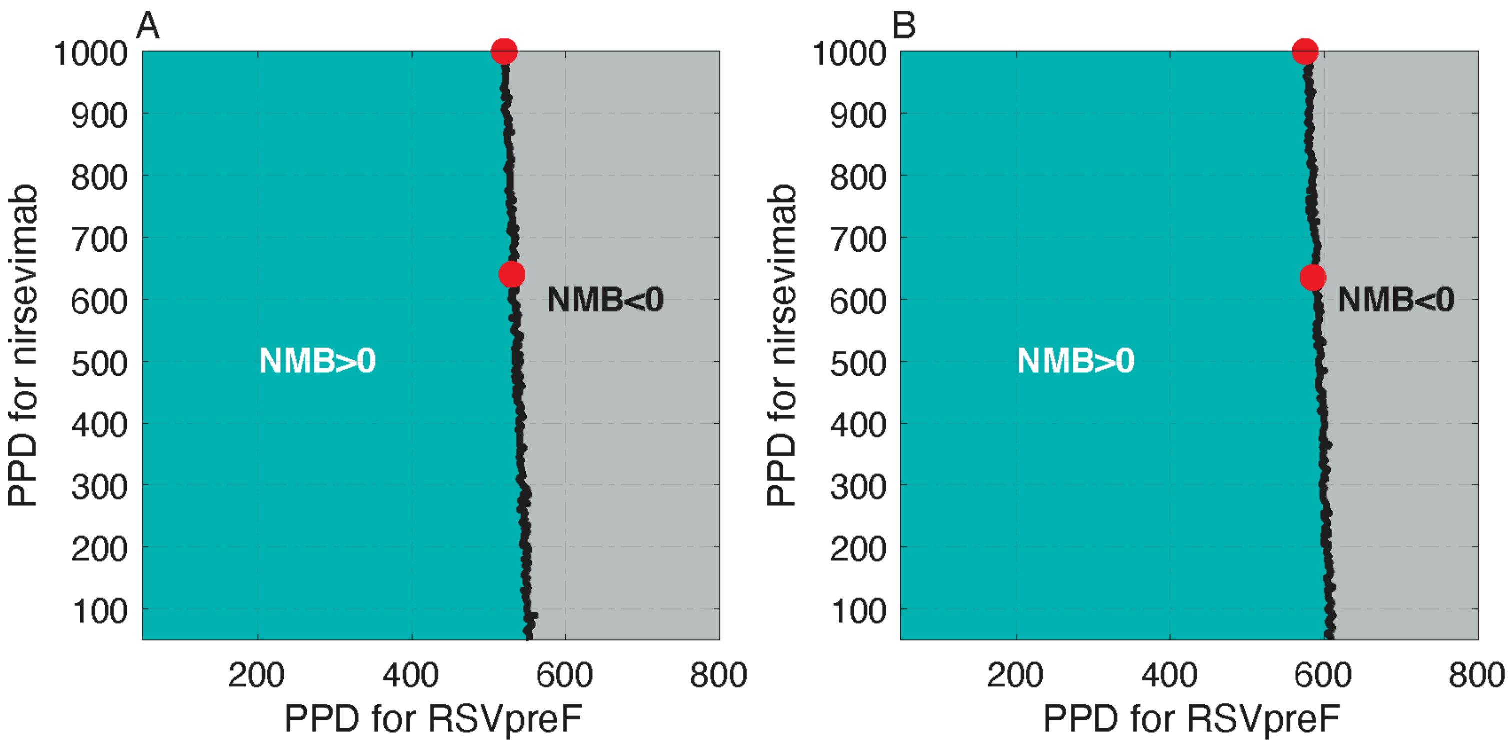

**Figure A23.** Net monetary benefit (NMB) of the combined infant and maternal immunisation program at the WTP of $70,000 per QALY gained as a function of PPD for nirsevimab and RSVpreF. Panels (A) and (B) correspond to the analysis from a societal perspective with sigmoidal and constant vaccine efficacy profiles, respectively. Red circles correspond to the PPD values in Table A33.

##### 6.2. 80% coverage of nirsevimab and 60% coverage of maternal vaccination

Reduction of health outcomes are the same as presented in section 3.2 of this Supplemental.

###### 6.2.1. *Cost-effectiveness analysis without monetary loss of life*

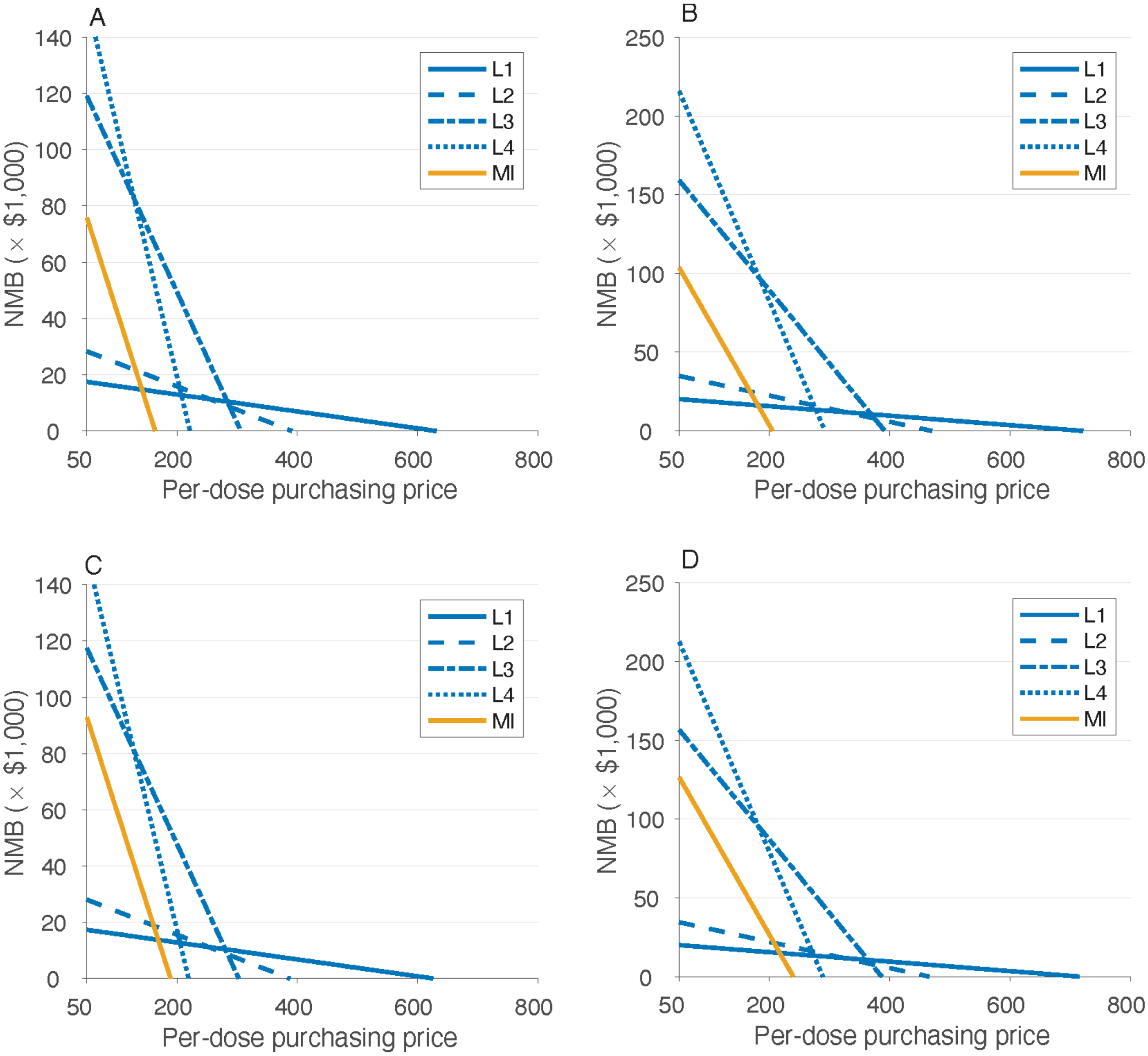

**Figure A24.** Estimates of the net monetary benefit (NMB) as a function of price per dose at the WTP threshold of $70,000 per QALY gained. Panels (A) and (B) correspond to the analysis from healthcare and societal perspectives, respectively, with sigmoidal vaccine efficacy profiles. Panels (C) and (D) correspond to the analysis from the healthcare and societal perspectives, respectively, with constant vaccine efficacy profiles.

**Table A34.** Model estimates of cost-effectiveness analyses associated with infant and maternal immunisation programs as standalone prevention strategies from healthcare and societal perspectives at the WTP of $70,000 per QALY gained, using sigmoidal vaccine efficacy profiles. All strategies were compared to the baseline with no intervention.

| Prevention strategy | Maximum PPD, $ | Incremental costs, $ (95% CI) | QALYs gained (95% CI) | ICER, $/QALY (95% CI) | Probability of being cost-effective | Budget impact per 100,000 population, $ |
|---|---|---|---|---|---|---|
| *Healthcare perspective* | | | | | | |
| L1 | 625 | 1,279 (-61 to 2,589) | 0.020 (0.014 to 0.027) | 62,523 (-28,11 to 151,206) | 58% | 1,180 |
| L2 | 390 | 2,189 (680 to 3,695) | 0.031 (0.024 to 0.040) | 69,603 (20,526 to 132,935) | 50% | 2,199 |
| L3 | 305 | 4,967 (2,109 to 7,798) | 0.077 (0.066 to 0.088) | 64,557 (26,778 to 106,680) | 60% | 5,018 |
| L4 | 220 | 5,143 (1,415 to 8,756) | 0.091 (0.080 to 0.103) | 56,623 (15,056 to 100,241) | 73% | 5,139 |
| MI | 160 | 2,389 (-318 to 5,013) | 0.068 (0.058 to 0.080) | 34,976 (-4,485 to 77,612) | 95% | 2,393 |
| *Societal perspective* | | | | | | |
| L1 | 720 | 1,418 (12 to 2,816) | 0.020 (0.014 to 0.028) | 69,543 (490 to 160,533) | 50% | 4,185 |
| L2 | 465 | 1,847 (179 to 3,473) | 0.031 (0.024 to 0.040) | 58,736 (5,311 to 122,968) | 65% | 8,430 |
| L3 | 390 | 4,584 (1,279 to 7,824) | 0.077 (0.066 to 0.089) | 59,548 (16,162 to 105,915) | 67% | 44,569 |

| | | | | | | |
|---|---|---|---|---|---|---|
| L4 | 290 | 4,424<br>(215 to 8,673) | 0.091<br>(0.080 to 0.102) | 48,691<br>(2,268 to 98,420) | 81% | 67,469 |
| MI | 205 | 4,667<br>(1,667 to 7,664) | 0.068<br>(0.058 to 0.080) | 68,223<br>(23,401 to 117,995) | 52% | 32,423 |

**Table A35.** Model estimates of cost-effectiveness analyses associated with infant and maternal immunisation programs as standalone prevention strategies from healthcare and societal perspectives at the WTP of $70,000 per QALY gained, using constant vaccine efficacy profiles. All strategies were compared to the baseline with no intervention.

| Prevention strategy | Maximum PPD, $ | Incremental costs, $ (95% CI) | QALYs gained (95% CI) | ICER, $/QALY (95% CI) | Probability of being cost-effective | Budget impact per 100,000 population, $ |
|---|---|---|---|---|---|---|
| *Healthcare perspective* | | | | | | |
| L1 | 625 | 1,381<br>(40 to 2,687) | 0.020<br>(0.014 to 0.027) | 67,634<br>(1,794 to 156,827) | 53% | 1,433 |
| L2 | 385 | 1,979<br>(443 to 3,474) | 0.031<br>(0.024 to 0.040) | 62,837<br>(13,542 to 122,251) | 60% | 2,025 |
| L3 | 300 | 4,181<br>(1,285 to 6,997) | 0.077<br>(0.066 to 0.088) | 54,338<br>(16,468 to 95,551) | 78% | 4,255 |
| L4 | 215 | 2,681<br>(-927 to 6,227) | 0.091<br>(0.080 to 0.103) | 29,490<br>(-10,022 to 70,933) | 97% | 2,721 |
| MI | 185 | 2,309<br>(-599 to 5,097) | 0.073<br>(0.062 to 0.085) | 31,544<br>(-7,883 to 73,556) | 97% | 2,312 |
| *Societal perspective* | | | | | | |
| L1 | 715 | 1,412 | 0.020 | 69,236 | 50% | 4,138 |

| | | | | | | |
|---|---|---|---|---|---|---|
| | | (-39 to 2,792) | (0.014 to 0.027) | (-1,843 to 160,493) | | |
| L2 | 465 | 2,187<br>(517 to 3,820) | 0.031<br>(0.024 to 0.040) | 69,620<br>(15,408 to 135,143) | 50% | 8,671 |
| L3 | 385 | 4,525<br>(1,139 to 7,852) | 0.077<br>(0.066 to 0.088) | 58,640<br>(14,514 to 106,013) | 69% | 43,806 |
| L4 | 285 | 3,011<br>(-1,284 to 7,328) | 0.091<br>(0.080 to 0.103) | 33,127<br>(-13,956 to 83,476) | 93% | 65,050 |
| MI | 235 | 2,030<br>(-1,207 to 5,259) | 0.073<br>(0.063 to 0.085) | 27,715<br>(-15,848 to 74,388) | 96% | 35,679 |

**Table A36.** Model estimates of cost-effectiveness analyses associated with the combined infant and maternal immunisation program from healthcare and societal perspectives at the WTP of $70,000 per QALY gained, using sigmoidal vaccine efficacy profiles. All strategies were compared to the baseline with no intervention.

| Nirsevimab PPD, $ | RSVpreF PPD, $ | Incremental costs, $ (95% CI) | QALYs gained (95% CI) | ICER, $/QALY (95% CI) | Probability of being cost-effective | Budget impact per 100,000 population, $ |
|---|---|---|---|---|---|---|
| *Healthcare perspective* | | | | | | |
| 625 | 145 | 2,155<br>(-789 to 5,034) | 0.075<br>(0.064 to 0.087) | 28,634<br>(-10,255 to 70,030) | 98% | 2,181 |
| 220 | 165 | 3,323<br>(370 to 6,225) | 0.075<br>(0.064 to 0.087) | 44,207<br>(4,742 to 86,554) | 89% | 3,355 |
| *Societal perspective* | | | | | | |

| | | | | | | |
|---|---|---|---|---|---|---|
| 720 | 185 | 2,184<br>(-1,090 to 5,426) | 0.075<br>(0.064 to 0.087) | 29,020<br>(-14,136 to 75,183) | 96% | 31,730 |
| 290 | 205 | 2,582<br>(-650 to 5,822) | 0.075<br>(0.064 to 0.087) | 34,267<br>(-8,500 to 81,059) | 94% | 32,152 |

**Table A37.** Model estimates of cost-effectiveness analyses associated with the combined infant and maternal immunisation program from healthcare and societal perspectives at the WTP of $70,000 per QALY gained, using constant vaccine efficacy profiles. All strategies were compared to the baseline with no intervention.

| Nirsevimab PPD, $ | RSVpreF PPD, $ | Incremental costs, $ (95% CI) | QALYs gained (95% CI) | ICER, $/QALY (95% CI) | Probability of being cost-effective | Budget impact per 100,000 population, $ |
|---|---|---|---|---|---|---|
| *Healthcare perspective* | | | | | | |
| 625 | 170 | 2,975<br>(-40 to 5,991) | 0.079<br>(0.068 to 0.091) | 37,468<br>(-537 to 78,834) | 94% | 3,018 |
| 215 | 190 | 4,022<br>(1,011 to 7,148) | 0.079<br>(0.068 to 0.091) | 50,665<br>(12,098 to 94,734) | 82% | 4,041 |
| *Societal perspective* | | | | | | |
| 715 | 220 | 3,896<br>(359 to 7,351) | 0.079<br>(0.068 to 0.091) | 49,051<br>(43,26 to 96,418) | 81% | 39,090 |
| 285 | 240 | 4,283<br>(745 to 7,673) | 0.079<br>(0.068 to 0.091) | 53,929<br>(9,035 to 101,768) | 76% | 39,512 |

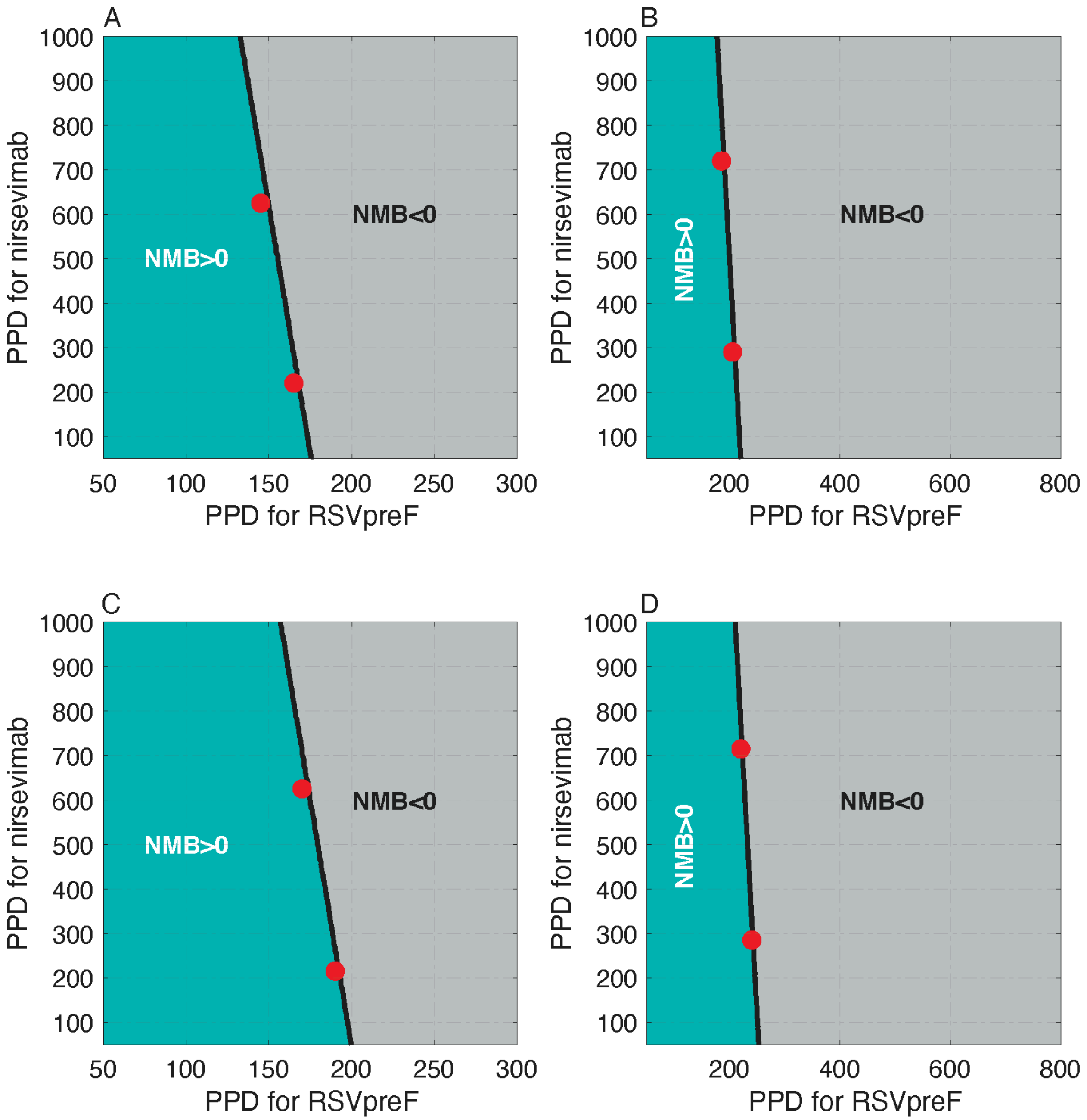

**Figure A25.** Net monetary benefit (NMB) of the combined infant and maternal immunisation program at the WTP of $70,000 per QALY gained as a function of PPD for nirsevimab and RSVpreF. Panels (A) and (B) correspond to the analysis from healthcare and societal perspectives, respectively, with sigmoidal vaccine efficacy profiles. Panels (C) and (D) correspond to the analysis from the healthcare and societal perspectives, respectively, with constant vaccine efficacy profiles. Red circles correspond to the PPD values in Tables A36 and A37.

##### 6.2.2. *Cost-effectiveness analysis with monetary loss of life*

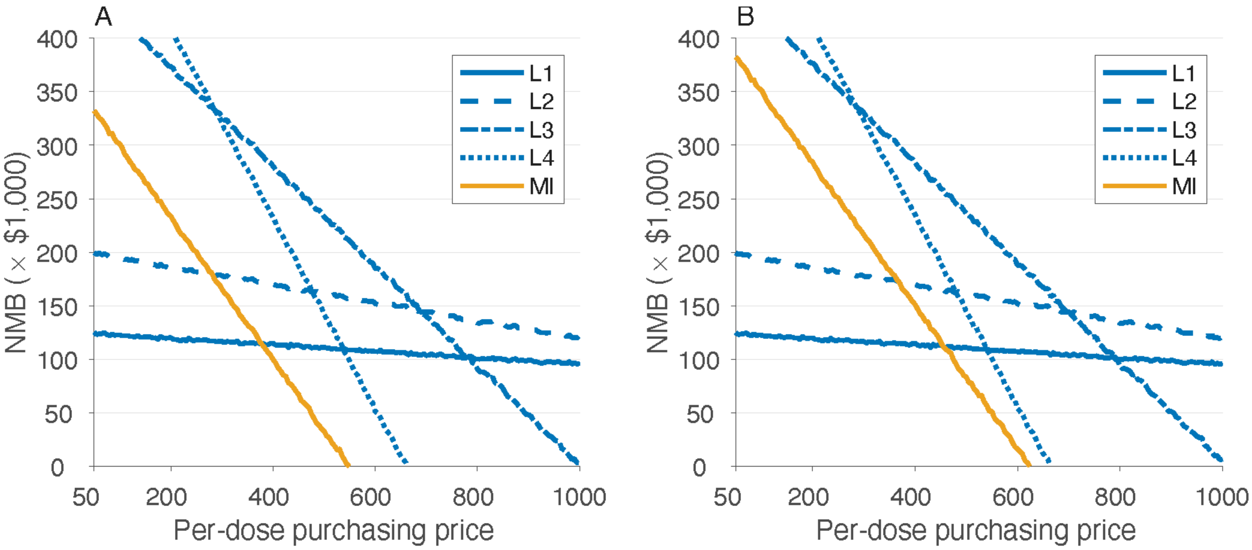

**Figure A26.** Estimated net monetary benefit (NMB) as a function of price per dose at the WTP threshold of $70,000 per QALY gained with the inclusion of monetary loss of life due to RSV-related infant mortality. Panels (A) and (B) correspond to the analysis from a societal perspective using sigmoidal and constant vaccine efficacy profiles, respectively. Note that the inclusion of monetary loss due to infant mortality (as an indirect cost) does not affect cost-effectiveness analysis from a healthcare perspective.

**Table A38.** Model estimates of cost-effectiveness analyses associated with infant and maternal immunisation programs as standalone prevention strategies from a societal perspective at the WTP of $70,000 per QALY gained. All strategies were compared to the baseline with no intervention with the inclusion of monetary loss of life due to RSV-related infant mortality. Since the monetary loss of life, as an indirect cost, does not affect cost-effectiveness from a healthcare perspective, the results of Table A34 and A35 are applicable for the healthcare perspective.

| Prevention strategy | Maximum PPD, $ | Incremental costs, $ (95% CI) | QALYs gained (95% CI) | ICER, $/QALY (95% CI) | Probability of being cost-effective | Budget impact per 100,000 population, $ |
|---|---|---|---|---|---|---|
| *Societal perspective, sigmoidal vaccine efficacy profiles* | | | | | | |
| L1 | 1,000 | -33,700 (-54,217 to -15,243) | 0.881 (0.513 to 1.295) | -38,274 (-42,304 to -29,416) | Cost-saving | 12,601 |
| L2 | 1,000 | -22,491 (-48,607 to 1,060) | 1.390 (0.930 to 1.898) | -16,175 (-25,432 to 1,140) | 100% | 52,877 |
| L3 | 1,000 | 169,134 (136,177 to 200,153) | 2.432 (1.832 to 3.122) | 69,504 (44,069 to 109,710) | 50% | 328,402 |
| L4 | 660 | 196,246 (161,825 to 229,689) | 2.803 (2.163 to 3.499) | 69,847 (46,405 to 107,012) | 50% | 396,926 |
| MI | 545 | 135,497 (105,141 to 163,923) | 1.973 (1.417 to 2.572) | 68,899 (40,795 to 116,021) | 52% | 292,684 |
| *Societal perspective, constant vaccine efficacy profiles* | | | | | | |
| L1 | 1,000 | -33,560 (-54,101 to -15,073) | 0.880 (0.513 to 1.295) | -38,115 (-42,186 to -29,129) | Cost-saving | 12,705 |
| L2 | 1,000 | -22,153 (-48,236 to 1,425) | 1.390 (0.930 to 1.898) | -15,932 (-25,280 to 1,532) | 100% | 53,118 |
| L3 | 1,000 | 169,177 | 2.480 | 68,257 | 53% | 329,966 |

| | | | | | | |
|---|---|---|---|---|---|---|
| | | (135,932 to 200,388) | (1.878 to 3.124) | (43,319 to 107,606) | | |
| L4 | 660 | 196,973<br>(162,364 to 231,073) | 2.856<br>(2.186 to 3.545) | 68,972<br>(45,842 to 105,412) | 52% | 398,960 |
| MI | 620 | 151,336<br>(119,116 to 181,867) | 2.203<br>(1.602 to 2.849) | 68,775<br>(42,031 to 113,753) | 52% | 275,920 |

**Table A39.** Model estimates of cost-effectiveness analyses associated with the combined infant and maternal immunisation program from a societal perspective at the WTP of $70,000 per QALY gained. All strategies were compared to the baseline with no intervention with the inclusion of monetary loss of life due to RSV-related infant mortality. Since the monetary loss of life, as an indirect cost, does not affect cost-effectiveness from a healthcare perspective, the results of Table A36 and A37 are applicable for the healthcare perspective.

| Nirsevimab PPD, $ | RSVpreF PPD, $ | Incremental costs, $ (95% CI) | QALYs gained (95% CI) | ICER, $/QALY (95% CI) | Probability of being cost-effective | Budget impact per 100,000 population, $ |
|---|---|---|---|---|---|---|
| *Societal perspective, sigmoidal vaccine efficacy profiles* | | | | | | |
| 1,000 | 565 | 153,896<br>(121,674 to 183,664) | 2.255<br>(1.650 to 2.895) | 68,260<br>(42,156 to 111,078) | 53% | 293,734 |
| 660 | 585 | 157255 124816<br>187622 | 2.255<br>(1.650 to 2.895) | 69887 43093<br>113721 | 49% | 296,861 |
| *Societal perspective, constant vaccine efficacy profiles* | | | | | | |
| 1,000 | 635 | 170,187<br>(136,513 to 200,934) | 2.438<br>(1.835 to 3.126) | 69,916<br>(44,134 to 109,821) | 50% | 324,601 |
| 660 | 650 | 169,798<br>(135,507 to 200,892) | 2.438<br>(1.835 to 3.126) | 69,645<br>(43,335 to 110,023) | 50% | 324,391 |

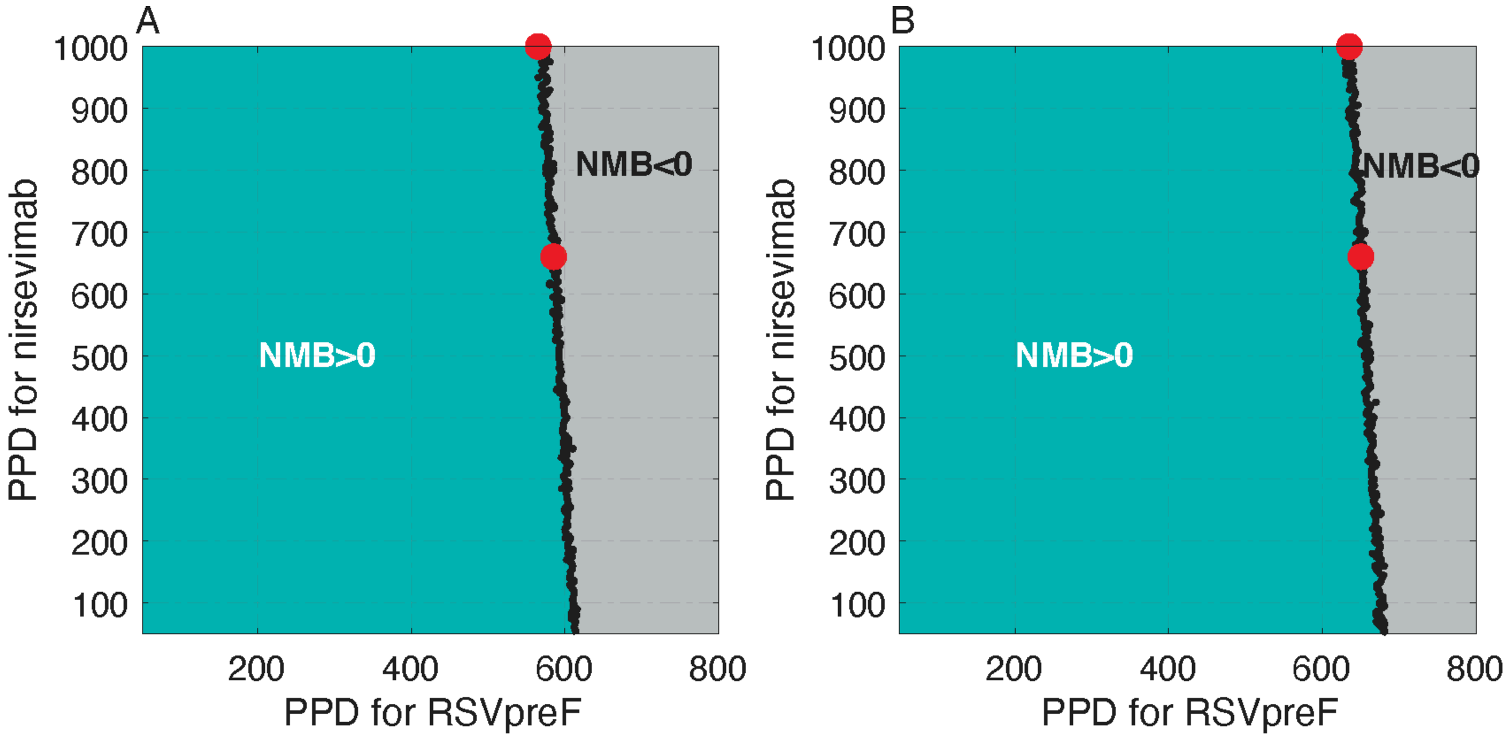

**Figure A27.** Net monetary benefit (NMB) of the combined infant and maternal immunisation program at the WTP of $70,000 per QALY gained as a function of PPD for nirsevimab and RSVpreF. Panels (A) and (B) correspond to the analysis from a societal perspective with sigmoidal and constant vaccine efficacy profiles, respectively. Red circles correspond to the PPD values in Table A39.

#### 7. Cost-effectiveness planes for immunisation programs with the WTP of $50,000 per QALY gained

##### 7.1. 100% coverage of nirsevimab and 100% coverage of maternal vaccination without monetary loss of life

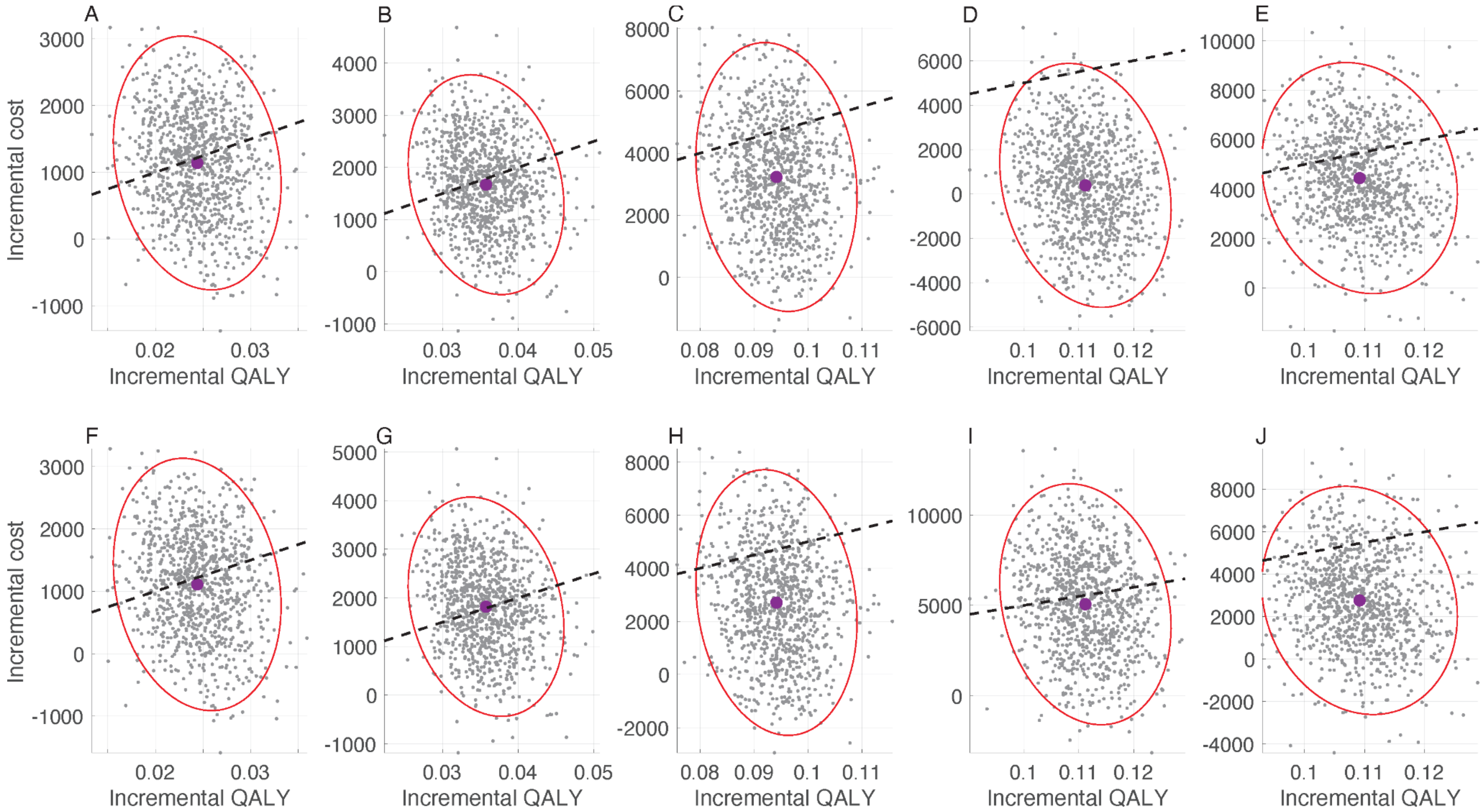

**Figure A28.** Cost-effectiveness planes for standalone immunisation programs: (A,F) L1; (B,G) L2; (C,H) L3; (D,I) L4, and (E,J) MI from healthcare (A,B,C,D,E) and societal (F,G,H,I,J) perspectives using sigmoidal vaccine efficacy profiles. The maximum PPD for

nirsevimab and RSVpreF correspond to estimates reported in Table 3 of the main text. Black dashed-line corresponds to the WTP threshold of $50,000 per QALY gained.

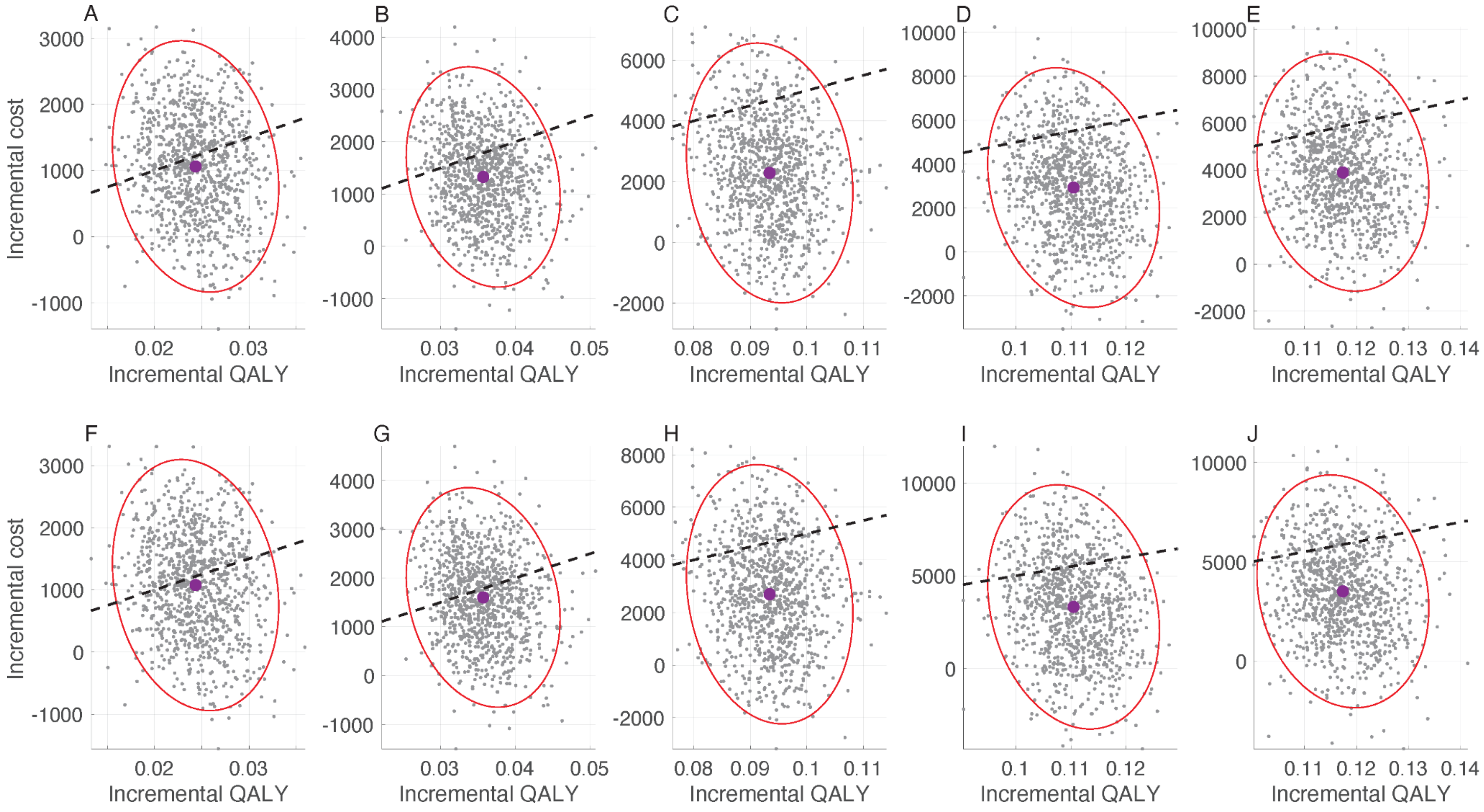

**Figure A29.** Cost-effectiveness planes for standalone immunisation programs: (A,F) L1; (B,G) L2; (C,H) L3; (D,I) L4, and (E,J) MI from healthcare (A,B,C,D,E) and societal (F,G,H,I,J) perspectives using constant vaccine efficacy profiles. The maximum PPD for nirsevimab and RSVpreF correspond to estimates reported in Table 4 of the main text. Black dashed-line corresponds to the WTP threshold of $50,000 per QALY gained.

##### 7.2. 100% coverage of nirsevimab and 100% coverage of maternal vaccination with monetary loss of life

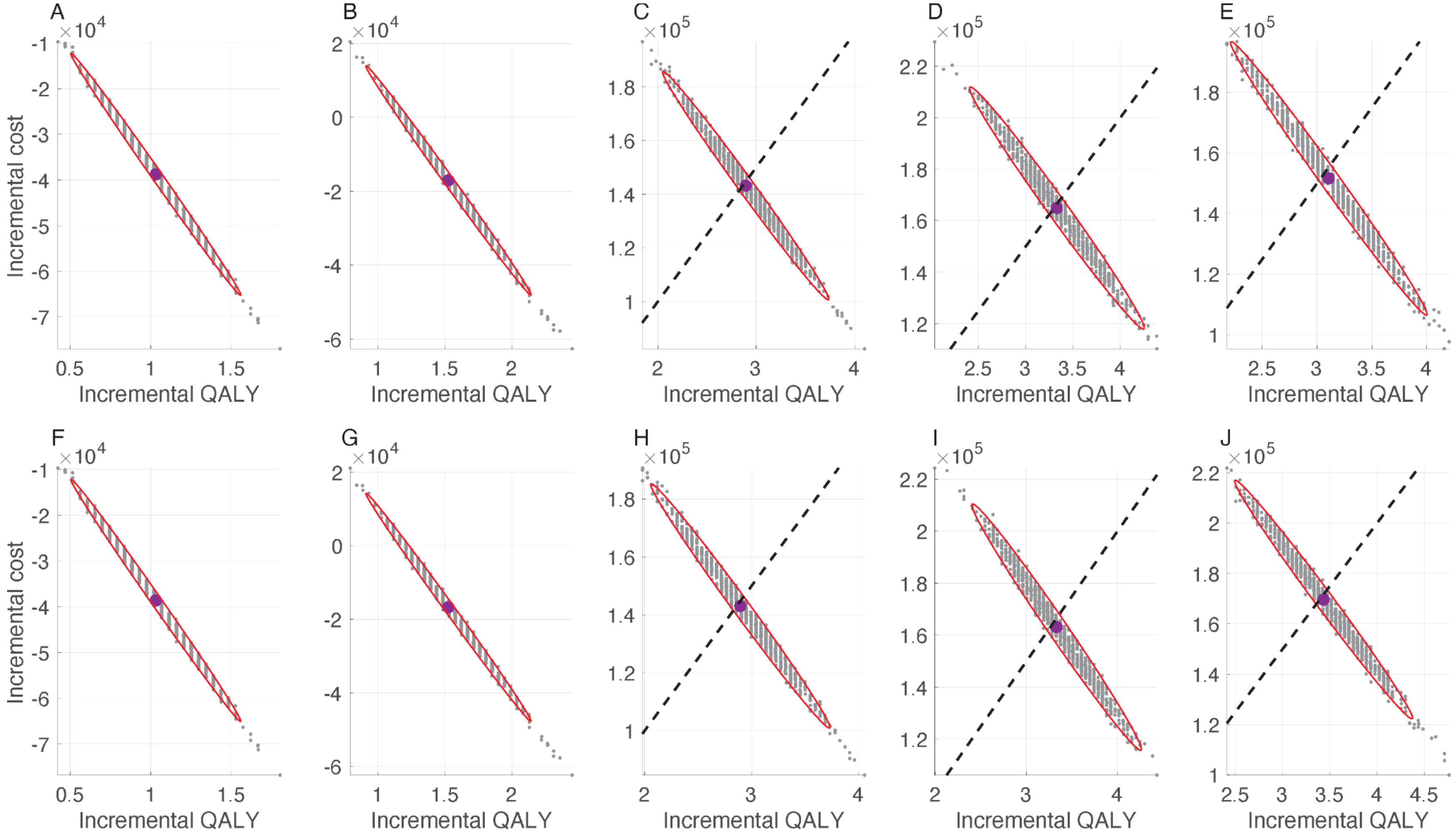

**Figure A30.** Cost-effectiveness planes for standalone immunisation programs: (A,F) L1; (B,G) L2; (C,H) L3; (D,I) L4, and (E,J) MI from a societal perspective using sigmoidal (A,B,C,D,E) and constant (F,G,H,I,J) vaccine efficacy profiles. The maximum PPD for

nirsevimab and RSVpreF correspond to estimates reported in Table A4. Black dashed-line corresponds to the WTP threshold of $50,000 per QALY gained.

##### 7.3. 80% coverage of nirsevimab and 60% coverage of maternal vaccination without monetary loss of life

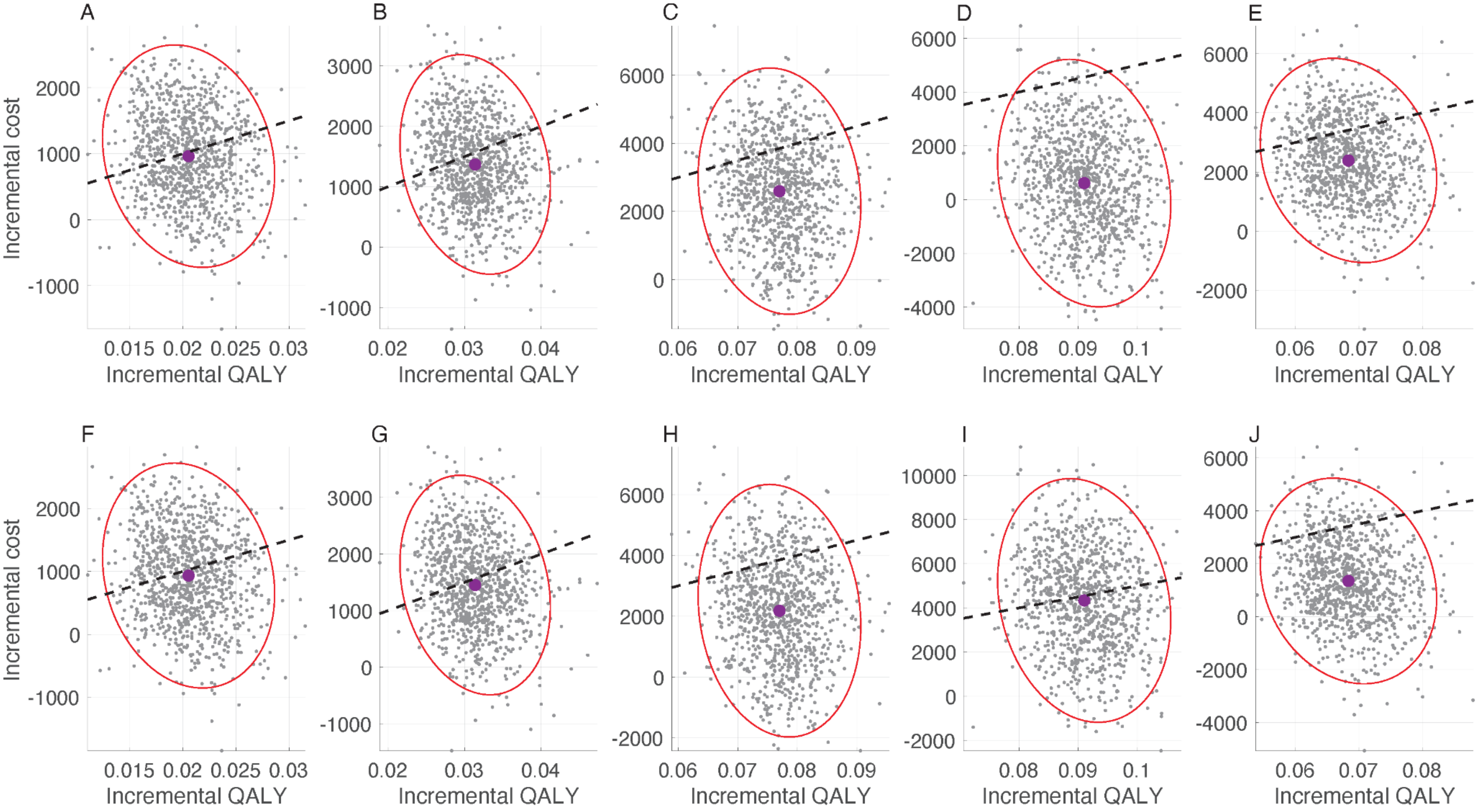

**Figure A31.** Cost-effectiveness planes for standalone immunisation programs: (A,F) L1; (B,G) L2; (C,H) L3; (D,I) L4, and (E,J) MI from healthcare (A,B,C,D,E) and societal (F,G,H,I,J) perspectives using sigmoidal vaccine efficacy profiles. The maximum PPD for

nirsevimab and RSVpreF correspond to estimates reported in Table A7. Black dashed-line corresponds to the WTP threshold of $50,000 per QALY gained.

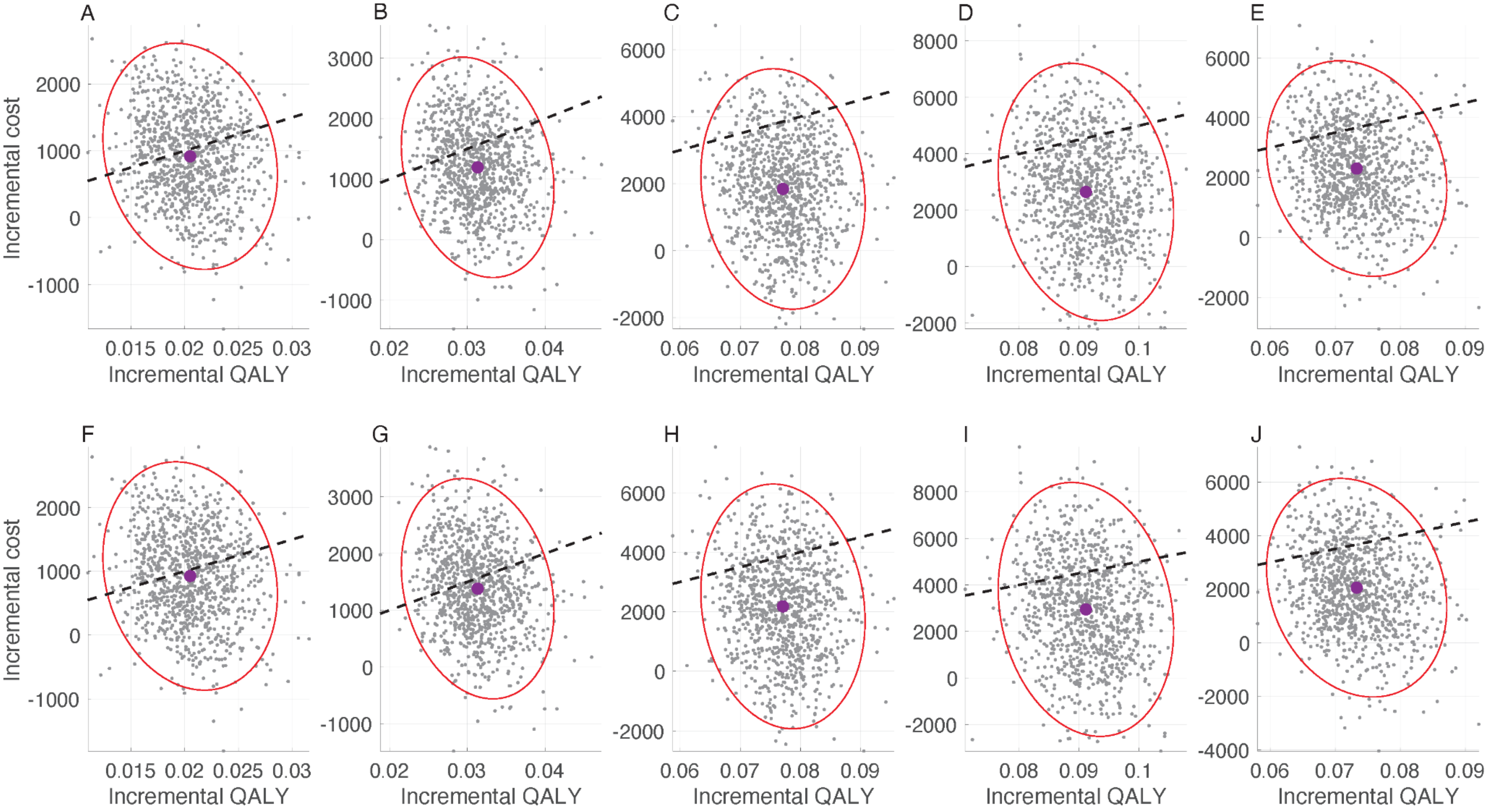

**Figure A32.** Cost-effectiveness planes for standalone immunisation programs: (A,F) L1; (B,G) L2; (C,H) L3; (D,I) L4, and (E,J) MI from healthcare (A,B,C,D,E) and societal (F,G,H,I,J) perspectives using constant vaccine efficacy profiles. The maximum PPD for

nirsevimab and RSVpreF correspond to estimates reported in Table A8. Black dashed-line corresponds to the WTP threshold of $50,000 per QALY gained.

##### 7.4. 80% coverage of nirsevimab and 60% coverage of maternal vaccination with monetary loss of life

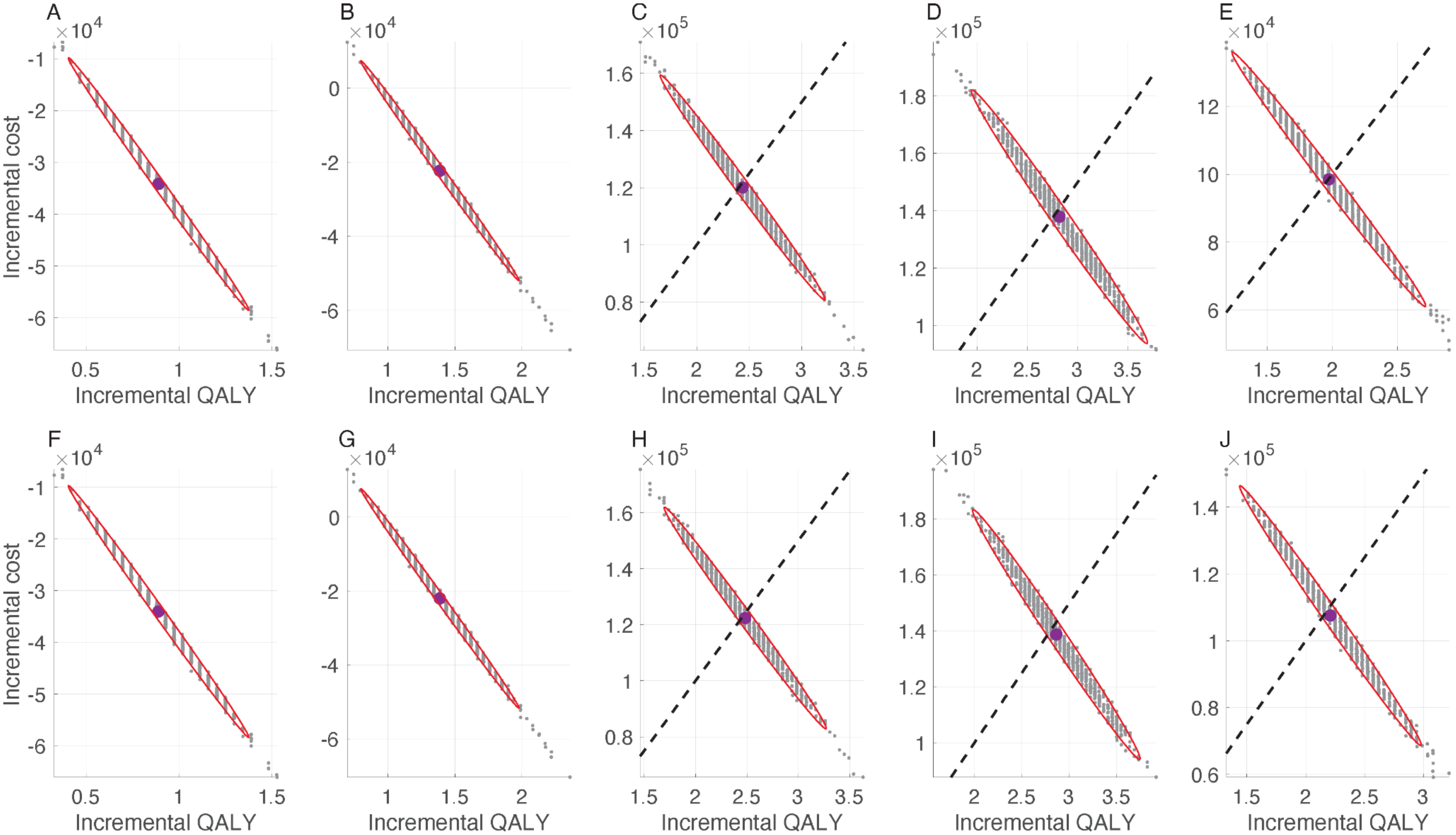

**Figure A33.** Cost-effectiveness planes for standalone immunisation programs: (A,F) L1; (B,G) L2; (C,H) L3; (D,I) L4, and (E,J) MI from a societal perspective using sigmoidal (A,B,C,D,E) and constant (F,G,H,I,J) vaccine efficacy profiles. The maximum PPD for

nirsevimab and RSVpreF correspond to estimates reported in Table A11. Black dashed-line corresponds to the WTP threshold of $50,000 per QALY gained.

#### 8. Cost-effectiveness planes for immunisation programs with the WTP of $30,000 per QALY gained

##### 8.1. 100% coverage of nirsevimab and 100% coverage of maternal vaccination without monetary loss of life

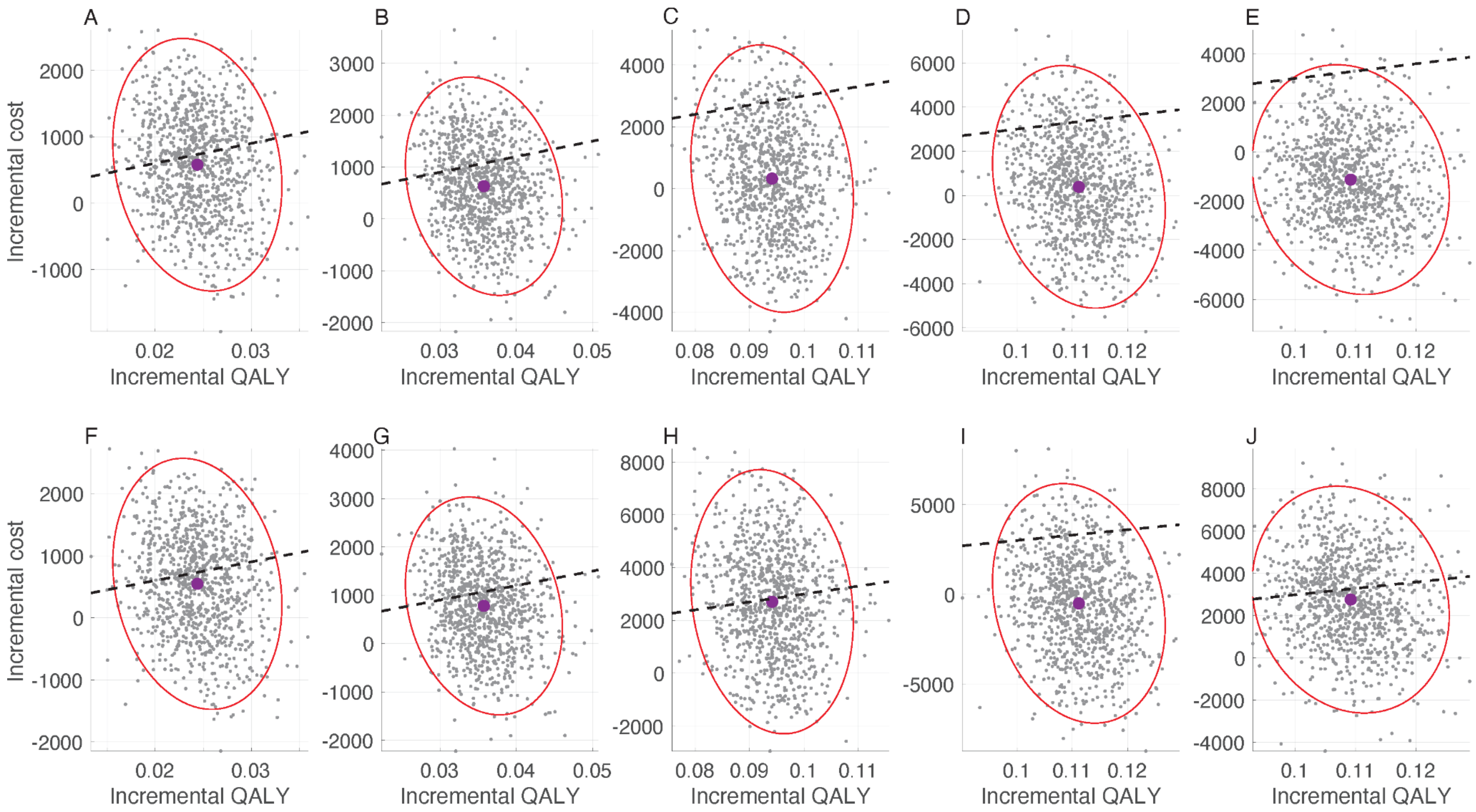

**Figure A34.** Cost-effectiveness planes for standalone immunisation programs: (A,F) L1; (B,G) L2; (C,H) L3; (D,I) L4, and (E,J) MI from healthcare (A,B,C,D,E) and societal (F,G,H,I,J) perspectives using sigmoidal vaccine efficacy profiles. The maximum PPD for nirsevimab and RSVpreF correspond to estimates reported in Table A13. Black dashed-line corresponds to the WTP threshold of $30,000 per QALY gained.

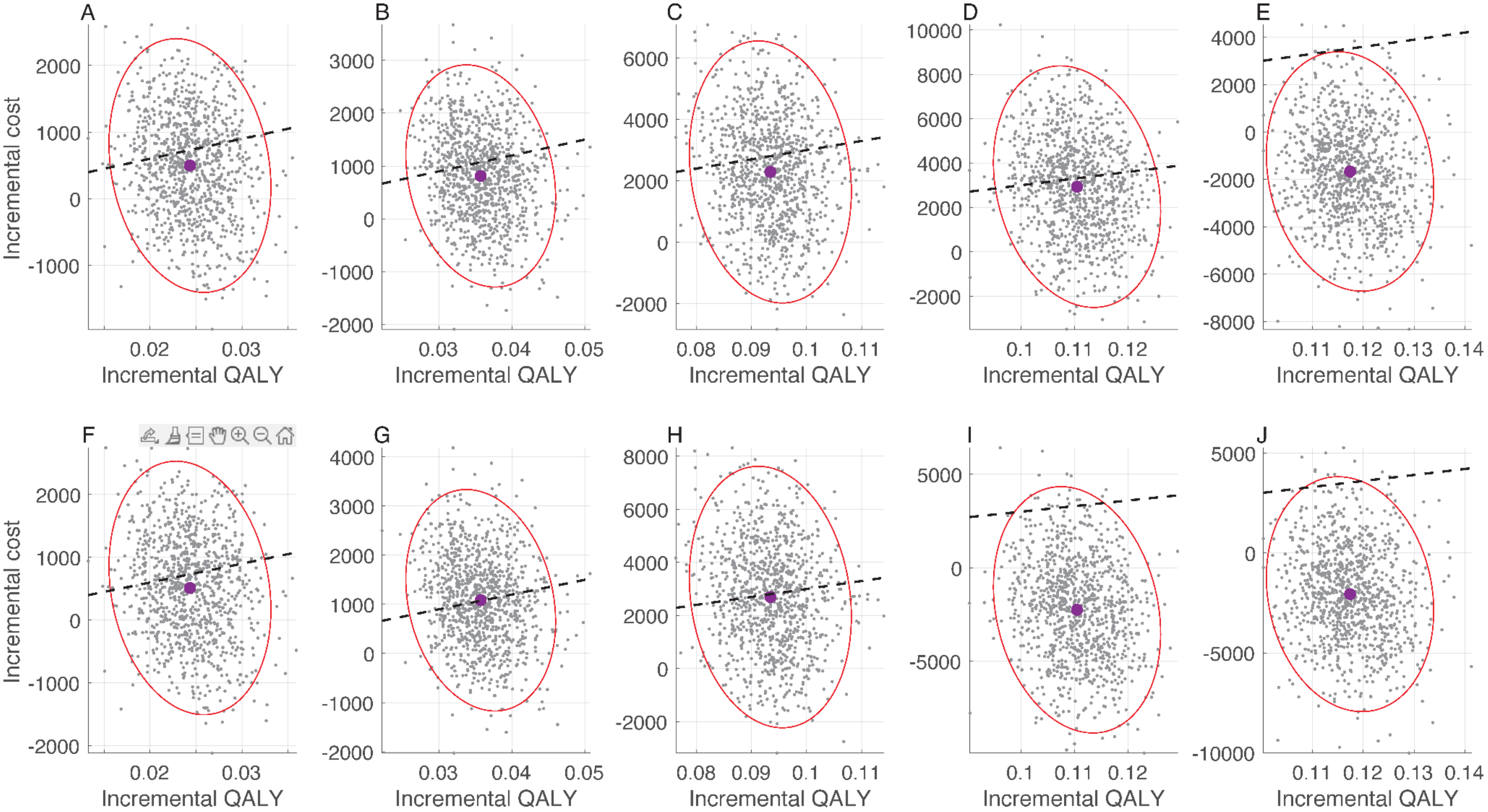

**Figure A35.** Cost-effectiveness planes for standalone immunisation programs: (A,F) L1; (B,G) L2; (C,H) L3; (D,I) L4, and (E,J) MI from healthcare (A,B,C,D,E) and societal (F,G,H,I,J) perspectives using constant vaccine efficacy profiles. The maximum PPD for nirsevimab and RSVpreF correspond to estimates reported in Table A14. Black dashed-line corresponds to the WTP threshold of $30,000 per QALY gained.

##### 8.2. 100% coverage of nirsevimab and 100% coverage of maternal vaccination with monetary loss of life

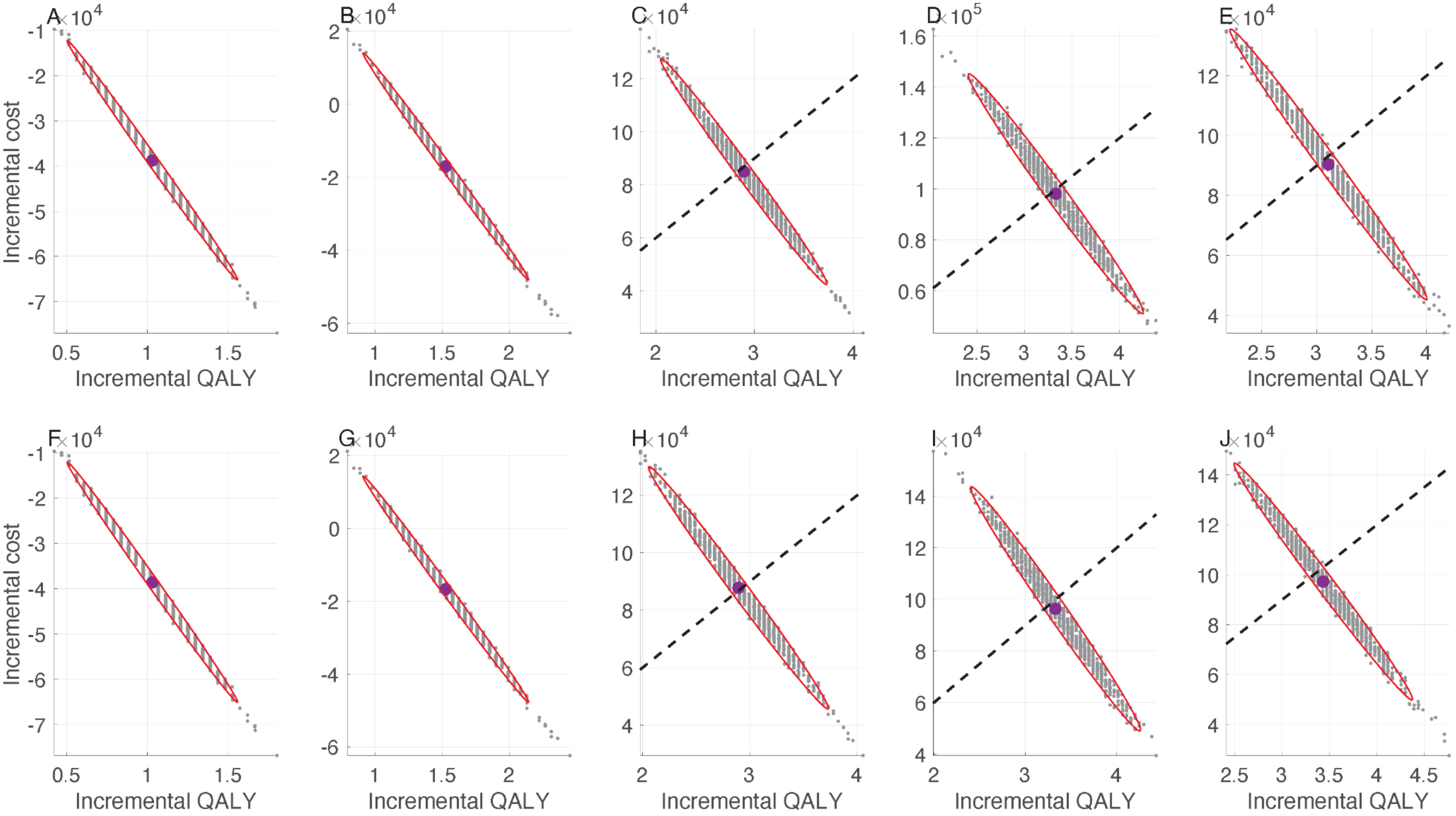

**Figure A36.** Cost-effectiveness planes for standalone immunisation programs: (A,F) L1; (B,G) L2; (C,H) L3; (D,I) L4, and (E,J) MI from a societal perspective using sigmoidal (A,B,C,D,E) and constant (F,G,H,I,J) vaccine efficacy profiles. The maximum PPD for nirsevimab and RSVpreF correspond to estimates reported in Table A17. Black dashed-line corresponds to the WTP threshold of $30,000 per QALY gained.

##### 8.3. 80% coverage of nirsevimab and 60% coverage of maternal vaccination without monetary loss of life

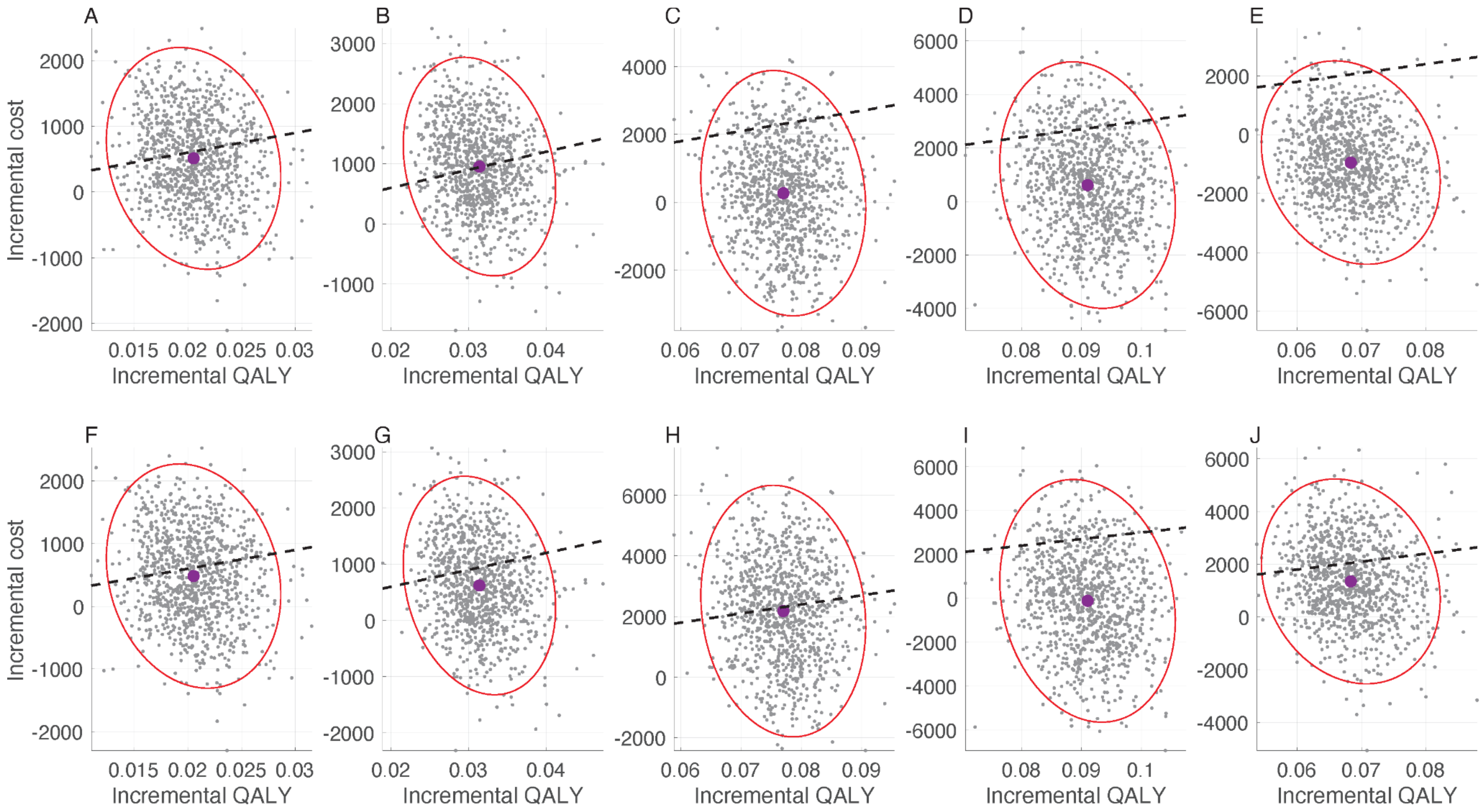

**Figure A37.** Cost-effectiveness planes for standalone immunisation programs: (A,F) L1; (B,G) L2; (C,H) L3; (D,I) L4, and (E,J) MI from healthcare (A,B,C,D,E) and societal (F,G,H,I,J) perspectives using sigmoidal vaccine efficacy profiles. The maximum PPD for

nirsevimab and RSVpreF correspond to estimates reported in Table A19. Black dashed-line corresponds to the WTP threshold of $30,000 per QALY gained.

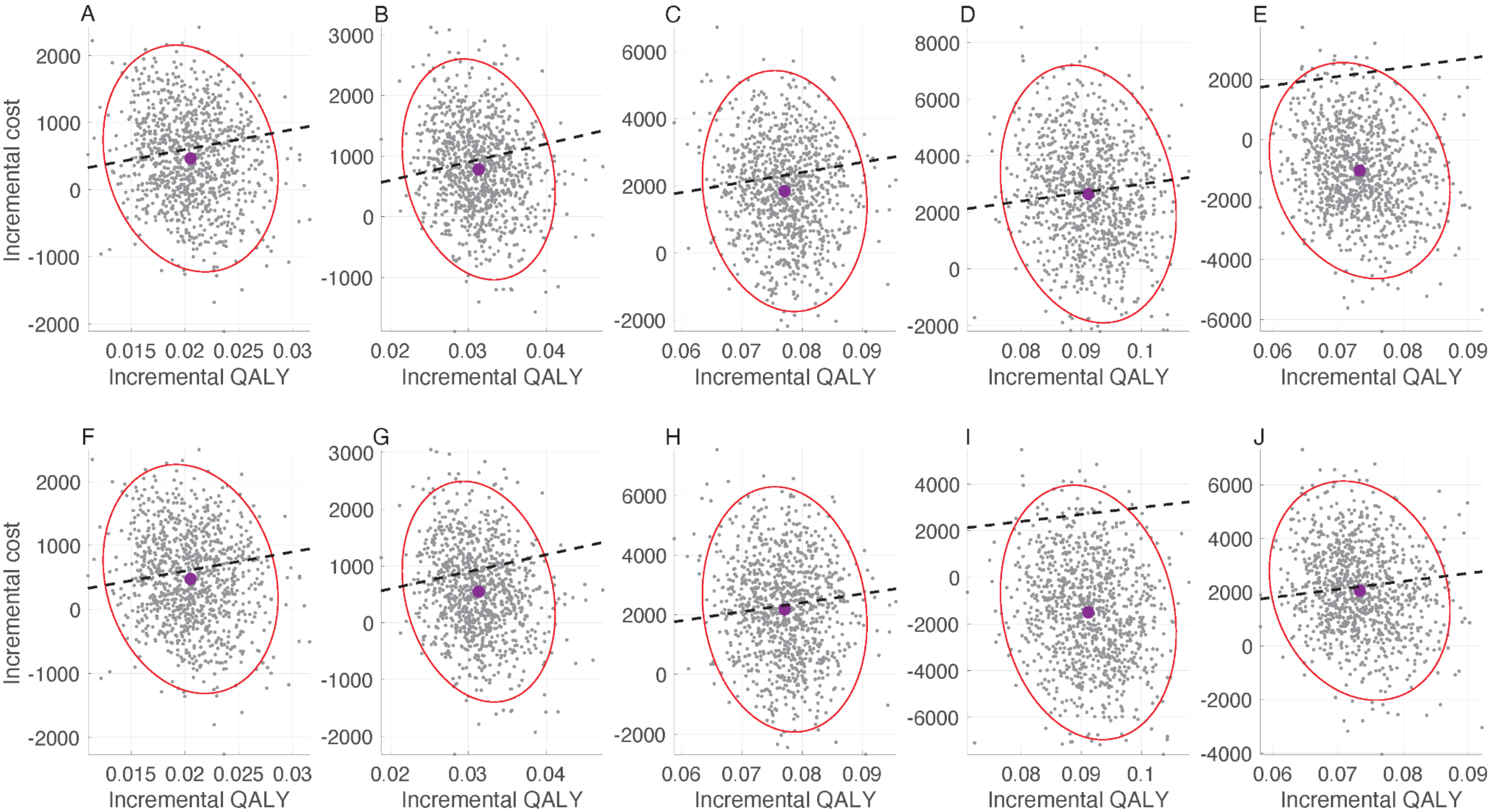

**Figure A38.** Cost-effectiveness planes for standalone immunisation programs: (A,F) L1; (B,G) L2; (C,H) L3; (D,I) L4, and (E,J) MI from healthcare (A,B,C,D,E) and societal (F,G,H,I,J) perspectives using constant vaccine efficacy profiles. The maximum PPD for

nirsevimab and RSVpreF correspond to estimates reported in Table A20. Black dashed-line corresponds to the WTP threshold of $30,000 per QALY gained.

##### 8.4. 80% coverage of nirsevimab and 60% coverage of maternal vaccination with monetary loss of life

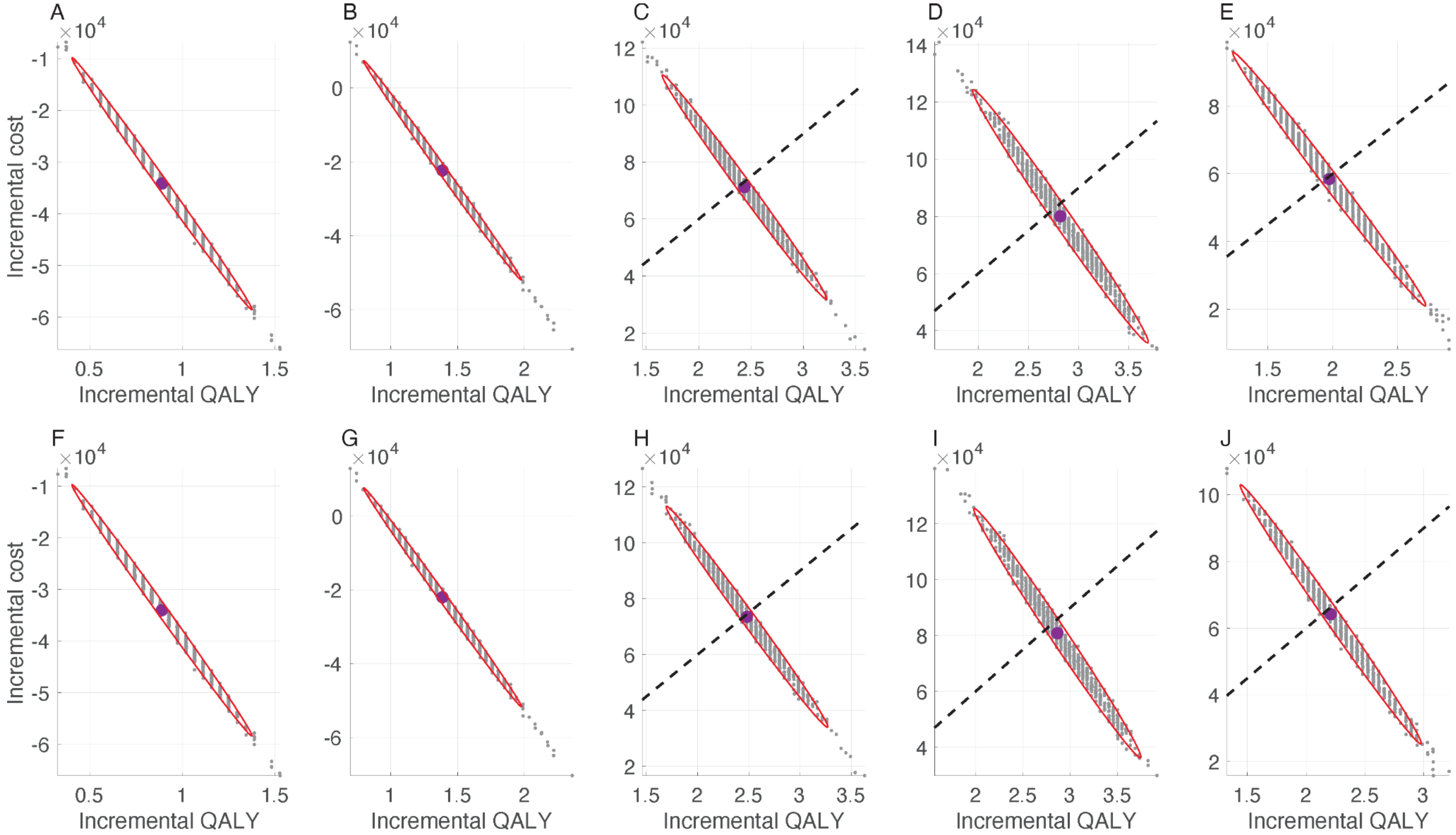

**Figure A39.** Cost-effectiveness planes for standalone immunisation programs: (A,F) L1; (B,G) L2; (C,H) L3; (D,I) L4, and (E,J) MI from a societal perspective using sigmoidal (A,B,C,D,E) and constant (F,G,H,I,J) vaccine efficacy profiles. The maximum PPD for

nirsevimab and RSVpreF correspond to estimates reported in Table A23. Black dashed-line corresponds to the WTP threshold of $30,000 per QALY gained.

#### 9. Cost-effectiveness planes for immunisation programs with the WTP of $70,000 per QALY gained

##### 9.1. 100% coverage of nirsevimab and 100% coverage of maternal vaccination without monetary loss of life

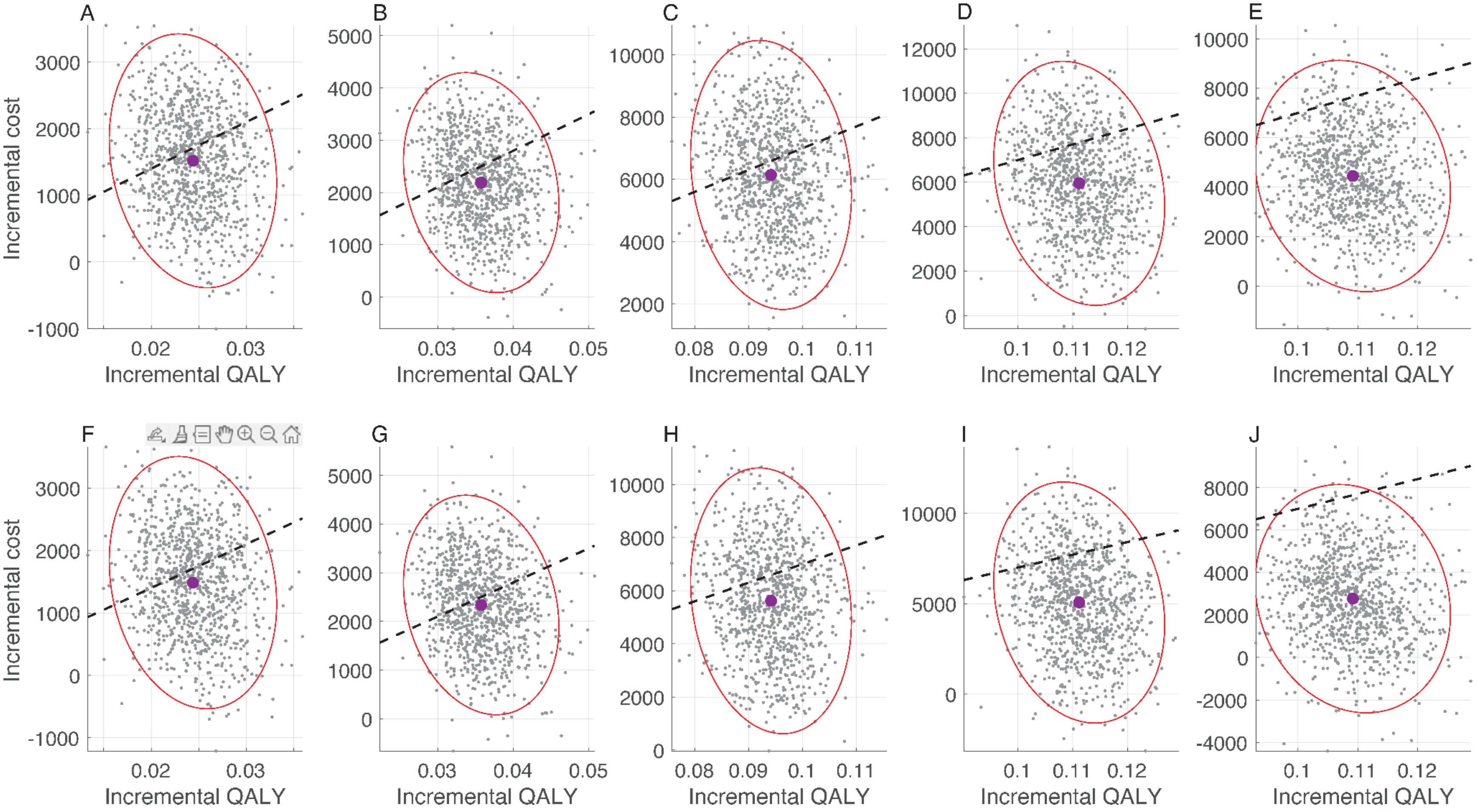

**Figure A40.** Cost-effectiveness planes for standalone immunisation programs: (A,F) L1; (B,G) L2; (C,H) L3; (D,I) L4, and (E,J) MI from healthcare (A,B,C,D,E) and societal (F,G,H,I,J) perspectives using sigmoidal vaccine efficacy profiles. The maximum PPD for nirsevimab and RSVpreF correspond to estimates reported in Table A25. Black dashed-line corresponds to the WTP threshold of $70,000 per QALY gained.

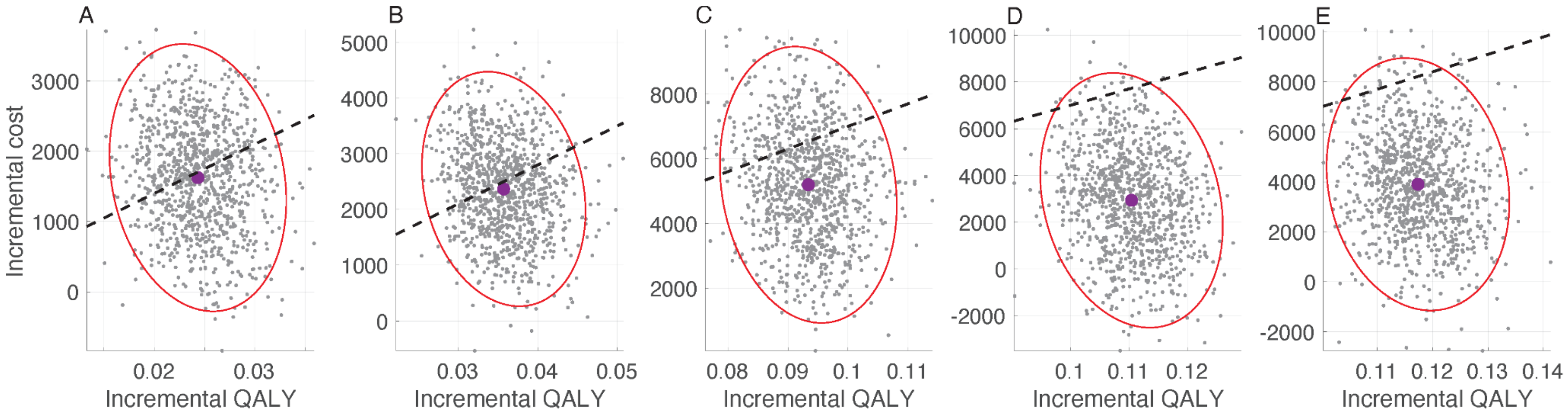

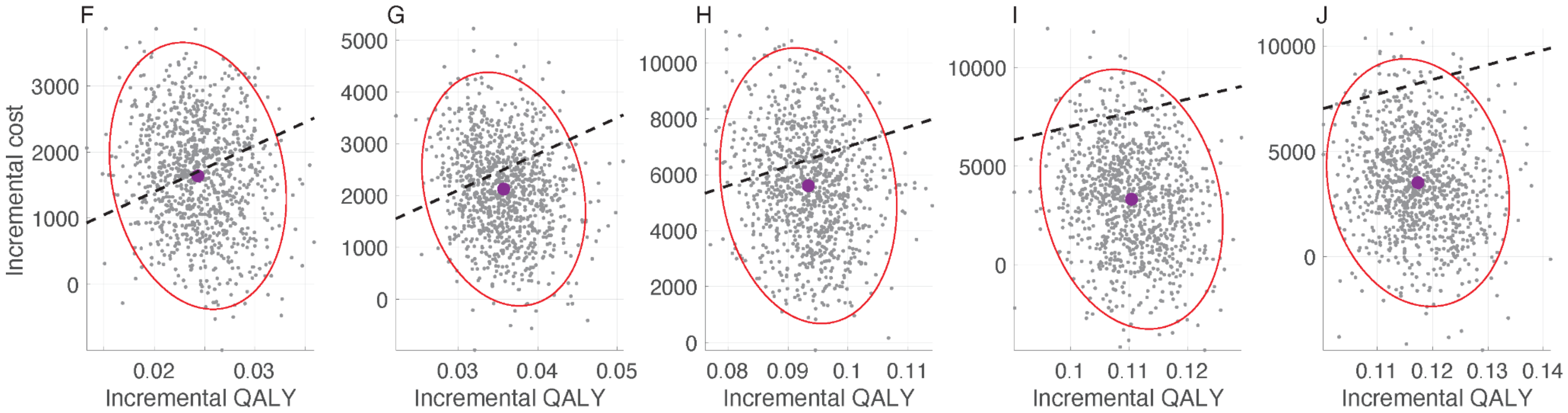

**Figure A41.** Cost-effectiveness planes for standalone immunisation programs: (A,F) L1; (B,G) L2; (C,H) L3; (D,I) L4, and (E,J) MI from healthcare (A,B,C,D,E) and societal (F,G,H,I,J) perspectives using constant vaccine efficacy profiles. The maximum PPD for

nirsevimab and RSVpreF correspond to estimates reported in Table A26. Black dashed-line corresponds to the WTP threshold of $70,000 per QALY gained.

##### 9.2. 100% coverage of nirsevimab and 100% coverage of maternal vaccination with monetary loss of life

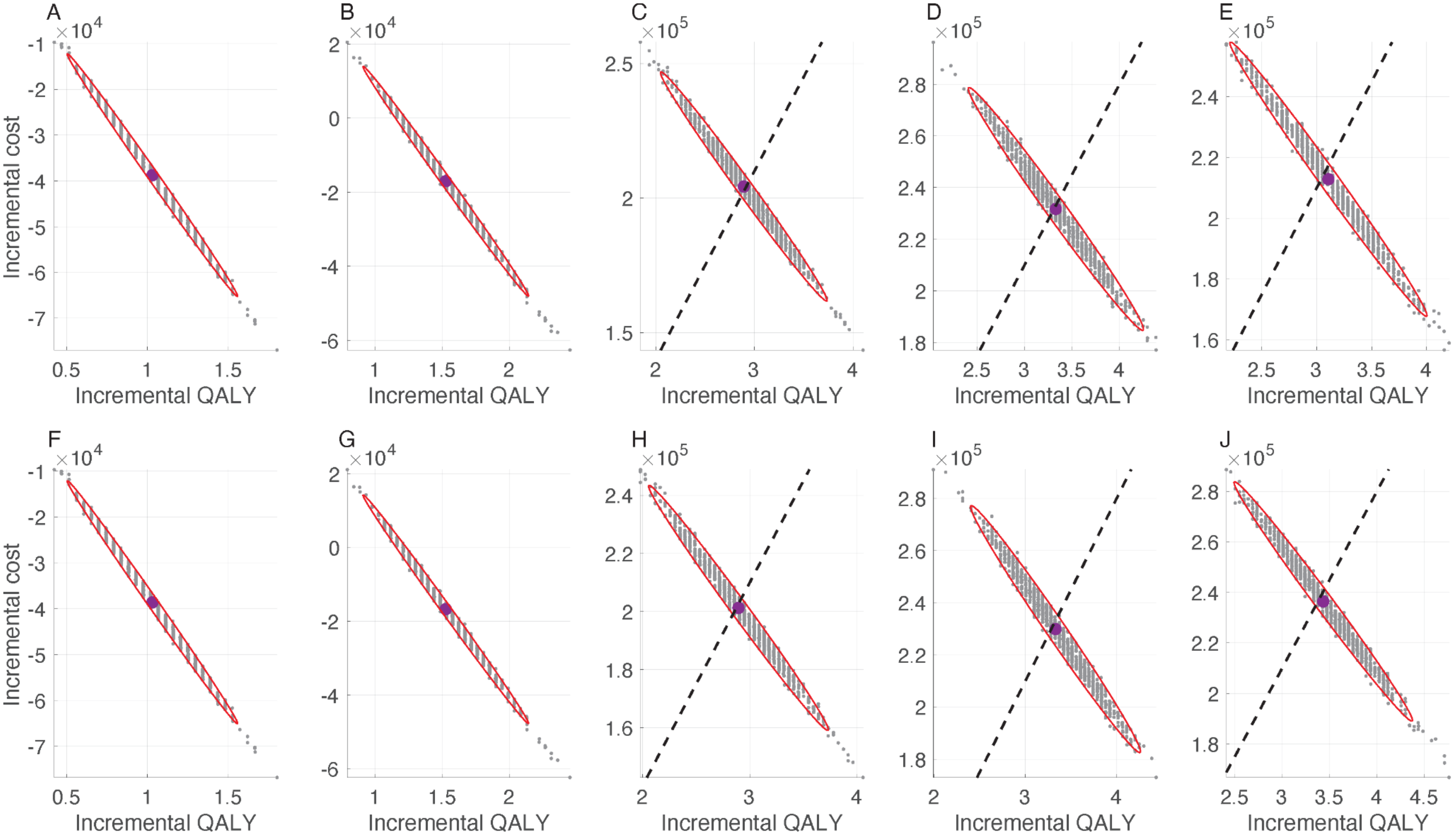

**Figure A42.** Cost-effectiveness planes for standalone immunisation programs: (A,F) L1; (B,G) L2; (C,H) L3; (D,I) L4, and (E,J) MI from a societal perspective using sigmoidal (A,B,C,D,E) and constant (F,G,H,I,J) vaccine efficacy profiles. The maximum PPD for

nirsevimab and RSVpreF correspond to estimates reported in Table A29. Black dashed-line corresponds to the WTP threshold of $70,000 per QALY gained.

##### 9.3. 80% coverage of nirsevimab and 60% coverage of maternal vaccination without monetary loss of life

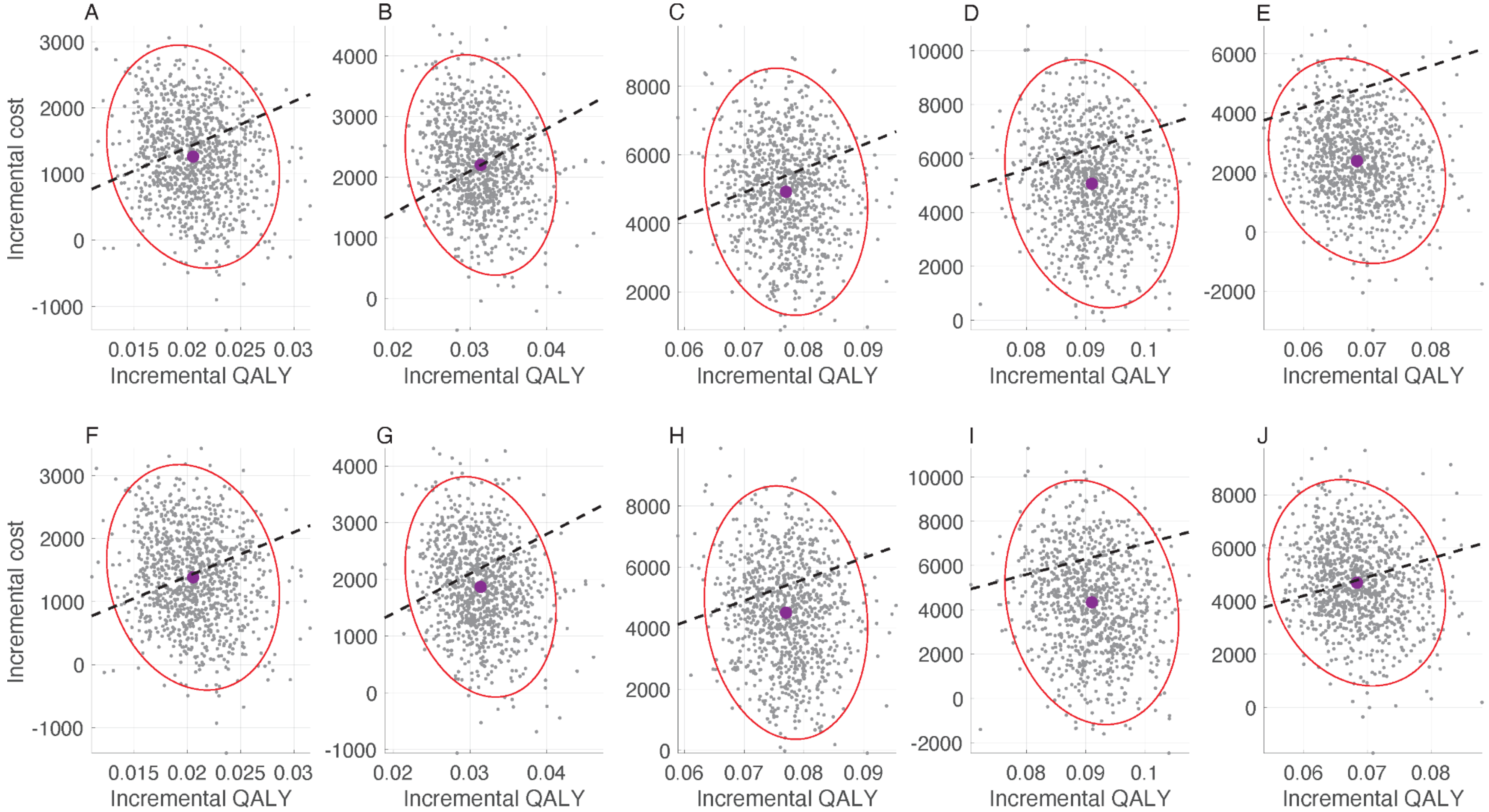

**Figure A43.** Cost-effectiveness planes for standalone immunisation programs: (A,F) L1; (B,G) L2; (C,H) L3; (D,I) L4, and (E,J) MI from healthcare (A,B,C,D,E) and societal (F,G,H,I,J) perspectives using sigmoidal vaccine efficacy profiles. The maximum PPD for

nirsevimab and RSVpreF correspond to estimates reported in Table A31. Black dashed-line corresponds to the WTP threshold of $70,000 per QALY gained.

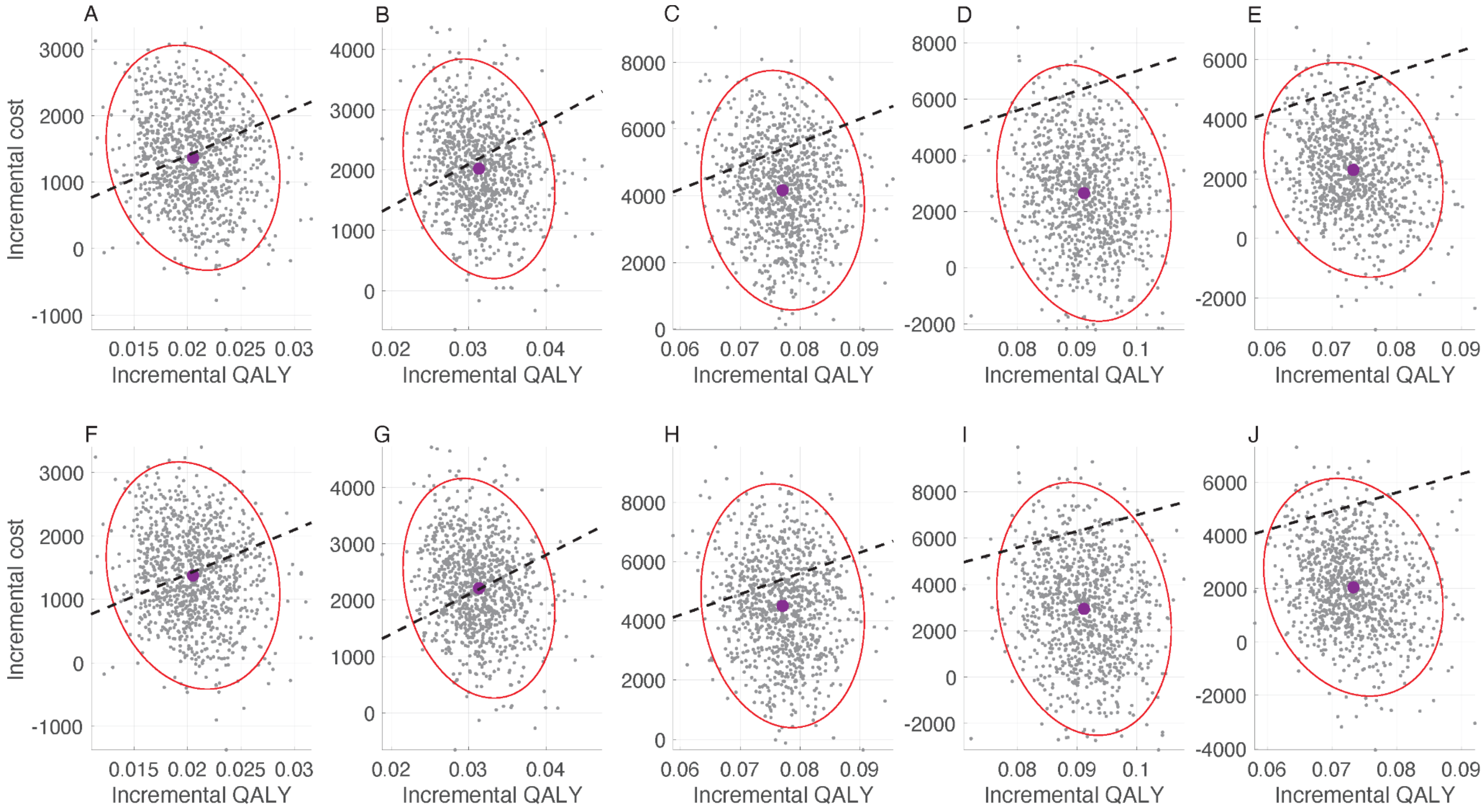

**Figure A44.** Cost-effectiveness planes for standalone immunisation programs: (A,F) L1; (B,G) L2; (C,H) L3; (D,I) L4, and (E,J) MI from healthcare (A,B,C,D,E) and societal (F,G,H,I,J) perspectives using constant vaccine efficacy profiles. The maximum PPD for nirsevimab and RSVpreF correspond to estimates reported in Table A32. Black dashed-line corresponds to the WTP threshold of $70,000 per QALY gained.

#### 9.4. 80% coverage of nirsevimab and 60% coverage of maternal vaccination with monetary loss of life

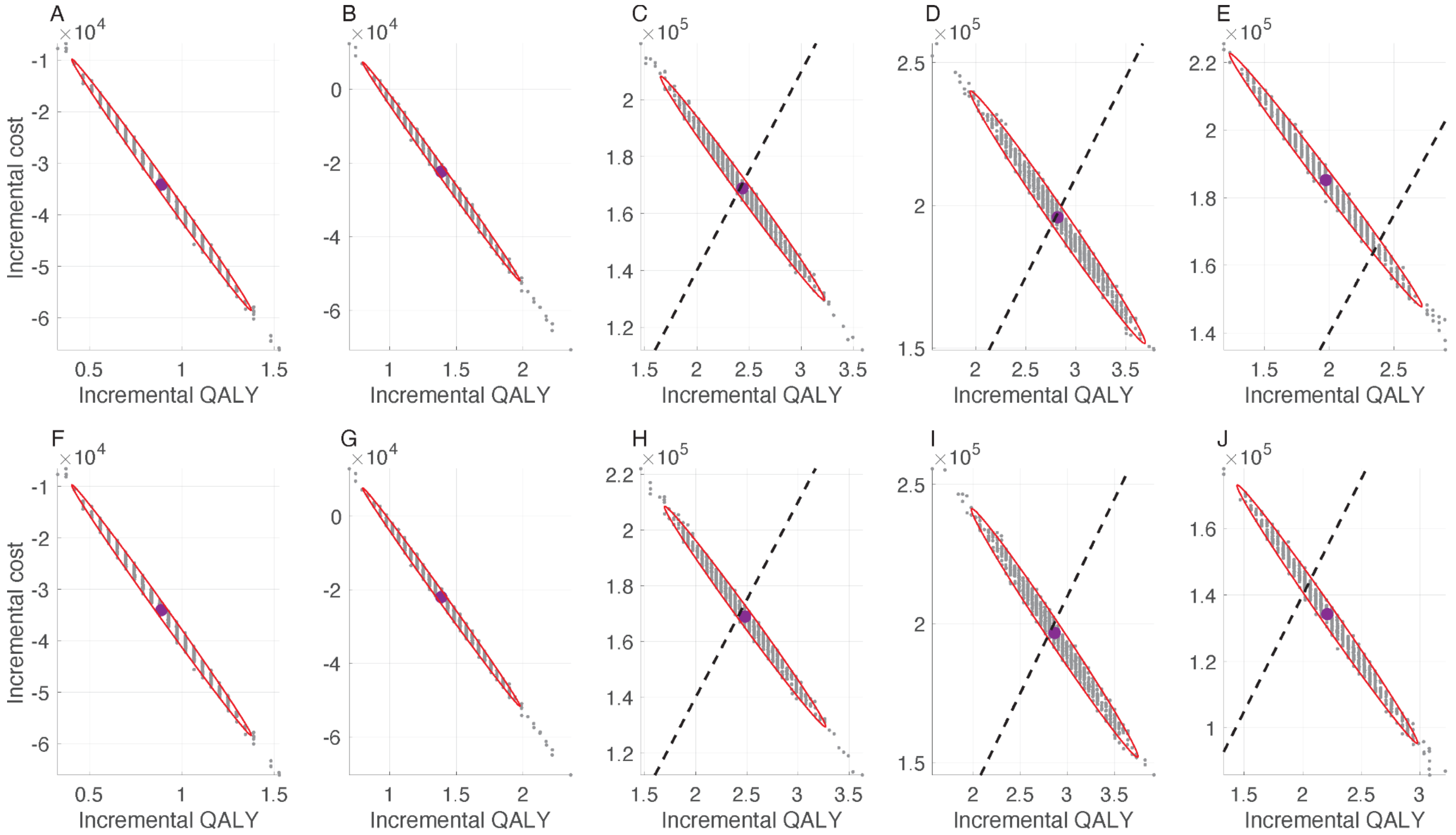

**Figure A45.** Cost-effectiveness planes for standalone immunisation programs: (A,F) L1; (B,G) L2; (C,H) L3; (D,I) L4, and (E,J) MI from a societal perspective using sigmoidal (A,B,C,D,E) and constant (F,G,H,I,J) vaccine efficacy profiles. The maximum PPD for nirsevimab and RSVpreF correspond to estimates reported in Table A35. Black dashed-line corresponds to the WTP threshold of $70,000 per QALY gained.

#### References

1. Rafferty, E. *et al.* Evaluating the Individual Healthcare Costs and Burden of Disease Associated with RSV Across Age Groups. *PharmacoEconomics* **40**, 633–645 (2022).
2. Vadlamudi, N. K. Characterizing Respiratory Syncytial Virus-Related Pediatric Disease Severity in Canada. (2022).
3. Parikh, R. C. *et al.* Chronologic Age at Hospitalization for Respiratory Syncytial Virus Among Preterm and Term Infants in the United States. *Infect. Dis. Ther.* **6**, 477–486 (2017).
4. Ratti, C., Greca, A. D., Bertoncelli, D., Rubini, M. & Tchana, B. Prophylaxis protects infants with congenital heart disease from severe forms of RSV infection: an Italian observational retrospective study: Palivizumab prophylaxis in children with congenital heart disease. *Ital. J. Pediatr.* **49**, 4 (2023).
5. Paes, B. *et al.* Defining the Risk and Associated Morbidity and Mortality of Severe Respiratory Syncytial Virus Infection Among Infants with Chronic Lung Disease. *Infect. Dis. Ther.* **5**, 453–471 (2016).
6. Ortega-Sanchez, I. R. Economics of Preventing Respiratory Syncytial Virus Lower Respiratory Tract Infections (RSV- LRTI) among US Infants with Nirsevimab. (2023).
